# Active learning impairments in substance use disorders when resolving the explore-exploit dilemma: A replication and extension of previous computational modeling results

**DOI:** 10.1101/2023.04.03.23288037

**Authors:** Samuel Taylor, Claire A. Lavalley, Navid Hakimi, Jennifer L. Stewart, Maria Ironside, Haixia Zheng, Evan White, Salvador Guinjoan, Martin P. Paulus, Ryan Smith

**Affiliations:** Laureate Institute for Brain Research, Tulsa, OK, USA

**Keywords:** substance use disorder, learning rate, active inference, decision-making, computational psychiatry, explore-exploit dilemma, reinforcement learning, transdiagnostic, replication, prediction.

## Abstract

**Background:** Substance use disorders (SUDs) represent a major public health risk. Yet, our understanding of the mechanisms that maintain these disorders remains incomplete. In a recent computational modeling study, we found initial evidence that SUDs are associated with slower learning rates from negative outcomes and less value-sensitive choice (low “action precision”), which could help explain continued substance use despite harmful consequences.

**Methods:** Here we aimed to replicate and extend these results in a pre-registered study with a new sample of 168 individuals with SUDs and 99 healthy comparisons (HCs). We performed the same computational modeling and group comparisons as in our prior report (doi: 10.1016/j.drugalcdep.2020.108208) to confirm previously observed effects. After completing all pre-registered replication analyses, we then combined the previous and current datasets (N = 468) to assess whether differences were transdiagnostic or driven by specific disorders.

**Results:** Replicating prior results, SUDs showed slower learning rates for negative outcomes in both Bayesian and frequentist analyses (η^2^=.02). Previously observed differences in action precision were not confirmed. Logistic regressions including all computational parameters as predictors in the combined datasets could differentiate several specific disorders from HCs, but could not differentiate most disorders from each other.

**Conclusions:** These results provide robust evidence that individuals with SUDs have more difficulty adjusting behavior in the face of negative outcomes than HCs. They also suggest this effect is common across several different SUDs. Future research should examine its neural basis and whether learning rates could represent a new treatment target or moderator of treatment outcome.

## 1. INTRODUCTION

Substance use disorders (SUDs) have devastating mental and physical health consequences (1–5), and lead to large societal and financial burden (6). They also show high co-morbidity with depression and anxiety disorders, which can be both a cause and consequence of substance use (1–3). Yet, our understanding of the mechanisms that lead to substance use, and of those that moderate recovery and abstinence, remains poor. Given current relapse rates with existing treatments (7), a better understanding of these mechanisms may be crucial for identifying effective clinical targets; and they could also offer important insights regarding etiology and prevention.

Computational modeling offers a promising approach for characterizing these mechanisms in SUDs (8, 9), as it can disentangle processes underlying perception, learning, and decision-making that may contribute to these disorders (9, 10). A growing body of work has now applied this approach – highlighting several computational processes that differ from healthy participants and that may relate to vulnerability, severity, and/or treatment outcomes (11, 12). Much of this work pertains to deficits in “model-based” (i.e., goal-directed, prospective) control, which are thought to amplify habitual and impulsive choice (e.g., for recent reviews, see (12, 13)). Other studies have also highlighted deficits in computational mechanisms subserving perception (particularly with respect to internal bodily states; i.e., interoception (14, 15)) and reinforcement learning (e.g., see (16–18)). Thus, this approach has already been helpful in elucidating a number of mechanisms that may underlie SUDs.

Building on this body of work, we recently applied a computational approach to examine the way individuals with SUDs balance information-seeking and reward-seeking (19). Relative to healthy comparisons, modeling revealed slower learning rates for negative outcomes across multiple SUDs, as well as faster learning rates for positive outcomes and less precise (i.e., less value-sensitive) action selection. This pattern of results was also moderately stable when assessed in the same participants one year later (20). These findings suggested that substance use might persist despite painful life consequences due to slower learning from these negative outcomes. Returns to drug use after improvements in treatment could also be caused by imprecise action selection mechanisms (i.e., switching away from actions with positive outcomes). In the current pre-registered study (https://osf.io/u3sbg), we assess the reliability of these findings in a new, independent sample with similar characteristics to those in the original study. The same task and computational analyses were applied, and we report which results do and do not replicate. We subsequently extend these results by combining the two datasets, providing power to assess whether effects are driven by specific SUDs or co-morbid affective disorders or whether they represent a transdiagnostic marker across SUDs.

## 2. METHODS

### 2.1 Participants

Participants were sampled from the confirmatory dataset (N=550) of the larger Tulsa 1000 (T1000) study (21). The T1000 project recruited a longitudinal community sample with dimensional measures designed around the NIMH Research Domain Criteria framework. The sample in total consists of 1050 individuals, ages 18-55, recruited through radio, electronic media, word-of-mouth, and treatment center referrals. Participants were screened through various dimensional psychopathology measures, including Patient Health Questionnaire (PHQ-9 (22)) ≥ 10, Overall Anxiety Severity and Impairment Scale (OASIS (23)) >= 8, and/or Drug Abuse Screening Assessment (DAST-10 (24)) ≥ 3. Healthy comparisons (HCs) did not show elevated symptoms on any of the previous measures. Exclusion criteria for participants were: (i) positive tests for drug use, (ii) meeting criteria for psychotic, bipolar, or obsessive-compulsive disorders, (iii) a history of moderate-to-severe traumatic brain injury, neurological disorders, or severe or unstable medical conditions, (iv) active suicidal intent or plan, or (v) change in medication dose within 6 weeks. Full inclusion/exclusion criteria are reported in (21). The study was approved by the Western Institutional Review Board. All participants provided written informed consent prior to completion of the study protocol, in accordance with the Declaration of Helsinki, and were compensated for participation. ClinicalTrials.gov identifier: #NCT02450240.

Participants were divided into two groups: participants with one or more SUDs (with or without co-morbid anxiety/depression; N = 168) and participants without any mental health diagnosis (HCs; N = 99). Diagnoses were determined according to DSM-IV or DSM-5 criteria using the Mini International Neuropsychiatric Inventory (MINI) (25) and confirmed through clinical conferences with a board-certified psychiatrist. **Table 1** provides summary statistics for demographic and clinical measures, as well as two-sample t-tests assessing group differences (and corresponding Bayes factors [BFs], calculated using the BayesFactor package in R (26)). **Table 2** provides a summary of specific diagnoses within the SUDs group. **Table S1.1** in **Supplementary Materials 1** compares the demographic characteristics and clinical measures in this sample to those in our previous study (19). The only difference observed was that SUDs in the current sample had lower scores than the previous sample on the Wide Range Achievement Test reading score (WRAT) (27), a measure of premorbid intelligence (*t*(253) = 2.27, *p* = 0.02). This measure was included because of expected differences in baseline intellectual functioning relative to HCs.

**Table 1:**
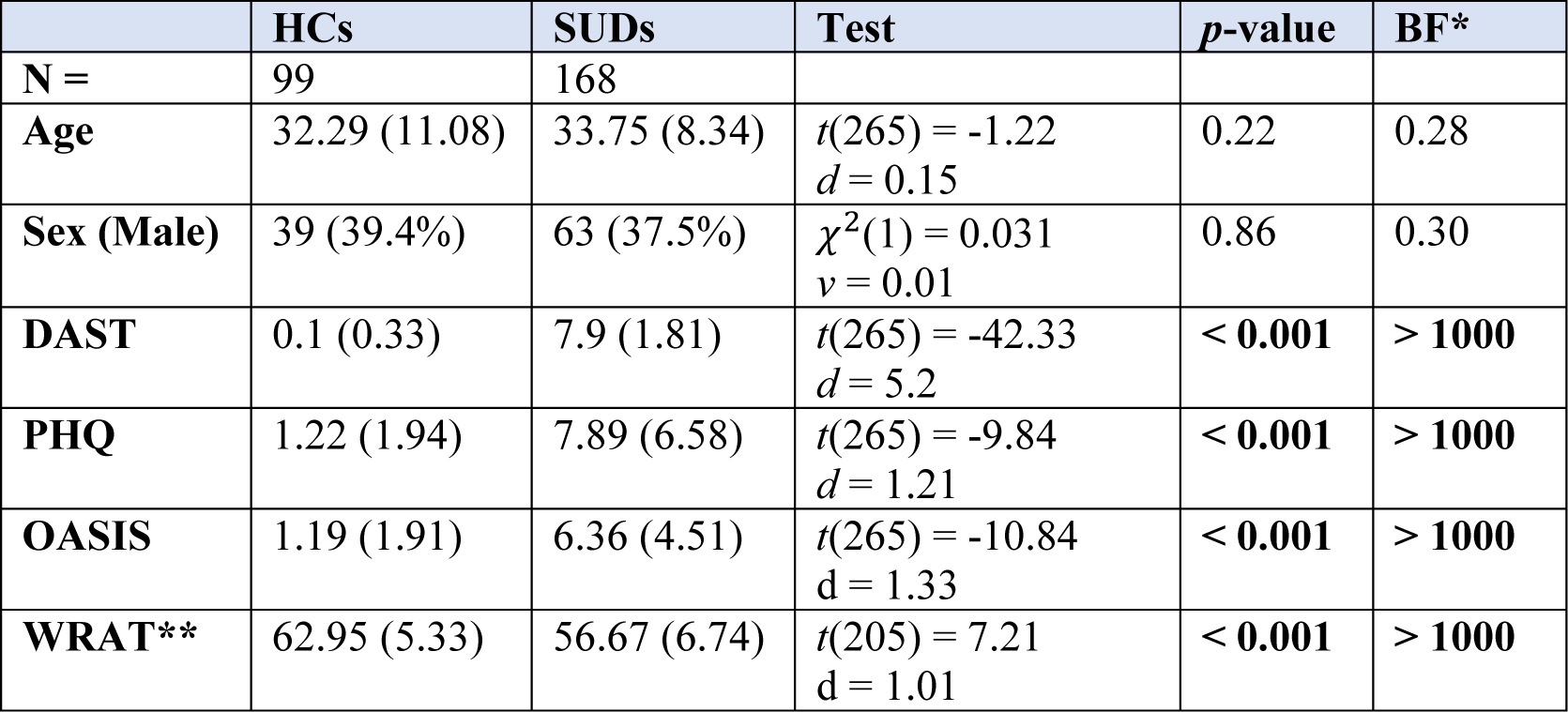
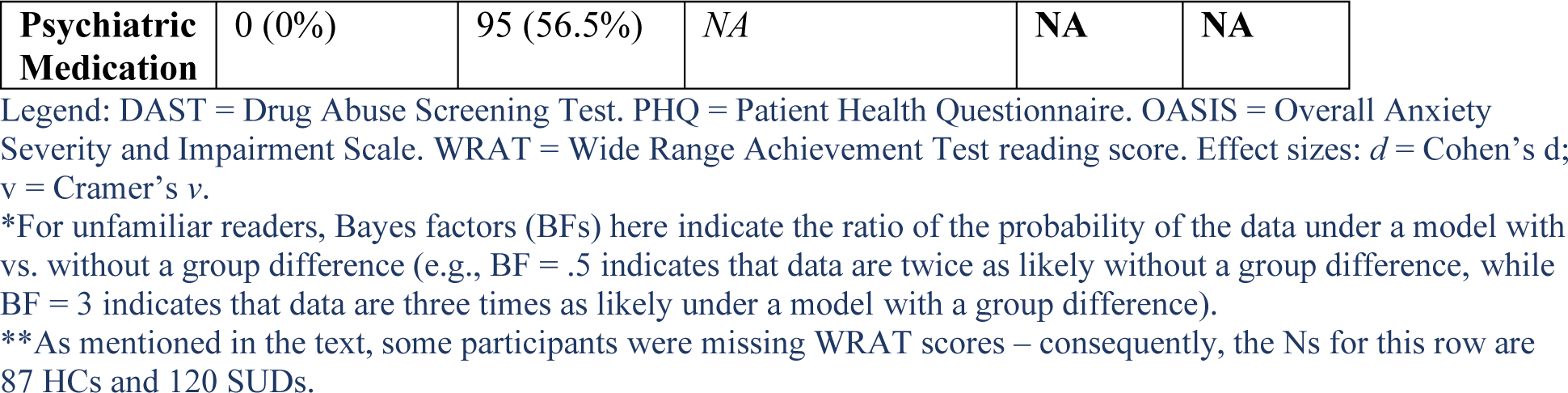
Demographic and Clinical Characteristics

|  | HCs | SUDs | Test | *p*-value | BF* |
| --- | --- | --- | --- | --- | --- |
| **N =** | 99 | 168 |  |  |  |
| **Age** | 32.29 (11.08) | 33.75 (8.34) | $t(265) = -1.22$<br>$d = 0.15$ | 0.22 | 0.28 |
| **Sex (Male)** | 39 (39.4%) | 63 (37.5%) | $\chi^2(1) = 0.031$<br>$v = 0.01$ | 0.86 | 0.30 |
| **DAST** | 0.1 (0.33) | 7.9 (1.81) | $t(265) = -42.33$<br>$d = 5.2$ | < **0.001** | > **1000** |
| **PHQ** | 1.22 (1.94) | 7.89 (6.58) | $t(265) = -9.84$<br>$d = 1.21$ | < **0.001** | > **1000** |
| **OASIS** | 1.19 (1.91) | 6.36 (4.51) | $t(265) = -10.84$<br>$d = 1.33$ | < **0.001** | > **1000** |
| **WRAT\*\*** | 62.95 (5.33) | 56.67 (6.74) | $t(205) = 7.21$<br>$d = 1.01$ | < **0.001** | > **1000** |
| **Psychiatric Medication** | 0 (0%) | 95 (56.5%) | NA | NA | NA |

Legend: DAST = Drug Abuse Screening Test. PHQ = Patient Health Questionnaire. OASIS = Overall Anxiety Severity and Impairment Scale. WRAT = Wide Range Achievement Test reading score. Effect sizes: $d$ = Cohen's $d$ ; $v$ = Cramer's $v$ .
\*For unfamiliar readers, Bayes factors (BFs) here indicate the ratio of the probability of the data under a model with vs. without a group difference (e.g., BF = .5 indicates that data are twice as likely without a group difference, while BF = 3 indicates that data are three times as likely under a model with a group difference).
\*\*As mentioned in the text, some participants were missing WRAT scores – consequently, the Ns for this row are 87 HCs and 120 SUDs.

**Table 2:**
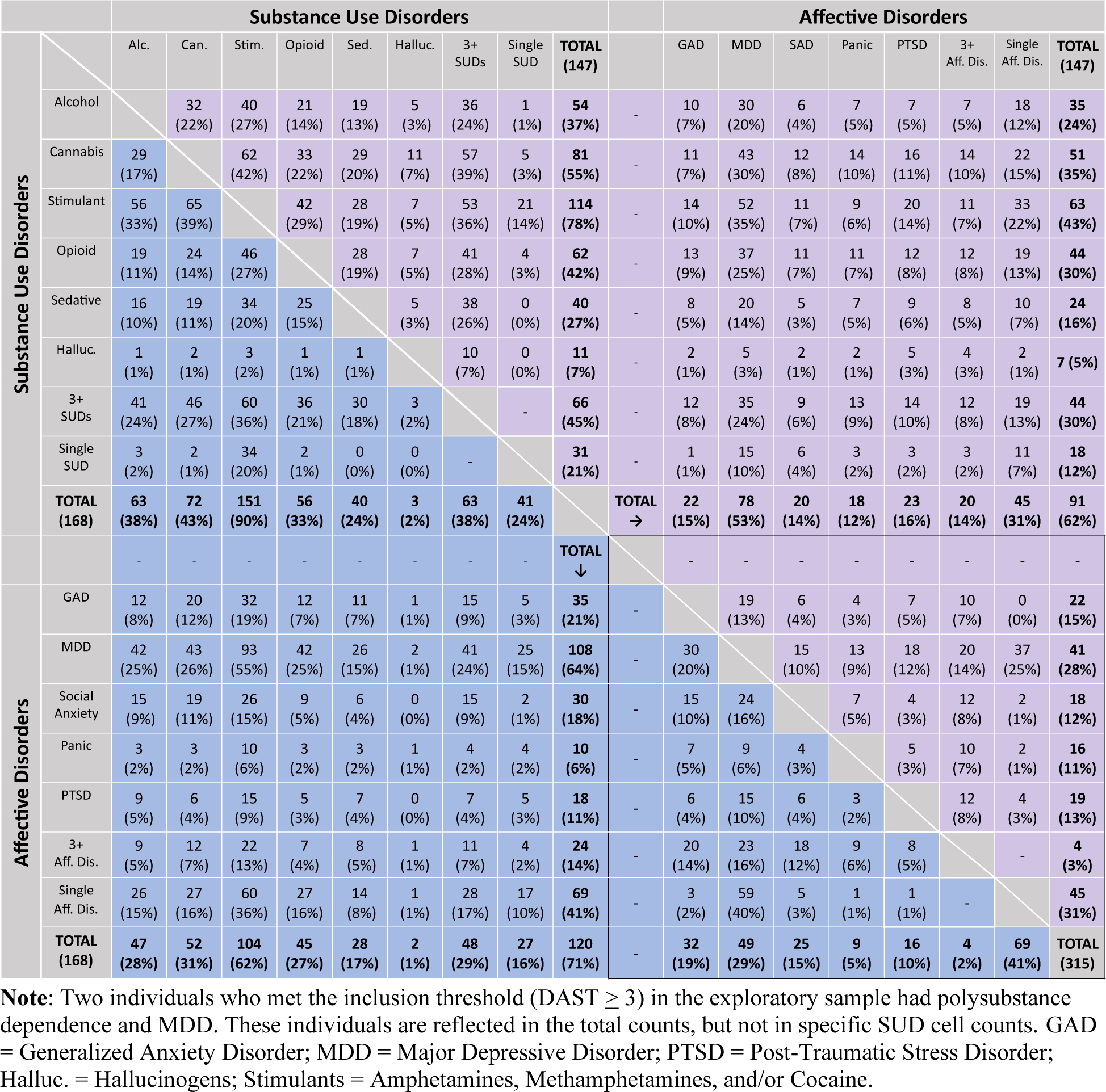
Lifetime DSM-IV/DSM-5 Psychiatric Diagnoses within SUDs

### 2.2 Three-Armed Bandit Task

Participants completed a standard three-armed bandit task (28). For 20 blocks of 16 sequential choices, they had to learn from trial-and-error which of three options was most likely to deliver a win (described in detail in our previous study (19); also see (20)). To maximize reward in each block, participants needed to strike a balance between sampling untested options (exploration) and capitalizing on the options that appeared most rewarding (exploitation). **Figure 1** shows the task interface and provides additional details.

**Figure 1:**
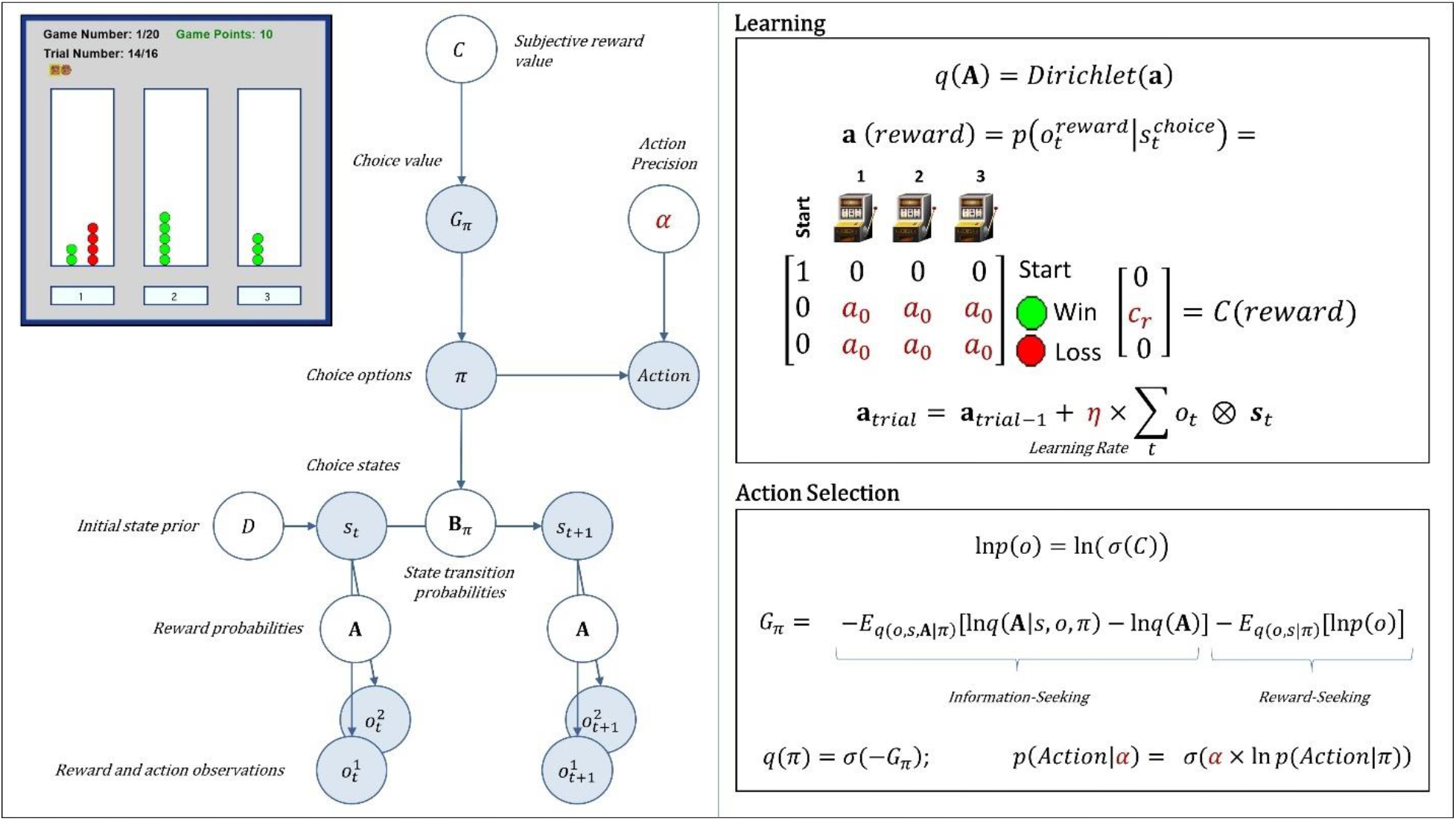
*Upper Left*: Screenshot of the task interface. In each block (N = 20), there were 16 trials where participants could choose the left, middle, or right options (buttons 1, 2, or 3 at the bottom). Each choice could lead to either a win (green circle) or a loss (red circle), which would appear above the chosen option. *Left*: A graphical depiction of the task model. Shaded circles denote inferred variables on each trial. White circles denote fixed model parameters. Arrows indicate dependencies between variables (see **Table S1.2** for a detailed explanation). *Right*: Model equations for learning and choice. Reward probabilities were encoded in a matrix **A**; i.e., 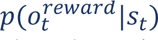. Approximate beliefs about these reward probabilities, *q*(**A**), were updated within a matrix **a**; i.e., 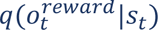. This matrix included a Dirichlet distribution over reward probabilities, the concentration parameters (*a_i,j_*) of which were updated after each observation according to the specified learning rule (shown in the top-left box, which includes the learning rate η). Policy values were based on expected free energy (*G*_π_), which favors policies that are expected to maximize both reward (encoded in *C*) and information gain (i.e., the magnitude of expected change in beliefs about reward probabilities [*q*(**A**)] after a new observation). Randomness in action selection given approximate posteriors over policies, *q*(π), was controlled by an action precision parameter α (note that σ indicates a softmax function). All free parameters in the model are shown in red.

### 2.3 Computational Modeling

A detailed computational model description can be found in **Table S1.2**. In brief, learning and choice are implemented by a set of inference and decision rules drawn from the active inference framework, which is designed to capture information-seeking under uncertainty. A detailed description of the mathematical framework can be found in (29, 30). The task model includes a set of observations (e.g., wins/losses), the subjective value assigned to those observations, choice states (option 1, 2, and 3), the probabilities of each observation under each choice state (e.g., the reward probabilities), and policies (actions) controlling transitions between choice states on each trial.

Free parameters within the model offer mechanistic explanations for individual differences in choice behavior. These parameters are summarized in **Table 3** and shown within the learning and choice equations in **Figure 1**.

**Table 3:** Free Model Parameters

| Free Parameter Name | Variable | Description |
| --- | --- | --- |
| **Action Precision** | $\alpha$ | Controls the degree of randomness in behavior – lower values indicate that a participant is less likely to exhibit consistent choices in the same decision context (i.e., under the same expected reward probabilities). This stochasticity is associated with a random exploration strategy (information gathering via random sampling), but it can also index overarching decision uncertainty or insensitivity to the relative value of each option. |
| **Reward Sensitivity** | $c_r$ | Indicates the subjective value of a win. Lower values lead to the prioritization of information-seeking, promoting goal-based (directed) exploration. |
| **Learning Rate** | $\eta$ | Reflects the magnitude with which beliefs about reward probabilities are updated after each observation. This can be split into separate learning rates for observed wins and losses, reflecting the fact that participants can be more sensitive to wins or losses as sources of information in updating beliefs. |
| **Information Insensitivity** | $a_0$ | Encodes the base levels of confidence in beliefs about reward probabilities before making initial choices at the start of each block. Higher values lead to reduced directed exploration. They also effectively slow learning, as observed wins/losses have a reduced impact on the normalized belief distribution over reward probabilities. |

We used a standard variational Bayes algorithm (variational Laplace (31)) to find parameter estimates that maximize the probability of participant behavior under the model, while also minimizing overfitting with a complexity cost (32). We then performed Bayesian model comparison (33, 34) across a set of 10 nested model variants (shown in **Table S1.3**) to determine the best model. The estimates from this best model were then used in subsequent statistical analyses.

Results of parameter recoverability and model identifiability analyses are reported in previous work (20). Parameters show good recoverability across the region of parameter space describing participant behavior, and the best-fit model is identifiable within model comparison.

### 2.4 Statistical Analysis Procedure

#### 2.4.1 Primary Replication Analyses

Statistical analyses replicate those in our previous study (19) and were pre-registered (https://osf.io/u3sbg). We first ran correlations to confirm how model parameter estimates were related to descriptive behavioral measures, including number of wins, mean reaction times (RTs) (after trimming using iterative Grubbs at a threshold of p < 0.01 (35)), and stay vs. shift behavior after wins vs. losses (i.e., whether participants shift to a new option or stay at the current option after a win or loss on the prior trial).

Our primary analyses then used parametric empirical Bayes (PEB) (36) to test general linear models with each model parameter estimate as the outcome variable, and using age, sex, baseline intellectual functioning (WRAT scores), and diagnostic group (SUDs and HCs) as predictors. PEB analyses allow incorporation of both posterior parameter means and variances when assessing these group-level models, as opposed to using the means alone as point estimates.

As in our prior study, secondary frequentist analyses (equivalent t-tests and multiple regressions) were also performed to assess group differences in the posterior parameter means. Supplementary model comparisons with these regressions, using Bayes factors (BFs) to compare the probability of the data under models with different possible combinations of predictors, were also performed (see note below **Table 4**).

**Table 4:** Parameter values (mean and SD) and group comparison

| Parameter | HCs | SUDs | Coefficients<br>& *t*-values | *p*-<br>value | Partial<br>$\eta^2$ | BF* |
| --- | --- | --- | --- | --- | --- | --- |
| N = | 99 | 168 |  |  |  |  |
| **Action Precision** | 2.61<br>(1.06) | 2.38<br>(1.48) | b = -0.2;<br>*t*(263) = -1.2 | 0.23 | 0.01 | Best Model (M1): Age (BF = 0.47)<br>M2: Age + Group (BF = 0.08) |
| **Reward Sensitivity** | 4.67<br>(1.87) | 4.43<br>(1.68) | b = -0.32;<br>*t*(263) = -1.47 | 0.14 | 0.01 | Best Model (M1): Age (BF = 562.51)<br>M2: Age + Group (BF = 29.76) |
| **Learning Rate (Wins)** | 0.48<br>(0.14) | 0.5<br>(0.15) | b = 0.02;<br>*t*(263) = 0.85 | 0.4 | 0.00 | Best Model (M1): Age (BF = 10.03)<br>M2: Age + Group (BF = 0.27) |
| **Learning Rate (Losses)** | 0.41<br>(0.14) | 0.37<br>(0.16) | b = -0.04;<br>*t*(263) = -2.09 | **0.04** | 0.02 | **Best Model (M1): Age + Group (BF = 13.68)**<br>M2: Age (BF = 13) |
| **Information Insensitivity** | 0.77<br>(0.33) | 0.83<br>(0.29) | b = 0.05;<br>*t*(263) = 1.41 | 0.16 | 0.01 | Best Model (M1): Age (BF = 3.23)<br>M2: Age + Group (BF = 0.17) |

**Note:** Inferential statistics reported here are based on linear models including age and sex as covariates. For analogous results in the subsample with available WRAT scores (including WRAT as an additional covariate), see **Supplementary Table S1.6**.
\*These Bayes factors reflect comparisons of evidence for models that did or did not include each possible combination of predictors (i.e., main effects of age, sex, group, and/or WRAT) in analogous Bayesian regressions. If the winning model included a main effect of group, the reported Bayes factor is relative to an intercept-only model, alongside the analogous Bayes factor of a model (M2) without the main effect of group (i.e., indicating the relative contribution of group effects in the model). If the winning model did not include group, this winning model and its Bayes factor are reported (relative to an intercept-only model), along with the Bayes factor for a model (M2) that added an effect of group. Bolded BF values indicate that the winning model included group as a main effect.

For comparison to model-based results, these same analyses were also performed on the descriptive behavioral measures. PEB analyses were performed in MATLAB (https://matlab.mathworks.com/). All other analyses were performed in R (2018; www.R-project.org/).

Our previous study employed propensity matching (on the basis of age, sex, and WRAT scores; using the optmatch package in R [https://cran.r-project.org/web/packages/optmatch/index.html]), which we initially planned to do here. However, the characteristics of this sample, and particularly the minimal overlap in WRAT scores between groups, prohibited our matching algorithm from finding a suitable subsample. Consequently, we only report analyses on the full sample using linear models that included age, sex, and WRAT as covariates. However, due to issues that arose during data collection, 12 HCs and 48 SUDs did not have available WRAT scores. In order to capitalize on the full sample, while also assessing possible effects of baseline intellectual functioning, we therefore ran each planned analysis once in the full sample (without including WRAT scores) and once in the subsample with available WRAT scores (while including this measure as a covariate). Results of analyses in the full sample are reported in detail below. Results within the analyses repeated in the subsample with WRAT scores are provided in detail within **Supplementary Materials 1 and 2**, but we also note important findings here.

Given the heterogeneity within our SUDs sample, we also planned to assess potential differences in computational parameters between different SUDs – specifically, differences between individuals with opioid use disorders and stimulant use disorders. While there was a sufficient number of individuals with stimulant use disorders without co-morbid opioid use disorders (N = 109), the low number of individuals with opioid use disorders that did not have co-morbid stimulant use disorders (N = 10) was too small to permit this analysis. However, after completing all pre-registered replication analyses, we combined the exploratory and confirmatory samples to examine disorder-specific effects (see section 2.4.3 and 3.5).

#### 2.4.2 Secondary Analyses

We ran correlations within SUDs to examine the relationship between model parameters and dimensional measures of symptom severity. Although not pre-registered, we also used t-tests to assess whether model parameters differed in medicated vs. unmedicated individuals in the SUDs group.

In addition to the 168 individuals with SUDs described above, the T1000 dataset also included 32 participants with an SUD diagnosis that had current symptom levels below the DAST ≥ 3 cutoff. These participants were not included in our primary analyses; however, they were used in some supplementary analyses described below. For more details about these additional participants, and comparison to the main sample, see **Table S1.4**.

#### 2.4.3 Disorder-Specific Analyses in Combined Samples

After completing all pre-registered replication analyses, we combined the exploratory and confirmatory datasets (N = 468) to ask questions about narrower diagnostic groups, which the previous or current dataset alone provided limited power to answer. Using logistic regressions, we first asked whether model parameters could differentiate HCs from those who had each of the specific substance use or affective disorders listed in **Table 2**. These regression models included each of the five parameters as predictors of diagnostic status (i.e., coding HCs = 0 and those with the specific disorder in question = 1, removing all other participants). We also examined those with no co-morbid SUDs when available sample size allowed, but high levels of co-morbidity did not permit this in most cases (i.e., only in stimulant use disorders; N = 55). These were considered post-hoc analyses done primarily to provide additional insights regarding whether specific overlapping disorders might drive the confirmed difference between HCs and SUDs overall. As such, we did not correct p-values for number of regressions; but we note that a Bonferroni correction for 11 regressions would entail a threshold of *p* < .0045. In similar logistic regressions, we then asked whether model parameters could differentiate one disorder vs. others (i.e., removing HCs and coding those with vs. without the specific disorder in question equal to 1 and 0, respectively).

#### 2.4.4 Predictive Categorization

Finally, to evaluate whether model parameters could predictively classify individuals into disorder groups, we trained logistic regression models (with the 5 parameters as joint predictors) on the exploratory sample and then assessed whether the trained models could accurately categorize individuals as either HCs or SUDs, both in general and for each specific SUD or affective disorder separately (i.e., HCs = 0, Disorder = 1 in each case, removing those without that the disorder being predicted). These predictive categorization analyses were performed using the *glm* function in R (https://www.rdocumentation.org/packages/stats/versions/3.6.2/topics/glm). Due to the different sample sizes per group, training data were also balanced using frequency weights. To assess performance, we report receiver operating characteristic curves (ROCs), reflecting the true positive rate (sensitivity) vs. false positive rate (1 -specificity) for different categorization thresholds, and the associated area under the curve (AUC). AUCs indicate how often a random sample will be assigned to the correct group with higher probability.

## 3. RESULTS

### 3.1 Model Validation

The winning model matched that found in our previous study (including action precision, reward sensitivity, separate learning rates for wins/losses, and insensitivity to information; **Table S1.3**), with a protected exceedance probability of 1.0. The average probability of participant actions under the model was .56 (SD = .10). The model assigned the highest probability to participant actions on 61% (SD = .10) of trials (note that chance = 33.33%).

Direct comparison of parameter values between the previous and current sample is provided in **Table S1.5**, with detailed results of associated linear models in **Table S2.1** and **S2.2** within **Supplementary Materials 2**. The parameter values in SUDs did not differ significantly between samples, nor were these values different between the two HC samples.

Relationships between model parameters and descriptive measures also replicated prior results (see **Figure 2**). Notable examples included: 1) number of wins was positively associated with action precision and reward sensitivity; 2) number of lose/shift choices was positively correlated with learning rate for losses; 3) stay choices were positively correlated with reward sensitivity and insensitivity to information; and 4) RTs were longer in those with slower learning rates for losses and faster in those with greater learning rates for wins, reward sensitivity, and insensitivity to information. The relationships with RTs are noteworthy, as model parameters are not fit to this aspect of behavior (i.e., the model itself does not simulate RTs).

**Figure 2:**
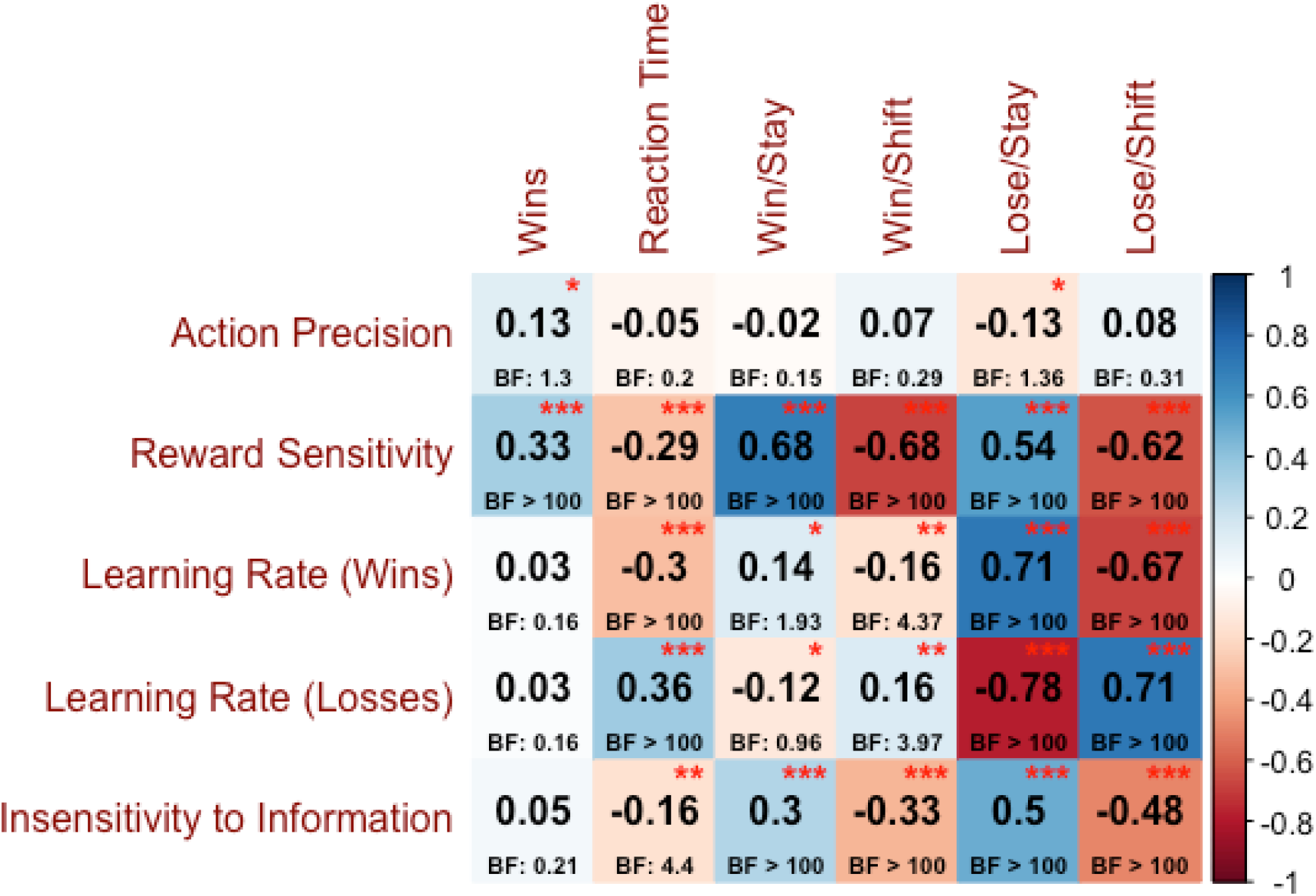
Correlations (and associated Bayes factors) between model parameter values and descriptive behavioral measures.

### 3.2 Group comparison

Replicating our prior results, PEB models with age and sex as covariates found strong evidence for group differences in learning rate for losses (slower in SUDs; posterior probability [*pp*] = 0.93; see **Figure 3**). These models also revealed some evidence for differences in reward sensitivity (greater in HCs; *pp* = 0.82). Note, however, that this latter finding was not present in our prior study. Including WRAT reading scores (in the subsample for which they were available) did not change these results (see **Figure S1.1**). See the *Additional Effects* section within **Supplementary Materials 1** for further effects observed in relation to age, sex, and WRAT scores.

**Figure 3:**
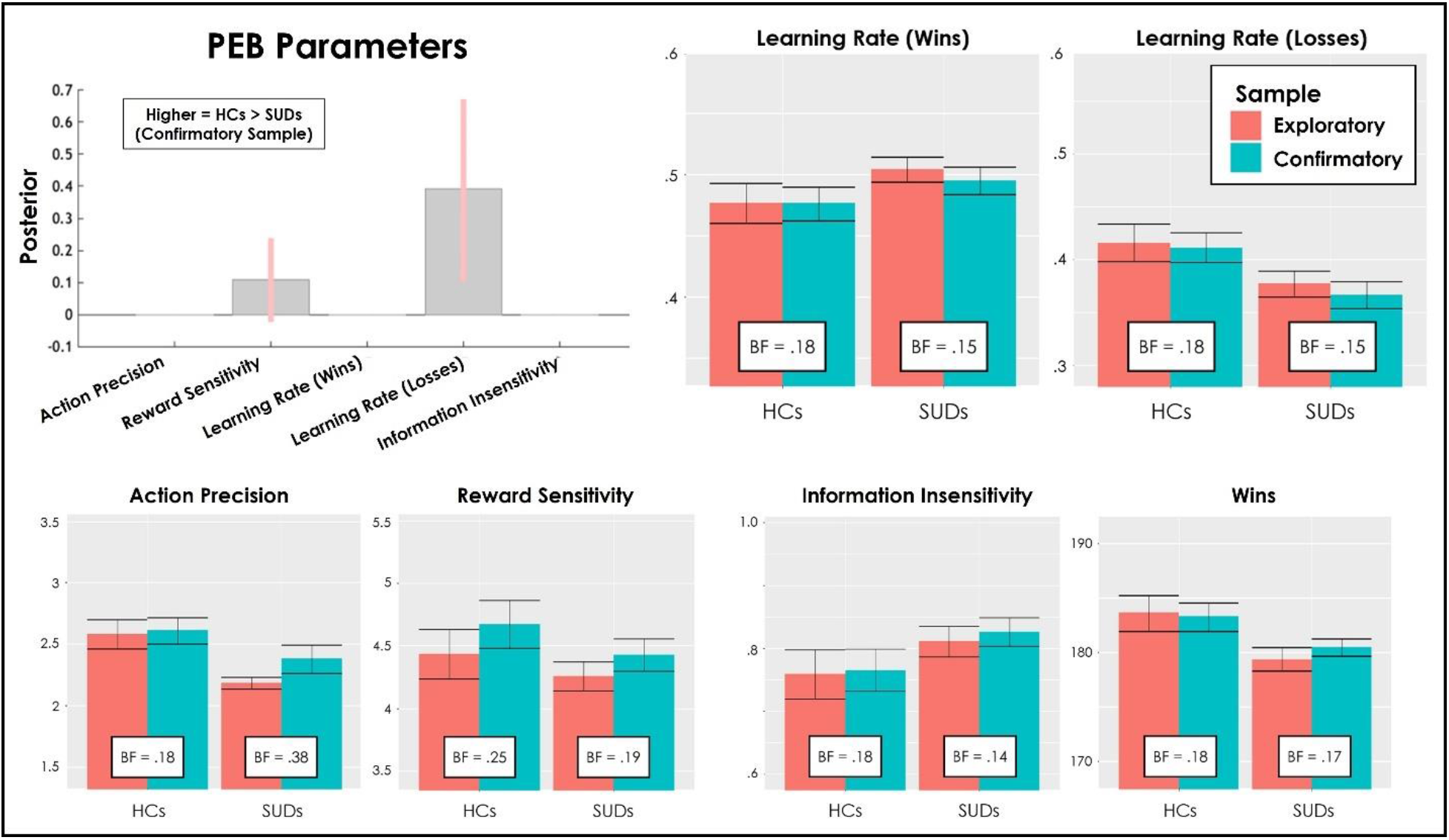
The top-left panel depicts results from Parametric Empirical Bayes (PEB) analyses showing the posterior means and credible intervals for effects of group (when accounting effects of age and sex). Positive values indicate greater values in HCs relative to SUDs. Learning rates are estimated/displayed in logit space, while all other parameters are in log space. Results replicate the group difference in learning rate for losses found in our prior study. All other panels compare the means and standard errors for each parameter in our prior study (exploratory sample; 54 HCs, 147 SUDs) to those in the current study (confirmatory sample; 99 HCs, 168 SUDs), when separating parameters by group. Also shown are Bayes factors (BFs) evaluating the evidence for differences between the two samples (BF < 1 indicates greater evidence for the absence of a difference). BFs indicated that the data were between 2.6 and 7.1 times more likely under a model with no difference between samples. For an identical plot restricting analyses to participants with available WRAT reading scores (and including these scores as predictors in the PEB analyses), see **Supplementary Figure S1.1**, which shows the same overall pattern of results (and in some cases shows stronger consistency between samples; most notably with action precision values).

As further assessment of successful replication, we also compared parameter estimates from the exploratory sample in our prior study to those in the present study for each group. We further computed BFs from Bayesian t-tests for each parameter/group to assess evidence for differences between samples. As shown in **Figure 3**, these BFs ranged from .14 to .38, indicating that the data are between 2.6 and 7.1 times more likely under a model with no difference between samples. This provides strong evidence that the new sample successfully replicates the findings in our prior study. Analogous results for the subsample with available WRAT scores are shown in **Figure S1.1**.

### 3.3 Supporting frequentist analyses

In complementary frequentist analyses, initial two-sample t-tests for each parameter confirmed the group difference in learning rates for losses (*t*(265) = 2.26, *p* = 0.02, *d* = 0.28), but did not support differences in any other parameter. **Table 4** reports results of subsequent regressions in the full sample analogous to the PEB models above, assessing whether age and sex could account for observed group differences. Equivalent Bayesian regressions were also used to identify the model (combination of predictors) for which the data provided the most evidence and how this compared to models either adding or removing potential group effects (see note below **Table 4**). Here we observed the same expected effects of group on learning rates for losses (lower values in SUDs).

Results of identical analyses on descriptive behavioral measures are reported in **Table 5**. We found greater wins in HCs than SUDs (*p* = 0.047; see **Table 7** and **Figure 3**), as found in our prior study. We also found fewer win/stay choices in SUDs, which was not observed previously. As in our prior study, we also repeated these analyses for the first and second halves of each task block separately (i.e., first 7 choices vs. subsequent choices) to assess periods where exploration vs. exploitation would be expected to dominate, respectively (see **Tables S1.8**). This showed that the SUDs group only made fewer win/stay choices than HCs in the second half of each game (*p* = .02; and greater win/shifts, *p* = .04), while HCs only showed greater wins in the first half of each game (*p* = .007). Additional relationships observed between age or sex and these measures are reported in **Table S2.3 and S2.4** and summarized in the *Additional Effects* section of **Supplementary Materials 1**.

**Table 5:** Descriptive behavioral measures and group comparison

| Measure | HCs | SUDs | Coefficients & *t*-values | *p*-value | Partial $\eta^2$ | BF* |
| --- | --- | --- | --- | --- | --- | --- |
| **N =** | 99 | 168 |  |  |  |  |
| **Wins** | 183.25<br>(13.19) | 180.44<br>(10.65) | b = -2.96;<br>*t*(263) = -2.00 | **0.047** | 0.01 | Best Model (M1): Group (BF = 0.77) |
| **Mean RT** | 0.56<br>(0.2) | 0.53<br>(0.25) | b = -0.03;<br>*t*(263) = -1.13 | 0.26 | 0.00 | Best Model (M1): Sex (BF = 0.39)<br>M2: Sex + Group (BF = 0.02) |
| **Win/Stay** | 140.65<br>(35.12) | 133.51<br>(33.08) | b = -8.99;<br>*t*(263) = -2.21 | **0.03** | 0.02 | **Best Model (M1): Age + Group + Sex (BF > 1000)**<br>M2: Age + Sex (BF > 1000)<br>M1/M2: BF = 1.31 |
| **Win/Shift** | 30.97<br>(30.88) | 34.79<br>(28.6) | b = 5.57;<br>*t*(263) = 1.58 | 0.11 | 0.01 | Best Model (M1): Age + Sex (BF > 1000)<br>M2: Age + Sex + Group (BF > 1000)<br>M1/M2: BF = 2.28 |
| **Lose/Stay** | 47.27<br>(27.07) | 50.48<br>(29.67) | b = 2.04;<br>*t*(263) = 0.58 | 0.57 | 0.00 | Best Model (M1): Age (BF = 565.78)<br>M2: Age + Group (BF = 14.67) |
| **Lose/Shift** | 81.11<br>(28.11) | 81.21<br>(31.75) | b = 1.38;<br>*t*(263) = 0.37 | 0.71 | 0.00 | Best Model (M1): Age (BF = 478.04)<br>M2: Age + Group (BF = 13.73) |

**Note:** Inferential statistics reported here are based on linear models including age and sex as covariates. For analogous results in the subsample with available WRAT scores (including WRAT as an additional covariate), see **Table S1.5**. For results dividing choices into early and late choices per block, see **Table S1.6**.
\*See note below **Table 6** for interpretation of these Bayes factor analyses. For results in which BFs for both M1 and M2 are >1000 relative to an intercept-only model, we also report the BF for M1 relative to M2 for comparative interpretability.

In most cases, similar results were seen in the subsample with WRAT scores (see **Tables S1.6-8**). The major exception was that this subsample did find higher action precision in HCs than SUDs, similar to our initial study. Evidence for the difference in learning rates for losses was also weaker in this subsample, but it was retained within the winning model in model comparison (BF = 2.93).

### 3.4 Relationships with symptoms

Relationships between model parameters and symptom measures in SUDs are reported in **Figure S1.2**. No significant relationships were observed, which failed to replicate the relationships observed in our previous study between PHQ/OASIS and information insensitivity, and between OASIS and reward sensitivity (BFs also provided moderate evidence against these relationships: 0.18 – 0.53).

When comparing the main sample of SUDs to the 32 diagnosed individuals with symptom severity below the cutoff of DAST ≥ 3, we found no differences in parameter values (see **Table S1.4**).

Comparison of those that were vs. were not on medication in the SUDs group revealed no significant differences for any model parameter (*t*s between −1.32 and .86, *p*s between .188 and .838).

### 3.5 Analysis of specific use disorders in combined samples

Summary statistics for computational parameters in specific disorder groups within the combined exploratory and confirmatory samples (N = 468) are shown in **Table S1.9**. Full results of the logistic regression models differentiating whether participants had each specific disorder relative to HCs are shown in **Table S1.10**. Accounting for other model parameters, these analyses found that action precision separately differentiated diagnostic status relative to HCs for those with each specific substance use and affective disorder (Wald *z* between −4.63 and −2.03, *p*s between < .001 and .042), with the exception of hallucinogen use disorder and PTSD. This was also true for those with stimulant use disorder without co-morbidities (z = −3.05, *p* = .002). Learning rates for wins separately predicted diagnostic status relative to HCs for those with stimulant use disorder (only marginally in those without co-morbidities; *p* = .079), opioid use disorder, sedative use disorder, major depressive disorder, and social anxiety disorder (*z* between −3.2 and −1.96, *p*s between .001 and .049). Learning rates for losses further predicted diagnostic status relative to HCs for those with the majority of specific disorders (including stimulant use disorders without co-morbidities; *z* between −3.57 and −2.08, *p*s between < .001 and .038), with the exception of hallucinogen use disorder, generalized anxiety disorder, panic disorder, or PTSD.

Full results of the logistic regression models predicting whether participants in the SUDs group had a specific disorder relative to other disorders are shown in **Table S1.11**. Accounting for other model parameters, these analyses found that greater action precision and faster learning rate for wins predicted diagnosis with stimulant use disorders vs. other disorders (*z* = 2.47 and 2.23, *p* = .014 and .026, respectively; and a marginal effect of learning rates for losses: *z* = 1.79, *p* = .073). Diagnosis with social anxiety disorder was also predicted by slower learning rates for wins relative to other disorders (*z* = −1.97, *p* = .049; and a marginal effect of learning rates for losses: *z* = −1.71, *p* = .09). Otherwise, model parameters did not differentiate between specific SUDs. Aside from social anxiety and learning rate for wins, they also did not indicate differences in SUDs with vs. without affective disorders for any model parameter.

The overall variance explained by the 5 parameters in significant logistic regression models ranged from 9% to 16% (based on pseudo-R^2^; for details, see **Table S1.10**). **Figure 4** shows parameter values for those with each specific disorder and indicates which could be differentiated with the logistic regressions.

**Figure 4.**
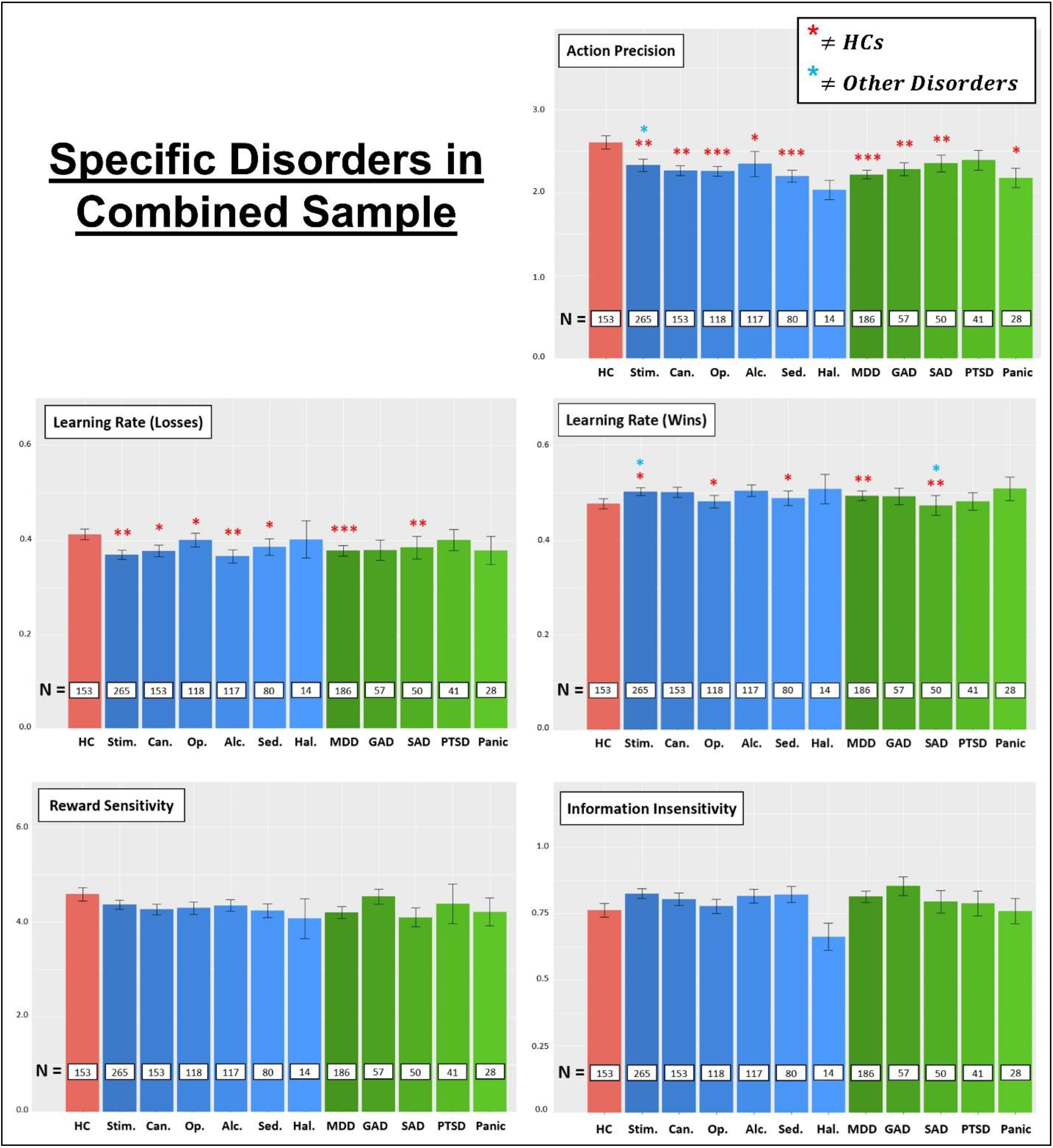
Comparison of HCs and subsets of individuals with specific SUDs/affective disorders within the combined exploratory and confirmatory samples (sample size per group is indicated within each bar; based on the groupings detailed in **Table 2**). Red stars indicate that, in logistic regressions, model parameters could predict whether individuals were HCs or had a specific disorder (i.e., removing individuals from analyses without the disorder in question; *p<.05, **p<.01, ***p<.001). Blue stars indicate that model parameters could further predict whether an individual had one disorder relative to other disorders (i.e., removing HCs from analyses). Statistical results are reported in **Tables S1.10-S1.11**. For a similar plot of general task performance (number of wins) by specific disorder group, see **Figure S1.3**. Legend: Stim. Only = stimulant use disorders without co-morbidities, Can. = cannabis use disorders, Op. = opioid use disorders, Alc. = alcohol use disorders, Sed. = sedative use disorders, Hal. = hallucinogen use disorders, SAD = social anxiety disorder, PTSD = post-traumatic stress disorders.

### 3.6 Predictive categorization

Results of predictive categorization analyses (ROCs, AUCs, accuracy, and confusion matrices) for specific disorders are shown in **Figure 5**. When tested on the confirmatory dataset, logistic regression models trained on the exploratory dataset varied in predictive classification accuracy (relative to HCs) between .55 and .66, depending on the specific disorder in question. AUCs ranged from .6 to .7, indicating poor to acceptable discrimination. When categorizing HCs vs. all individuals with SUDs together, accuracy = .63 and AUC = .66.

**Figure 5.**
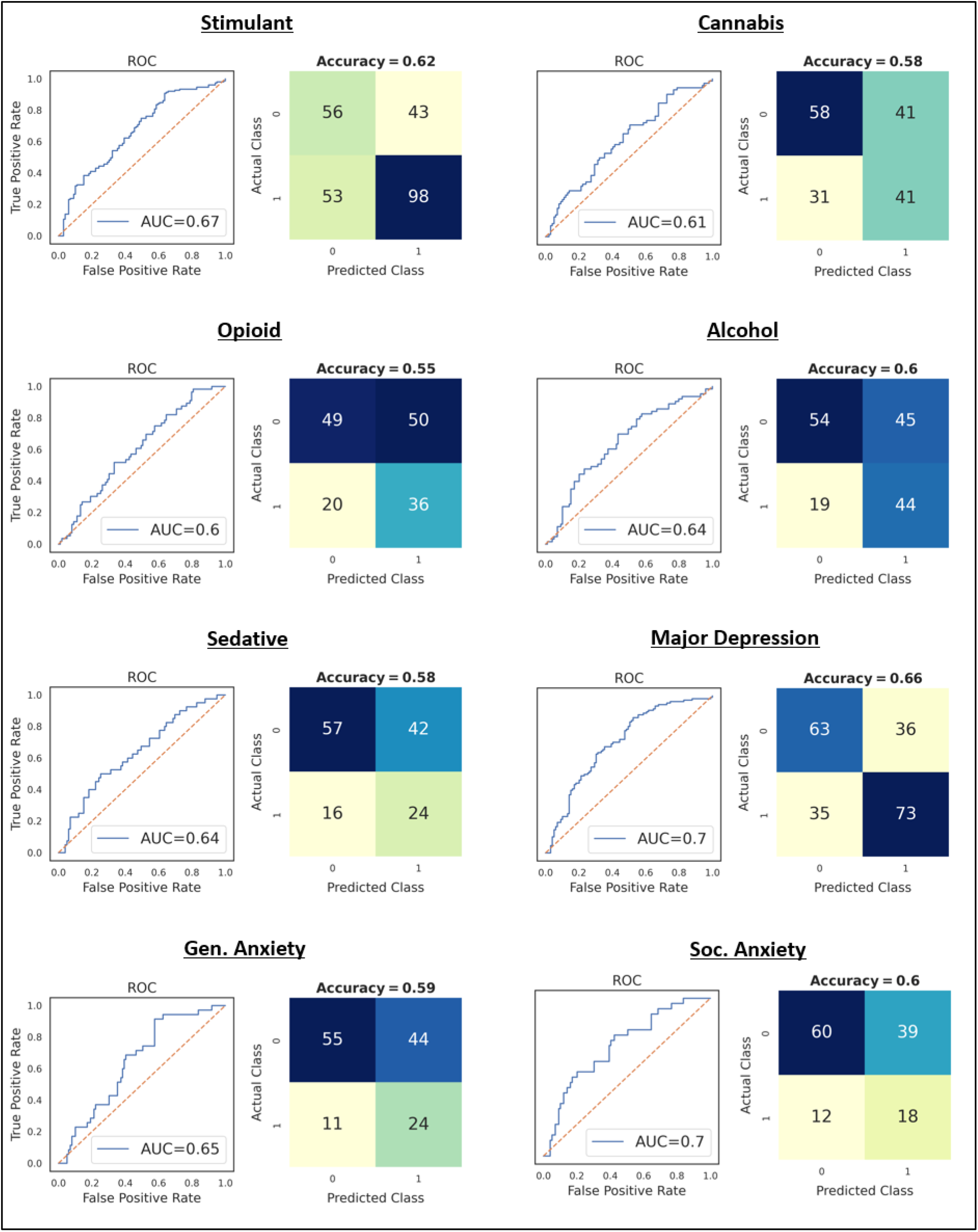
Results of logistic regression models trained on the exploratory sample and then tested on the confirmatory sample (classifying individuals as HCs vs. having a specific SUD or affective disorder). Receiver operating characteristic curves (ROCs) illustrate the true positive rate (sensitivity) vs. false positive rate (1 - specificity) for different categorization thresholds. Performance is quantified by associated area-under-the-curve (AUC) scores, reflecting how often a random sample will be assigned to the correct group with higher probability. Acceptable AUCs vary by application, but the following heuristic cutoff values have been proposed: 0.5 = No discrimination, 0.5-0.7 = Poor discrimination, 0.7-0.8 = Acceptable discrimination, 0.8-0.9= Excellent discrimination, and > 0.9 = Outstanding discrimination (37). Accuracy at a neutral threshold of 0.5 is also shown. Confusion matrices for each disorder (0 = HCs, 1 = Disorder) illustrate the numbers of accurate and inaccurate classifications for each group based on this criterion. Note that training data were balanced using frequency weights prior to training. Gen. Anxiety = Generalized Anxiety Disorder; Soc. Anxiety = Social Anxiety Disorder.

## 4. DISCUSSION

In this study, we found confirmatory evidence that substance users show selectively slower learning rates than HCs for negative outcomes. In contrast, previously observed differences in action precision and learning rates for positive outcomes did not replicate for most analyses in this new sample (with the exception of action precision in supplementary frequentist analyses accounting for WRAT scores). Also notable was the reduced reward sensitivity (suggesting greater directed exploration) observed within SUDs, which was not observed in our prior study.

Subsequent logistic regressions across participants in the exploratory and confirmatory samples showed that both action precision and learning rates for losses (and in some cases learning rates for wins) could jointly differentiate the presence of most specific substance use and affective disorders relative to HCs. This highlights the way a multi-dimensional computational phenotype can offer categorical information not available from summary statistics or a single parameter alone. In contrast, analogous logistic regressions found that most disorders could not be differentiated from one another, except for stimulant use disorders (which showed greater action precision and faster learning rates for wins relative to other disorders) and social anxiety disorder (slower learning rates for wins relative to other disorders). When considering co-morbid affective disorders, this general lack of differentiability further indicated, for example, that learning rates for losses and action precision did not differ in SUDs with vs. without MDD or anxiety disorders.

When logistic regressions were instead trained on the exploratory sample and tested on their ability to correctly categorize HCs vs. specific disorders in the confirmatory sample, performance was above chance but ranged from poor to acceptable discrimination across specific disorders. Thus, while the group differences we observed may offer important mechanistic insights, their potential clinical utility for diagnostic purposes should not be overstated.

Overall, these results provide strong evidence that the differences we observed in SUDs are transdiagnostic. They also provide added support for the hypothesis that individuals with SUDs adjust their behavior more slowly in the face of aversive outcomes, which could help explain continued drug use despite harmful effects. This is also supported by previous literature showing reduced sensitivity to negative outcomes (or to affective stimuli generally) in this population within other contexts (e.g., (18, 38–49)). Given this replication, there are now several future directions that should be considered. One important question, for example, is whether this difference is caused by substance use or whether it represents a pre-existing vulnerability factor. Answering this question will likely require longitudinal analyses of at-risk populations, as well as assessing potential relationships with pre-existing biological and psychological risk-factors. Another important topic to address will be whether this difference is affected by treatment and covaries with symptom severity. Randomized controlled trials assessing the effects of distinct treatments on learning rates will be necessary to shed light on this question. Additionally, neuroimaging studies designed to identify the neural correlates of these learning rate differences could highlight potential biological targets. Finally, it will be important to identify whether learning rates moderate treatment outcomes. In relation to this last point, we recently reported a 1-year follow-up study in our previous sample and unexpectedly found that slower learning rates for losses at baseline predicted greater symptom improvement in stimulant users at 1-year follow-up (20). Here we suggested that, despite potentially reducing learning from the harmful outcomes of drug use, this difference might also attenuate reactions to unpleasant aspects of the recovery process and facilitate adherence. However, as it was not an expected result, we aim to replicate this longitudinal effect in future work. If confirmed, it could indicate one concrete way in which measures of learning rate differences could be clinically informative.

Some remaining limitations are important to consider. First, our sample of SUDs was a heterogenous community sample from a particular region of the United States, the great majority of which were stimulant users, which may have limited our ability to detect effects specific to some substances and not others (with the exception of a moderate number of stimulant users that did not have co-morbidities in the combined sample), or to generalize findings to substance users in other regional or cultural contexts. It was also a treatment-seeking, abstinent sample, and thus may not generalize to non-treatment-seeking individuals. Many substance users also had affective disorders. However, parameters did not correlate with depression/anxiety symptoms and, with the exception of social anxiety disorder (slower learning rate for wins), model parameters in the combined sample could not differentiate SUDs with vs. without affective disorders. Finally, the correct interpretation is less clear for results that were significant in the previous or current sample, but not both (e.g., with respect to reduced action precision and reward sensitivity in SUDs). Future studies will therefore need to examine whether inconsistent results between samples are better understood as false positives in one sample or false negatives in the other.

In summary, this study lends added confidence to the generalizability of previous results suggesting slower learning from negative outcomes in SUDs and motivates a number of future directions to test the neurobiological basis of this difference and whether it might represent, for example, a pre-existing vulnerability factor, a predictor of treatment response, an objective severity marker, and/or a novel intervention target. These represent important next steps toward evaluating the clinical utility of this potentially important individual difference.

**Software Note**: All model simulations, model comparison, and parametric empirical Bayes analyses were implemented using standard routines (**spm_MDP_VB_X.m**, **spm_BMS.m**, **spm_dcm_peb.m**, **spm_dcm_peb_bmc.m**) that are available as MATLAB code in the latest version of SPM academic software: http://www.fil.ion.ucl.ac.uk/spm/. For the specific code used to build the three-armed bandit task model and fit parameters to data, see: https://github.com/rssmith33/3-armed_bandit_task_model.

## Data Availability

All data produced in the present study are available upon reasonable request to the authors

## Contributors

Samuel Taylor performed analyses and wrote the initial draft of the manuscript. Claire A. Lavalley and Navid Hakimi performed analyses and edited the manuscript. Jennifer L. Stewart, Maria Ironside, Haixia Zheng, Evan White, Salvador Guinjoan, and Martin P. Paulus reviewed and edited the manuscript. Ryan Smith performed analyses, reviewed and edited the manuscript, and supervised the project. All authors have approved the final article.

## Acknowledgment

This study was supported by the Laureate Institute for Brain Research.

## Supplementary Materials 1

**Table S1.1:** Demographic Differences in Parameters for Participants between Exploratory and Confirmatory Samples

|  | Exploratory | Confirmatory | Test | p-value | BF |
| --- | --- | --- | --- | --- | --- |
| **Healthy Controls** |  |  |  |  |  |
| **N =** | 54 | 99 |  |  |  |
| **Age** | 32.27 (11.35) | 32.29 (11.08) | $t(151) = -0.01$;<br>$d = 0.0$ | 0.99 | 0.18 |
| **Sex (Male)** | 24 (44%) | 39 (39.4%) | $\chi^2(1) = 0.19$<br>Cramer's $v = 0.04$ | 0.66 | 0.45 |
| **DAST** | 0.11 (0.37) | 0.1 (0.33) | $t(151) = 0.17$;<br>$d = 0.03$ | 0.86 | 0.18 |
| **PHQ** | 0.8 (1.28) | 1.22 (1.94) | $t(151) = -1.45$;<br>$d = 0.24$ | 0.15 | 0.47 |
| **OASIS** | 1.35 (1.94) | 1.19 (1.91) | $t(151) = 0.49$;<br>$d = 0.08$ | 0.62 | 0.2 |
| **WRAT** | 63.53 (4.93) | 62.95 (5.33) | $t(136) = 0.63$;<br>$d = 0.11$ | 0.53 | 0.23 |
| **Substance Use Disorder** |  |  |  |  |  |
| **N =** | 147 | 168 |  |  |  |
| **Age** | 34.05 (9.17) | 33.75 (8.34) | $t(313) = 0.3$;<br>$d = 0.03$ | 0.76 | 0.13 |
| **Sex (Male)** | 72(49%) | 63 (37.5%) | $\chi^2(1) = 3.76$<br>Cramer's $v = 0.11$ | 0.052 | 2.25 |
| **DAST** | 7.54 (2.22) | 7.9 (1.81) | $t(313) = -1.59$;<br>$d = 0.18$ | 0.11 | 0.42 |
| **PHQ** | 6.58 (5.7) | 7.89 (6.58) | $t(312) = -1.88$;<br>$d = 0.21$ | 0.06 | 0.68 |
| **OASIS** | 5.84 (4.63) | 6.36 (4.51) | $t(313) = -1$;<br>$d = 0.11$ | 0.32 | 0.2 |
| **WRAT** | 58.47 (5.85) | 56.67 (6.74) | $t(253) = 2.27$;<br>$d = 0.29$ | **0.02** | 1.56 |

**Table S1.2.**
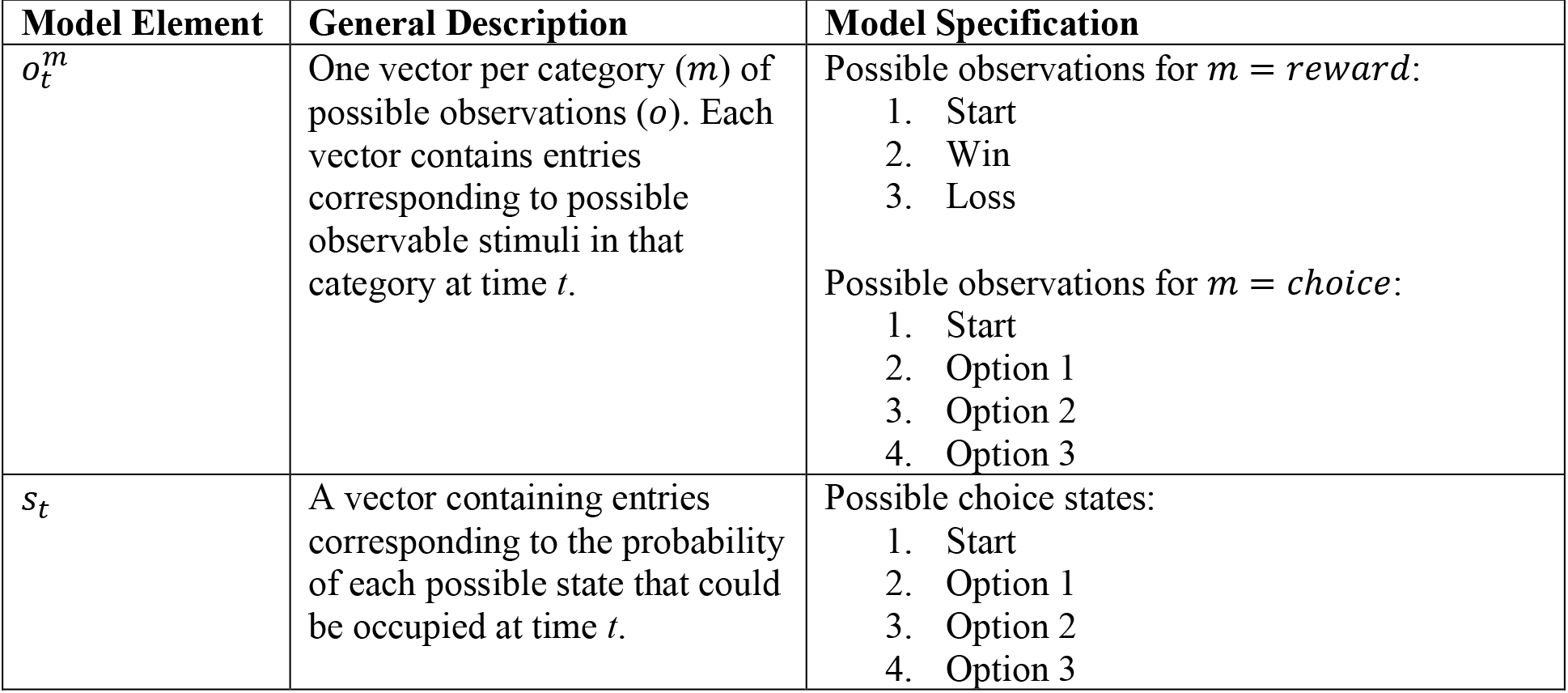
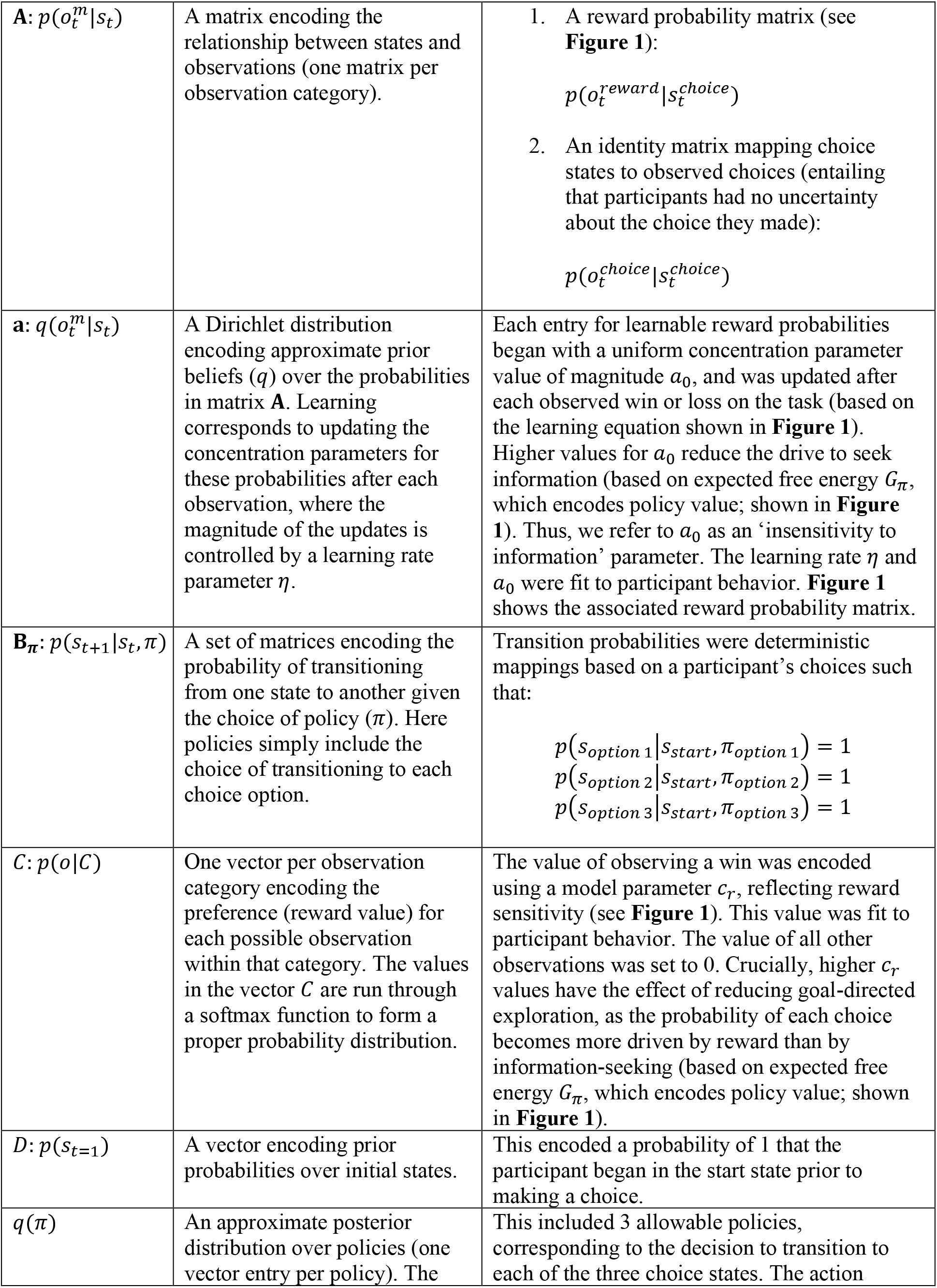
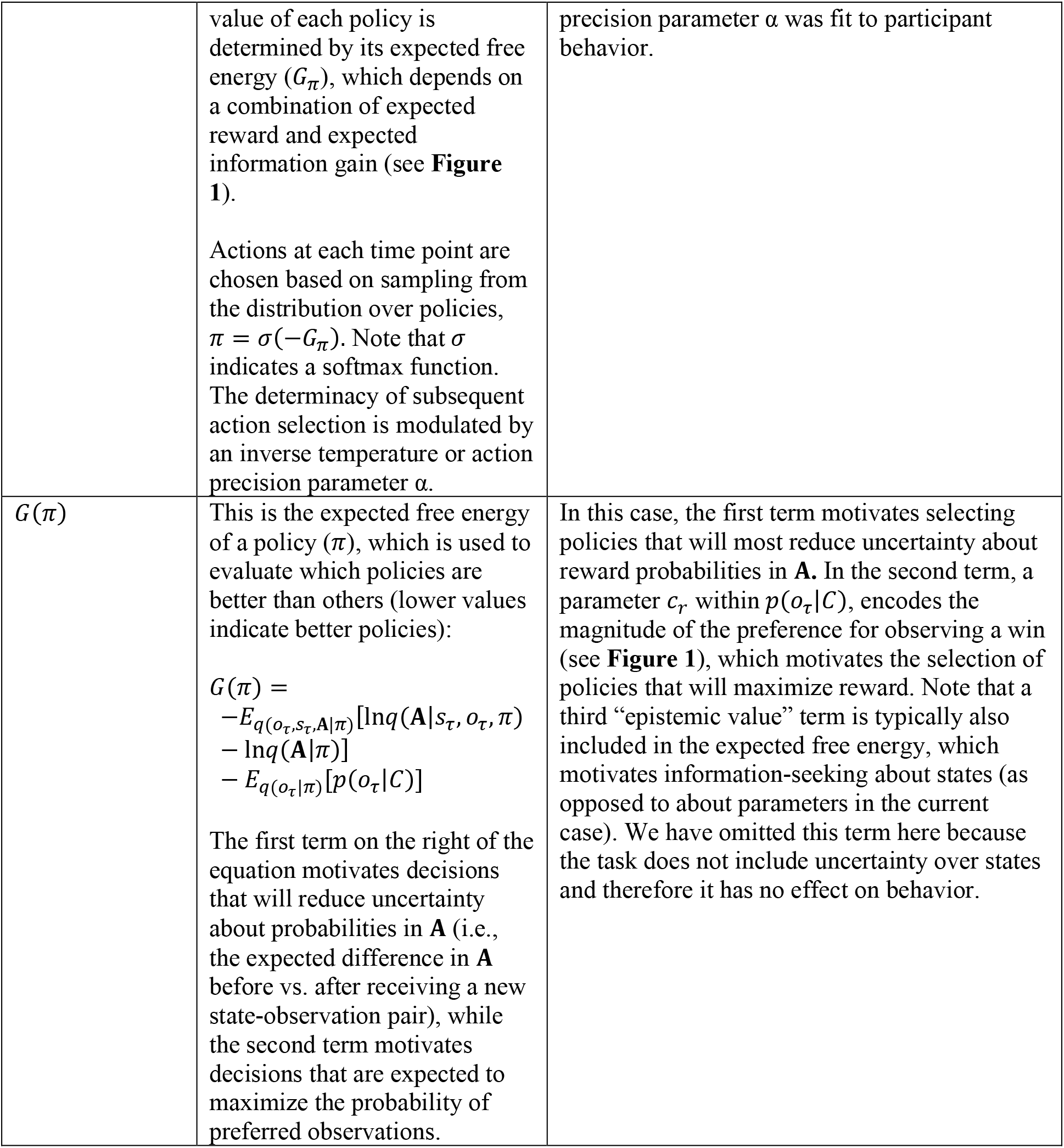
Elements of the generative model of the three-armed bandit task

| Model Element | General Description | Model Specification |
| --- | --- | --- |
| $o_t^m$ | One vector per category ($m$) of possible observations ($o$). Each vector contains entries corresponding to possible observable stimuli in that category at time $t$. | Possible observations for $m = \text{reward}$:<br>1. Start<br>2. Win<br>3. Loss<br>Possible observations for $m = \text{choice}$:<br>1. Start<br>2. Option 1<br>3. Option 2<br>4. Option 3 |
| $s_t$ | A vector containing entries corresponding to the probability of each possible state that could be occupied at time $t$. | Possible choice states:<br>1. Start<br>2. Option 1<br>3. Option 2<br>4. Option 3 |
| $\mathbf{A}: p(o_t^m \vert s_t)$ | A matrix encoding the relationship between states and observations (one matrix per observation category). | 1. A reward probability matrix (see **Figure 1**):<br>$p(o_t^{reward} \vert s_t^{choice})$<br>2. An identity matrix mapping choice states to observed choices (entailing that participants had no uncertainty about the choice they made):<br>$p(o_t^{choice} \vert s_t^{choice})$ |
| $\mathbf{a}: q(o_t^m \vert s_t)$ | A Dirichlet distribution encoding approximate prior beliefs ($q$) over the probabilities in matrix **A**. Learning corresponds to updating the concentration parameters for these probabilities after each observation, where the magnitude of the updates is controlled by a learning rate parameter $\eta$. | Each entry for learnable reward probabilities began with a uniform concentration parameter value of magnitude $a_0$, and was updated after each observed win or loss on the task (based on the learning equation shown in **Figure 1**). Higher values for $a_0$ reduce the drive to seek information (based on expected free energy $G_\pi$, which encodes policy value; shown in **Figure 1**). Thus, we refer to $a_0$ as an ‘insensitivity to information’ parameter. The learning rate $\eta$ and $a_0$ were fit to participant behavior. **Figure 1** shows the associated reward probability matrix. |
| $\mathbf{B}_\pi: p(s_{t+1} \vert s_t, \pi)$ | A set of matrices encoding the probability of transitioning from one state to another given the choice of policy ($\pi$). Here policies simply include the choice of transitioning to each choice option. | Transition probabilities were deterministic mappings based on a participant’s choices such that:<br>$p(s_{option\ 1} \vert s_{start}, \pi_{option\ 1}) = 1$ $p(s_{option\ 2} \vert s_{start}, \pi_{option\ 2}) = 1$ $p(s_{option\ 3} \vert s_{start}, \pi_{option\ 3}) = 1$ |
| $\mathbf{C}: p(o \vert C)$ | One vector per observation category encoding the preference (reward value) for each possible observation within that category. The values in the vector $\mathbf{C}$ are run through a softmax function to form a proper probability distribution. | The value of observing a win was encoded using a model parameter $c_r$, reflecting reward sensitivity (see **Figure 1**). This value was fit to participant behavior. The value of all other observations was set to 0. Crucially, higher $c_r$ values have the effect of reducing goal-directed exploration, as the probability of each choice becomes more driven by reward than by information-seeking (based on expected free energy $G_\pi$, which encodes policy value; shown in **Figure 1**). |
| $\mathbf{D}: p(s_{t=1})$ | A vector encoding prior probabilities over initial states. | This encoded a probability of 1 that the participant began in the start state prior to making a choice. |
| $q(\pi)$ | An approximate posterior distribution over policies (one vector entry per policy). The | This included 3 allowable policies, corresponding to the decision to transition to each of the three choice states. The action |
|  | value of each policy is determined by its expected free energy ($G_\pi$), which depends on a combination of expected reward and expected information gain (see **Figure 1**).<br>Actions at each time point are chosen based on sampling from the distribution over policies, $\pi = \sigma(-G_\pi)$. Note that $\sigma$ indicates a softmax function. The determinacy of subsequent action selection is modulated by an inverse temperature or action precision parameter $\alpha$. | precision parameter $\alpha$ was fit to participant behavior. |
| $G(\pi)$ | This is the expected free energy of a policy ($\pi$), which is used to evaluate which policies are better than others (lower values indicate better policies):<br>$G(\pi) = -E_{q(o_\tau, s_\tau, \mathbf{A}\vert \pi)}[\ln q(\mathbf{A}\vert s_\tau, o_\tau, \pi)] - \ln q(\mathbf{A}\vert \pi) - E_{q(o_\tau\vert \pi)}[p(o_\tau\vert C)]$<br>The first term on the right of the equation motivates decisions that will reduce uncertainty about probabilities in **A** (i.e., the expected difference in **A** before vs. after receiving a new state-observation pair), while the second term motivates decisions that are expected to maximize the probability of preferred observations. | In this case, the first term motivates selecting policies that will most reduce uncertainty about reward probabilities in **A**. In the second term, a parameter $c_r$, within $p(o_\tau\vert C)$, encodes the magnitude of the preference for observing a win (see **Figure 1**), which motivates the selection of policies that will maximize reward. Note that a third “epistemic value” term is typically also included in the expected free energy, which motivates information-seeking about states (as opposed to about parameters in the current case). We have omitted this term here because the task does not include uncertainty over states and therefore it has no effect on behavior. |

**Table S1.3:**
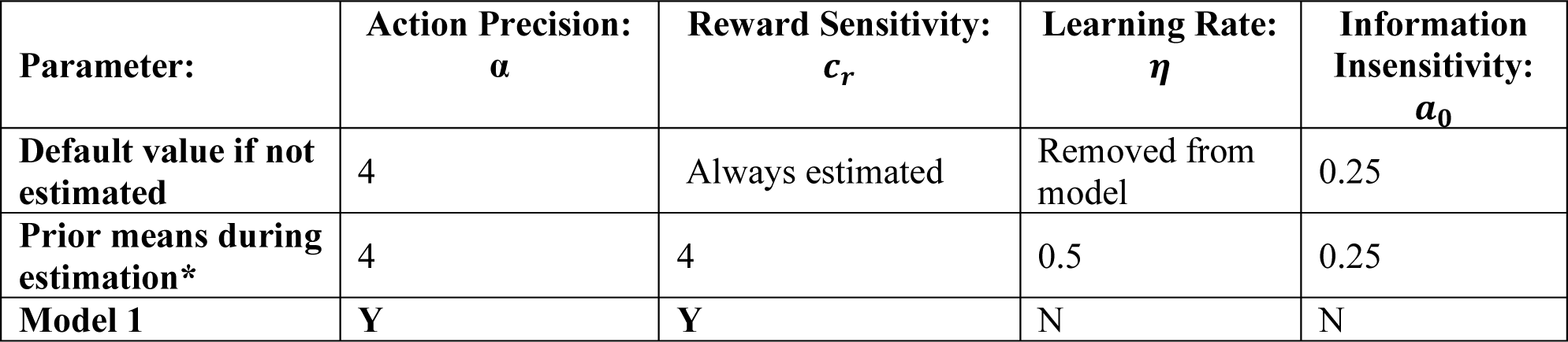
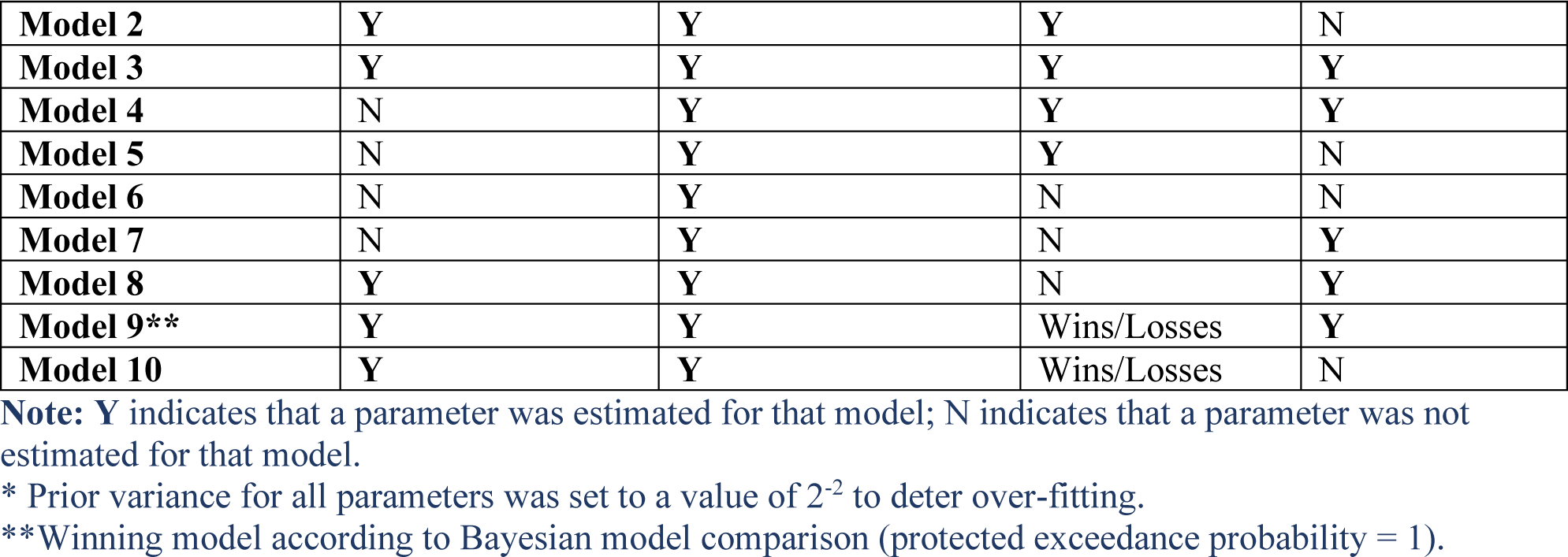
Nested Models

**Table S1.4:** Differences Between SUDs with vs. without DAST Scores Less than 3

| | SUDs<br>(DAST $\geq 3$) | SUDs<br>(DAST $< 3$) | *t*-value &<br>Cohen's *d* | *p*-value | BF |
| --- | --- | --- | --- | --- | --- |
| **Demographic Information** |  |  |  |  |  |
| **N =** | 168 | 32 |  |  |  |
| **Age** | 33.75 (8.34) | 34.51<br>(11.07) | $t(198) = -0.44$;<br>$d = 0.06$ | 0.66 | 0.22 |
| **Sex (Male)** | 0.38 (0.49) | 0.44 (0.5) | $t(198) = -0.66$;<br>$d = 0.09$ | 0.51 | 0.25 |
| **DAST** | 7.9 (1.81) | 0.75 (0.76) | $t(198) = 21.9$;<br>$d = 3.11$ | **0.001** | **> 1000** |
| **PHQ** | 7.89 (6.58) | 13.31 (5.24) | $t(198) = -4.4$;<br>$d = 0.63$ | **< 0.001** | **> 1000** |
| **OASIS** | 6.36 (4.51) | 9.81 (3.28) | $t(198) = -4.13$;<br>$d = 0.59$ | **< 0.001** | **372.01** |
| **WRAT** | 56.67 (6.74) | 62.48 (5.13) | $t(149) = -4.47$;<br>$d = 0.73$ | **< 0.001** | **> 1000** |
| **Parameters** |  |  |  |  |  |
| **Action Precision** | 2.38 (1.48) | 2.31 (0.75) | $t(198) = 0.25$;<br>$d = 0.04$ | 0.8 | 0.21 |
| **Reward Sensitivity** | 4.43 (1.68) | 4.66 (1.43) | $t(198) = -0.73$;<br>$d = 0.1$ | 0.47 | 0.26 |
| **Learning Rate (Wins)** | 0.5 (0.15) | 0.48 (0.14) | $t(198) = 0.4$;<br>$d = 0.06$ | 0.69 | 0.22 |
| **Learning Rate (Losses)** | 0.37 (0.16) | 0.41 (0.15) | $t(198) = -1.54$;<br>$d = 0.22$ | 0.12 | 0.59 |
| **Information Insensitivity** | 0.83 (0.29) | 0.77 (0.34) | $t(198) = 1.02$;<br>$d = 0.14$ | 0.31 | 0.32 |

**Table S1.5:**
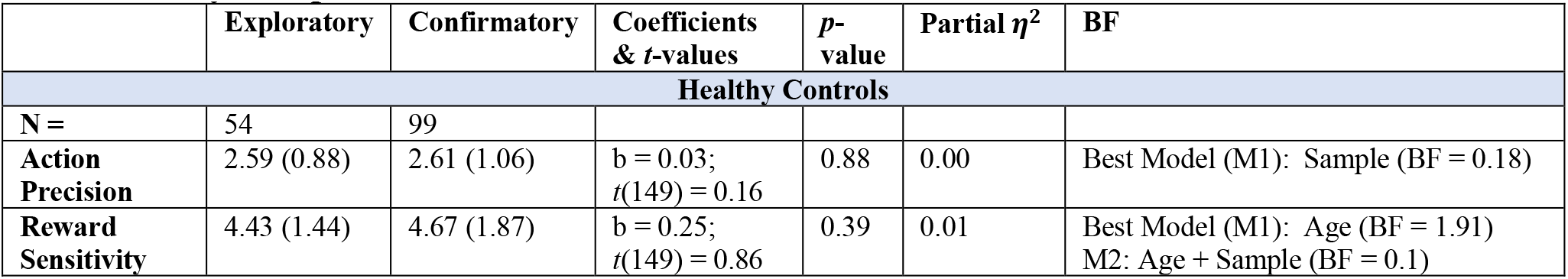
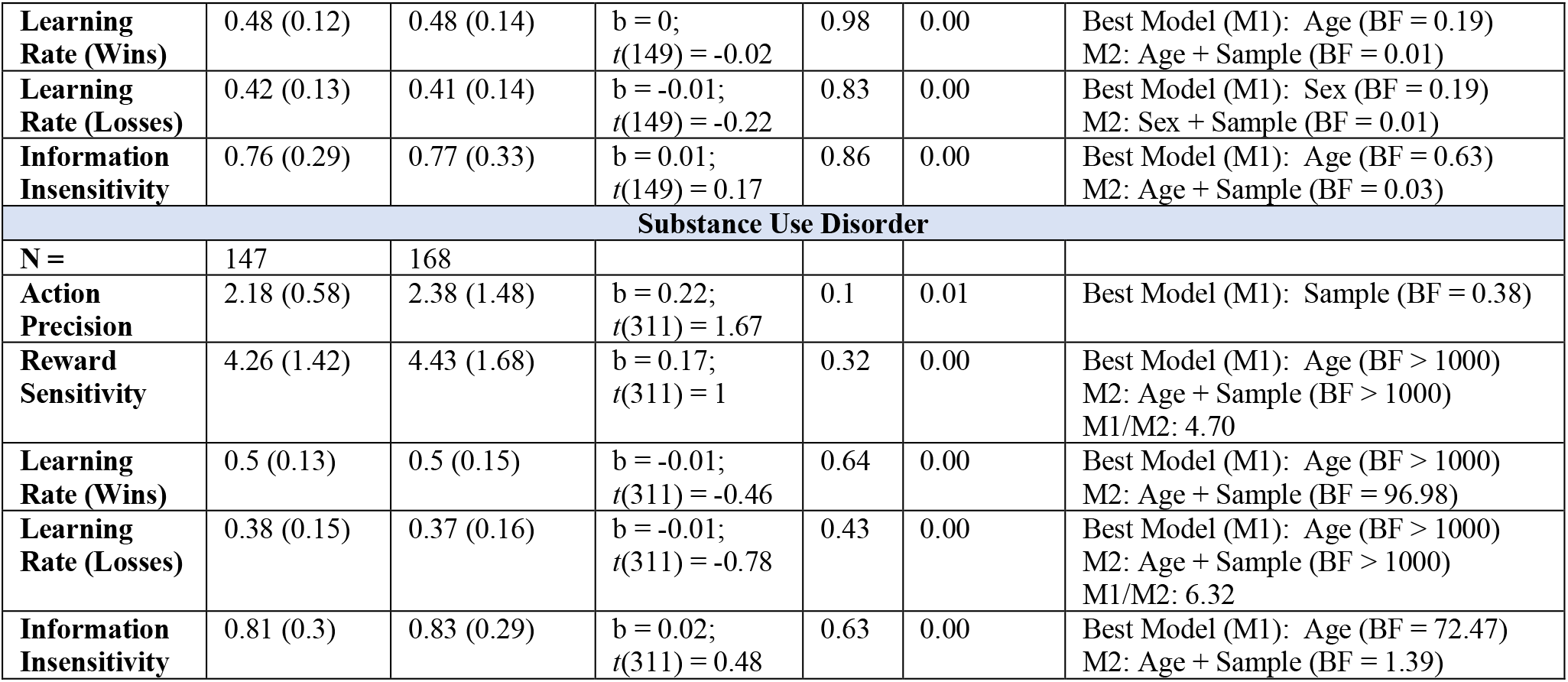
Differences in Parameters for Participants between Exploratory and Confirmatory Samples

| | Exploratory | Confirmatory | Coefficients<br>& *t*-values | *p*-<br>value | Partial $\eta^2$ | BF |
| --- | --- | --- | --- | --- | --- | --- |
| **Healthy Controls** |  |  |  |  |  |  |
| **N =** | 54 | 99 |  |  |  |  |
| **Action Precision** | 2.59 (0.88) | 2.61 (1.06) | $b = 0.03$;<br>$t(149) = 0.16$ | 0.88 | 0.00 | Best Model (M1): Sample (BF = 0.18) |
| **Reward Sensitivity** | 4.43 (1.44) | 4.67 (1.87) | $b = 0.25$;<br>$t(149) = 0.86$ | 0.39 | 0.01 | Best Model (M1): Age (BF = 1.91)<br>M2: Age + Sample (BF = 0.1) |
| **Learning Rate (Wins)** | 0.48 (0.12) | 0.48 (0.14) | $b = 0;$<br>$t(149) = -0.02$ | 0.98 | 0.00 | Best Model (M1): Age (BF = 0.19)<br>M2: Age + Sample (BF = 0.01) |
| **Learning Rate (Losses)** | 0.42 (0.13) | 0.41 (0.14) | $b = -0.01;$<br>$t(149) = -0.22$ | 0.83 | 0.00 | Best Model (M1): Sex (BF = 0.19)<br>M2: Sex + Sample (BF = 0.01) |
| **Information Insensitivity** | 0.76 (0.29) | 0.77 (0.33) | $b = 0.01;$<br>$t(149) = 0.17$ | 0.86 | 0.00 | Best Model (M1): Age (BF = 0.63)<br>M2: Age + Sample (BF = 0.03) |
| **Substance Use Disorder** |  |  |  |  |  |  |
| **N =** | 147 | 168 |  |  |  |  |
| **Action Precision** | 2.18 (0.58) | 2.38 (1.48) | $b = 0.22;$<br>$t(311) = 1.67$ | 0.1 | 0.01 | Best Model (M1): Sample (BF = 0.38) |
| **Reward Sensitivity** | 4.26 (1.42) | 4.43 (1.68) | $b = 0.17;$<br>$t(311) = 1$ | 0.32 | 0.00 | Best Model (M1): Age (BF > 1000)<br>M2: Age + Sample (BF > 1000)<br>M1/M2: 4.70 |
| **Learning Rate (Wins)** | 0.5 (0.13) | 0.5 (0.15) | $b = -0.01;$<br>$t(311) = -0.46$ | 0.64 | 0.00 | Best Model (M1): Age (BF > 1000)<br>M2: Age + Sample (BF = 96.98) |
| **Learning Rate (Losses)** | 0.38 (0.15) | 0.37 (0.16) | $b = -0.01;$<br>$t(311) = -0.78$ | 0.43 | 0.00 | Best Model (M1): Age (BF > 1000)<br>M2: Age + Sample (BF > 1000)<br>M1/M2: 6.32 |
| **Information Insensitivity** | 0.81 (0.3) | 0.83 (0.29) | $b = 0.02;$<br>$t(311) = 0.48$ | 0.63 | 0.00 | Best Model (M1): Age (BF = 72.47)<br>M2: Age + Sample (BF = 1.39) |

### Additional Effects

PEB models in the full sample revealed strong evidence for a negative association between age and action precision and a positive association between age and reward sensitivity (*pp* = 1 in both cases). There was also strong evidence for a positive relationship between age and learning rate for wins (*pp* = 0.93). Including WRAT reading scores (in the subsample for which they were available) did not change these results (see **Figure S1.1**), except the association between age and learning rate for wins was not observed.

As in the PEB analyses, some effects of age were also observed in frequentist regressions. In the full sample, there was a positive association between age and reward sensitivity (t(263) = 4.29, p < .001), learning rates for wins (t(263) = 2.91, p = .004), and information insensitivity (t(263) = 2.38, p = .02), and a negative association between age and learning rates for losses (t(263) = 2.82, p = .005). For complete statistical results associated with all effects, see **Table S2.3**.

A similar pattern of results was also observed in the subsample with available WRAT scores for reward sensitivity, learning rates for losses, and information insensitivity (see **Tables S1.6 and S2.4**). Unlike in the full sample, analyses in the subsample with WRAT scores found a significant difference in action precision (*p* < .01), similar to that observed in our prior study. The group difference in learning rate for losses was not significant (*p* = .11) in this subsample. However, it was retained in the best model in the associated Bayes factor analyses (BF = 2.93).

Results of descriptive behavioral measures are reported in **Table S1.7** for those with WRAT scores. Here found fewer win/stay choices in SUDs, which was not observed previously. As in our prior study, we also repeated these analyses for the first and second halves of each game separately (i.e., first 7 choices vs. subsequent choices) to assess periods where exploration vs. exploitation would be expected to dominate, respectively (see **Table S1.8**). The subsample including WRAT scores showed group differences in win/stay (greater in HCs) and win/shift (greater in SUDs) choices during both early and late choice. Unlike in the full sample, number of wins did not differ between groups.

In the full sample, there were positive associations between age and both win/stay (*t*(263) = 5.48, *p* < .001) and lose/stay (*t*(263) = 4.22, *p* < .001) choices, and negative associations between age and both win/shift (*t*(263) = 6.01, *p* < .001) and lose/shift (*t*(263) = 4.29, *p* < .001) choices. There were also effects of sex on both win/stay (greater in females; *t*(263) = 2.21, *p* = .03) and win/shift (greater in males; *t*(263) = 2.20, *p* = .03) choices, but no effects on lose/stay or lose/shift choices. The same pattern of results was also seen when including WRAT scores in the subset of participants in which they were available; WRAT scores were not associated with any variable (see **Table S2.3 and S2.4**).

**Table S1.6:** Group Differences in Parameters for Participants with WRAT Scores

| Parameter | HCs | SUDs | Coefficients & *t*-values | *p*-value | Partial $\eta^2$ | BF* |
| --- | --- | --- | --- | --- | --- | --- |
| N = | 87 | 120 |  |  |  |  |
| Action Precision | 2.62<br>(1.1) | 2.19<br>(0.65) | $b = 0.36$;<br>$t(202) = -2.54$ | **0.01** | 0.03 | **Best Model (M1): Group (BF = 39.76)** |
| Reward Sensitivity | 4.56<br>(1.87) | 4.33<br>(1.35) | $b = 0.37$;<br>$t(202) = -1.5$ | 0.14 | 0.01 | Best Model (M1): Age (BF = 76.72)<br>M2: Age + Group (BF = 1.65) |
| Learning Rate (Wins) | 0.47<br>(0.14) | 0.5<br>(0.14) | $b = 0.02$;<br>$t(202) = 0.9$ | 0.37 | 0.00 | Best Model (M1): Age (BF = 0.62)<br>M2: Age + Group (BF = 0.01) |
| Learning Rate (Losses) | 0.42<br>(0.13) | 0.36<br>(0.16) | $b = 0.04$;<br>$t(202) = -1.62$ | 0.11 | 0.01 | **Best Model (M1): Age + Group (BF = 2.93)**<br>M2: Age (BF = 1.83) |
| Information Insensitivity | 0.77<br>(0.35) | 0.82<br>(0.28) | $b = 0.03$;<br>$t(202) = 0.57$ | 0.57 | 0.00 | Best Model (M1): Age (BF = 1.75)<br>M2: Age + Group (BF = 0.02) |

\*These Bayes factors reflect comparisons of evidence for models that did or did not include each possible combination of predictors (i.e., main effects of age, sex, group, and/or WRAT) in analogous Bayesian regressions. If the winning model included a main effect of group, the reported Bayes factor is relative to an intercept-only model, alongside the analogous Bayes factor of a model without the main effect of group (i.e., indicating the relative contribution of group effects in the model). If the winning model did not include group, this winning model and its Bayes factor are reported (relative to an intercept-only model), along with the Bayes factor for a model that added an effect of group. Bolded BF values indicate that the winning model included group as a main effect.

**Table S1.7:** Descriptive Behavioral Measures for Participants with WRAT Scores

| Measure | HCs | SUDs | Coefficients & *t*-values | *p*-value | Partial $\eta^2$ | BF* |
| --- | --- | --- | --- | --- | --- | --- |
| N = | 87 | 120 |  |  |  |  |
| Wins | 182.44<br>(12.93) | 180.37<br>(10.64) | $b = 2.37$;<br>$t(202) = -1.25$ | 0.21 | 0.01 | Best Model (M1): Group (BF = 0.32) |
| Mean RT | 0.57<br>(0.2) | 0.55<br>(0.27) | $b = 0.04$;<br>$t(202) = -0.92$ | 0.36 | 0.00 | Best Model (M1): Age (BF = 0.4)<br>M2: Age + Group (BF < .001) |
| Win/Stay | 137.69<br>(35.34) | 132.1<br>(31.54) | $b = 12.94$;<br>$t(202) = -2.56$ | **0.01** | 0.03 | **Best Model (M1): Age + Group + Sex (BF > 1000)**<br>M2: Age + Sex (BF = 871.33) |
| Win/Shift | 33.11<br>(31.82) | 36.17<br>(27.24) | $b = 10.32$;<br>$t(202) = 2.35$ | **0.02** | 0.03 | Best Model (M1): Age (BF > 1000)<br>M2: Age + Group (BF > 1000)<br>M1/M2: BF = 2.82 |
| Lose/Stay | 46.66<br>(26.05) | 51.48<br>(29.01) | $b = 0.63$;<br>$t(202) = 0.14$ | 0.89 | 0.00 | Best Model (M1): Age (BF = 12.67)<br>M2: Age + Group (BF = 0.33) |
| Lose/Shift | 82.54<br>(26.19) | 80.25<br>(30.9) | $b = 1.99$;<br>$t(202) = 0.43$ | 0.67 | 0.00 | Best Model (M1): Age (BF = 9.12)<br>M2: Age + Group (BF = 0.2) |

\*See note below **Table S.3** for interpretation of these Bayes factor analyses.

**Table S1.8:**
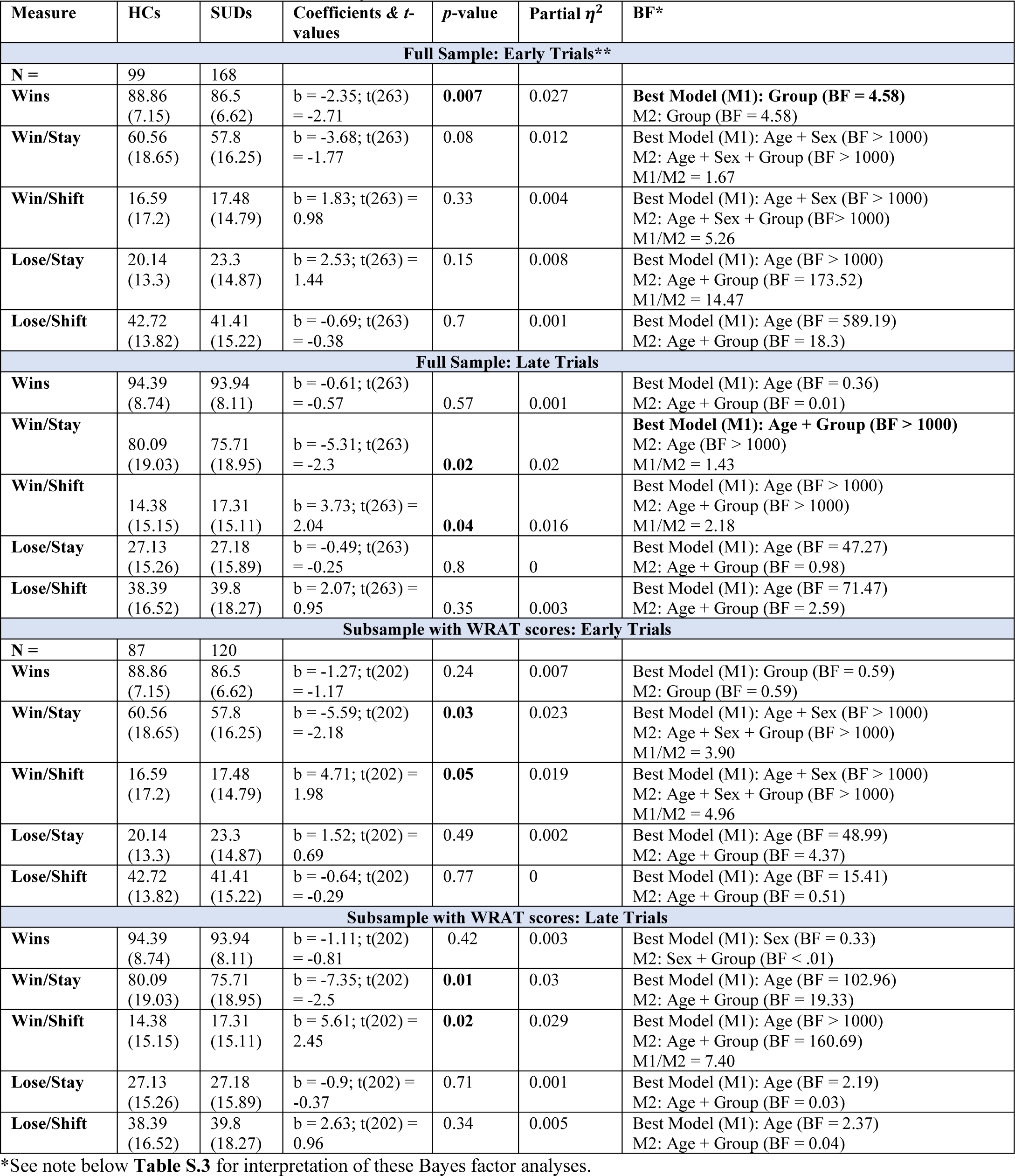
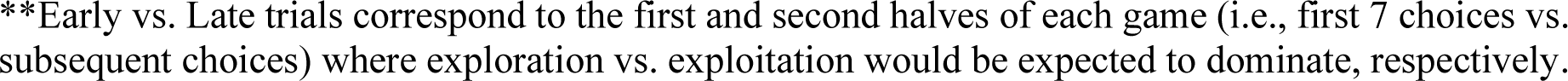
Choice Behavior in Early vs. Late Trials

| Measure | HCs | SUDs | Coefficients & *t*-values | *p*-value | Partial $\eta^2$ | BF* |
| --- | --- | --- | --- | --- | --- | --- |
| **Full Sample: Early Trials\*\*** |  |  |  |  |  |  |
| **N =** | 99 | 168 |  |  |  |  |
| **Wins** | 88.86<br>(7.15) | 86.5<br>(6.62) | b = -2.35; *t*(263) = -2.71 | **0.007** | 0.027 | **Best Model (M1): Group (BF = 4.58)**<br>M2: Group (BF = 4.58) |
| **Win/Stay** | 60.56<br>(18.65) | 57.8<br>(16.25) | b = -3.68; *t*(263) = -1.77 | 0.08 | 0.012 | Best Model (M1): Age + Sex (BF > 1000)<br>M2: Age + Sex + Group (BF > 1000)<br>M1/M2 = 1.67 |
| **Win/Shift** | 16.59<br>(17.2) | 17.48<br>(14.79) | b = 1.83; *t*(263) = 0.98 | 0.33 | 0.004 | Best Model (M1): Age + Sex (BF > 1000)<br>M2: Age + Sex + Group (BF > 1000)<br>M1/M2 = 5.26 |
| **Lose/Stay** | 20.14<br>(13.3) | 23.3<br>(14.87) | b = 2.53; *t*(263) = 1.44 | 0.15 | 0.008 | Best Model (M1): Age (BF > 1000)<br>M2: Age + Group (BF = 173.52)<br>M1/M2 = 14.47 |
| **Lose/Shift** | 42.72<br>(13.82) | 41.41<br>(15.22) | b = -0.69; *t*(263) = -0.38 | 0.7 | 0.001 | Best Model (M1): Age (BF = 589.19)<br>M2: Age + Group (BF = 18.3) |
| **Full Sample: Late Trials** |  |  |  |  |  |  |
| **Wins** | 94.39<br>(8.74) | 93.94<br>(8.11) | b = -0.61; *t*(263) = -0.57 | 0.57 | 0.001 | Best Model (M1): Age (BF = 0.36)<br>M2: Age + Group (BF = 0.01) |
| **Win/Stay** | 80.09<br>(19.03) | 75.71<br>(18.95) | b = -5.31; *t*(263) = -2.3 | **0.02** | 0.02 | **Best Model (M1): Age + Group (BF > 1000)**<br>M2: Age (BF > 1000)<br>M1/M2 = 1.43 |
| **Win/Shift** | 14.38<br>(15.15) | 17.31<br>(15.11) | b = 3.73; *t*(263) = 2.04 | **0.04** | 0.016 | Best Model (M1): Age (BF > 1000)<br>M2: Age + Group (BF > 1000)<br>M1/M2 = 2.18 |
| **Lose/Stay** | 27.13<br>(15.26) | 27.18<br>(15.89) | b = -0.49; *t*(263) = -0.25 | 0.8 | 0 | Best Model (M1): Age (BF = 47.27)<br>M2: Age + Group (BF = 0.98) |
| **Lose/Shift** | 38.39<br>(16.52) | 39.8<br>(18.27) | b = 2.07; *t*(263) = 0.95 | 0.35 | 0.003 | Best Model (M1): Age (BF = 71.47)<br>M2: Age + Group (BF = 2.59) |
| **Subsample with WRAT scores: Early Trials** |  |  |  |  |  |  |
| **N =** | 87 | 120 |  |  |  |  |
| **Wins** | 88.86<br>(7.15) | 86.5<br>(6.62) | b = -1.27; *t*(202) = -1.17 | 0.24 | 0.007 | Best Model (M1): Group (BF = 0.59)<br>M2: Group (BF = 0.59) |
| **Win/Stay** | 60.56<br>(18.65) | 57.8<br>(16.25) | b = -5.59; *t*(202) = -2.18 | **0.03** | 0.023 | Best Model (M1): Age + Sex (BF > 1000)<br>M2: Age + Sex + Group (BF > 1000)<br>M1/M2 = 3.90 |
| **Win/Shift** | 16.59<br>(17.2) | 17.48<br>(14.79) | b = 4.71; *t*(202) = 1.98 | **0.05** | 0.019 | Best Model (M1): Age + Sex (BF > 1000)<br>M2: Age + Sex + Group (BF > 1000)<br>M1/M2 = 4.96 |
| **Lose/Stay** | 20.14<br>(13.3) | 23.3<br>(14.87) | b = 1.52; *t*(202) = 0.69 | 0.49 | 0.002 | Best Model (M1): Age (BF = 48.99)<br>M2: Age + Group (BF = 4.37) |
| **Lose/Shift** | 42.72<br>(13.82) | 41.41<br>(15.22) | b = -0.64; *t*(202) = -0.29 | 0.77 | 0 | Best Model (M1): Age (BF = 15.41)<br>M2: Age + Group (BF = 0.51) |
| **Subsample with WRAT scores: Late Trials** |  |  |  |  |  |  |
| **Wins** | 94.39<br>(8.74) | 93.94<br>(8.11) | b = -1.11; *t*(202) = -0.81 | 0.42 | 0.003 | Best Model (M1): Sex (BF = 0.33)<br>M2: Sex + Group (BF < .01) |
| **Win/Stay** | 80.09<br>(19.03) | 75.71<br>(18.95) | b = -7.35; *t*(202) = -2.5 | **0.01** | 0.03 | Best Model (M1): Age (BF = 102.96)<br>M2: Age + Group (BF = 19.33) |
| **Win/Shift** | 14.38<br>(15.15) | 17.31<br>(15.11) | b = 5.61; *t*(202) = 2.45 | **0.02** | 0.029 | Best Model (M1): Age (BF > 1000)<br>M2: Age + Group (BF = 160.69)<br>M1/M2 = 7.40 |
| **Lose/Stay** | 27.13<br>(15.26) | 27.18<br>(15.89) | b = -0.9; *t*(202) = -0.37 | 0.71 | 0.001 | Best Model (M1): Age (BF = 2.19)<br>M2: Age + Group (BF = 0.03) |
| **Lose/Shift** | 38.39<br>(16.52) | 39.8<br>(18.27) | b = 2.63; *t*(202) = 0.96 | 0.34 | 0.005 | Best Model (M1): Age (BF = 2.37)<br>M2: Age + Group (BF = 0.04) |

\*See note below **Table S.3** for interpretation of these Bayes factor analyses.

**Table S1.9.** Summary statistics (mean and SD) for parameters by specific disorder group in combined exploratory and confirmatory samples

|  | Action Precision | Reward Sensitivity | Learning Rate (Wins) | Learning Rate (Losses) | Information Insensitivity |
| --- | --- | --- | --- | --- | --- |
| HCs (N = 153) | 2.60 (0.99) | 4.59 (1.73) | 0.48 (0.13) | 0.41 (0.14) | 0.76 (0.32) |
| Only Stimulant (N = 55) | 2.21 (0.65) | 4.48 (2.27) | 0.51 (0.13) | 0.37 (0.16) | 0.81 (0.33) |
| Stimulant (N = 265) | 2.33 (1.23) | 4.36 (1.57) | 0.50 (0.13) | 0.37 (0.16) | 0.82 (0.30) |
| Cannabis (N = 153) | 2.26 (0.75) | 4.27 (1.36) | 0.50 (0.13) | 0.38 (0.15) | 0.80 (0.29) |
| Opioid (N = 118) | 2.26 (0.63) | 4.30 (1.42) | 0.48 (0.14) | 0.40 (0.15) | 0.78 (0.29) |
| Alcohol (N = 117) | 2.34 (1.64) | 4.35 (1.36) | 0.50 (0.13) | 0.37 (0.16) | 0.82 (0.27) |
| Sedative (N = 80) | 2.20 (0.63) | 4.24 (1.29) | 0.49 (0.14) | 0.39 (0.16) | 0.82 (0.27) |
| Hallucinogens (N = 14) | 2.03 (0.43) | 4.07 (1.57) | 0.51 (0.12) | 0.40 (0.15) | 0.66 (0.19) |
| MDD (N = 186) | 2.22 (0.70) | 4.20 (1.67) | 0.49 (0.14) | 0.38 (0.15) | 0.81 (0.30) |
| GAD (N = 57) | 2.28 (0.57) | 4.54 (1.19) | 0.49 (0.13) | 0.38 (0.16) | 0.85 (0.27) |
| SAD (N = 50) | 2.35 (0.72) | 4.10 (1.39) | 0.47 (0.15) | 0.39 (0.17) | 0.79 (0.30) |
| PTSD (N = 41) | 2.39 (0.76) | 4.39 (2.65) | 0.48 (0.12) | 0.40 (0.14) | 0.79 (0.30) |
| Panic (N = 28) | 2.17 (0.61) | 4.22 (1.57) | 0.51 (0.13) | 0.38 (0.15) | 0.76 (0.25) |

**Table S1.10.**
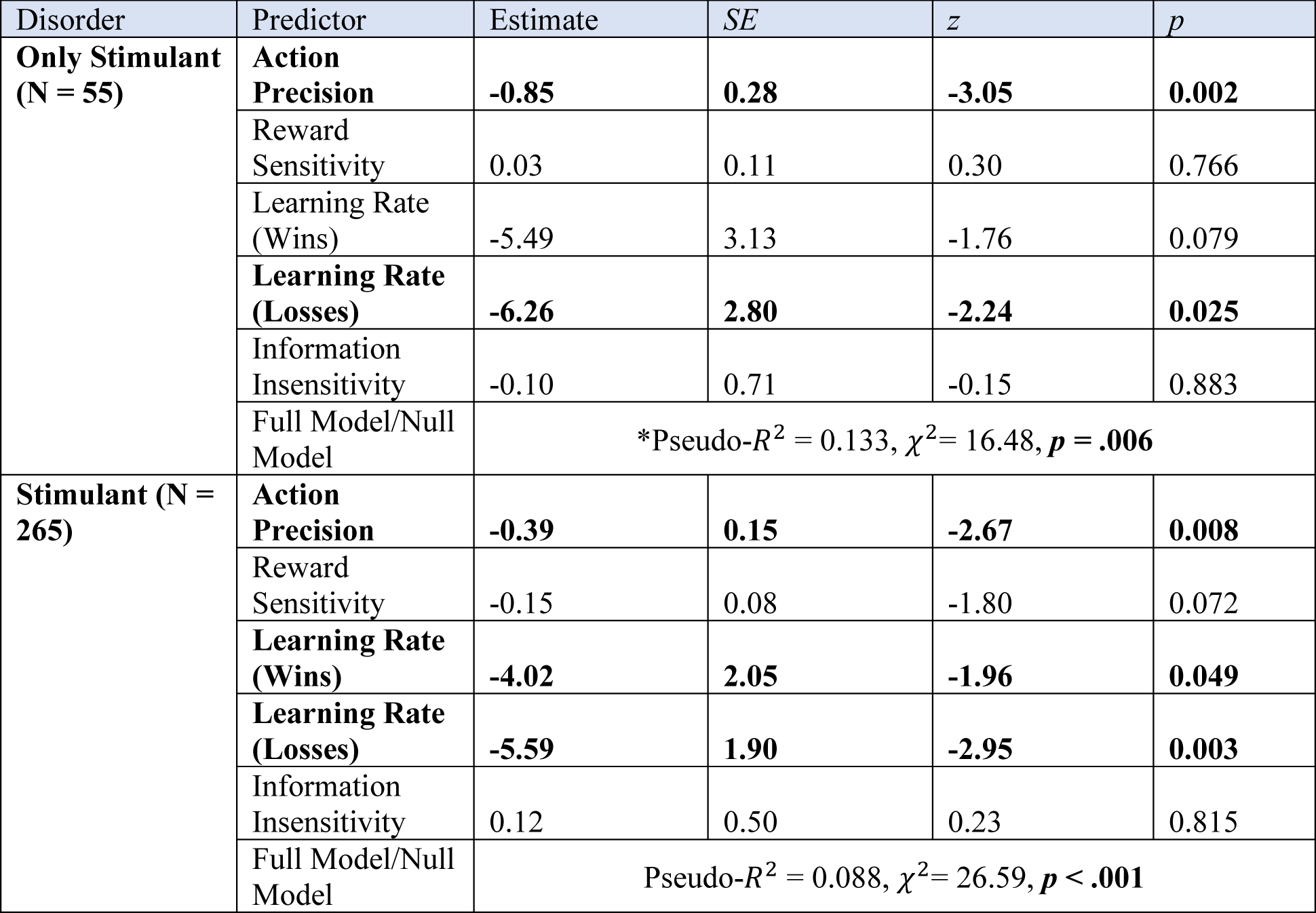
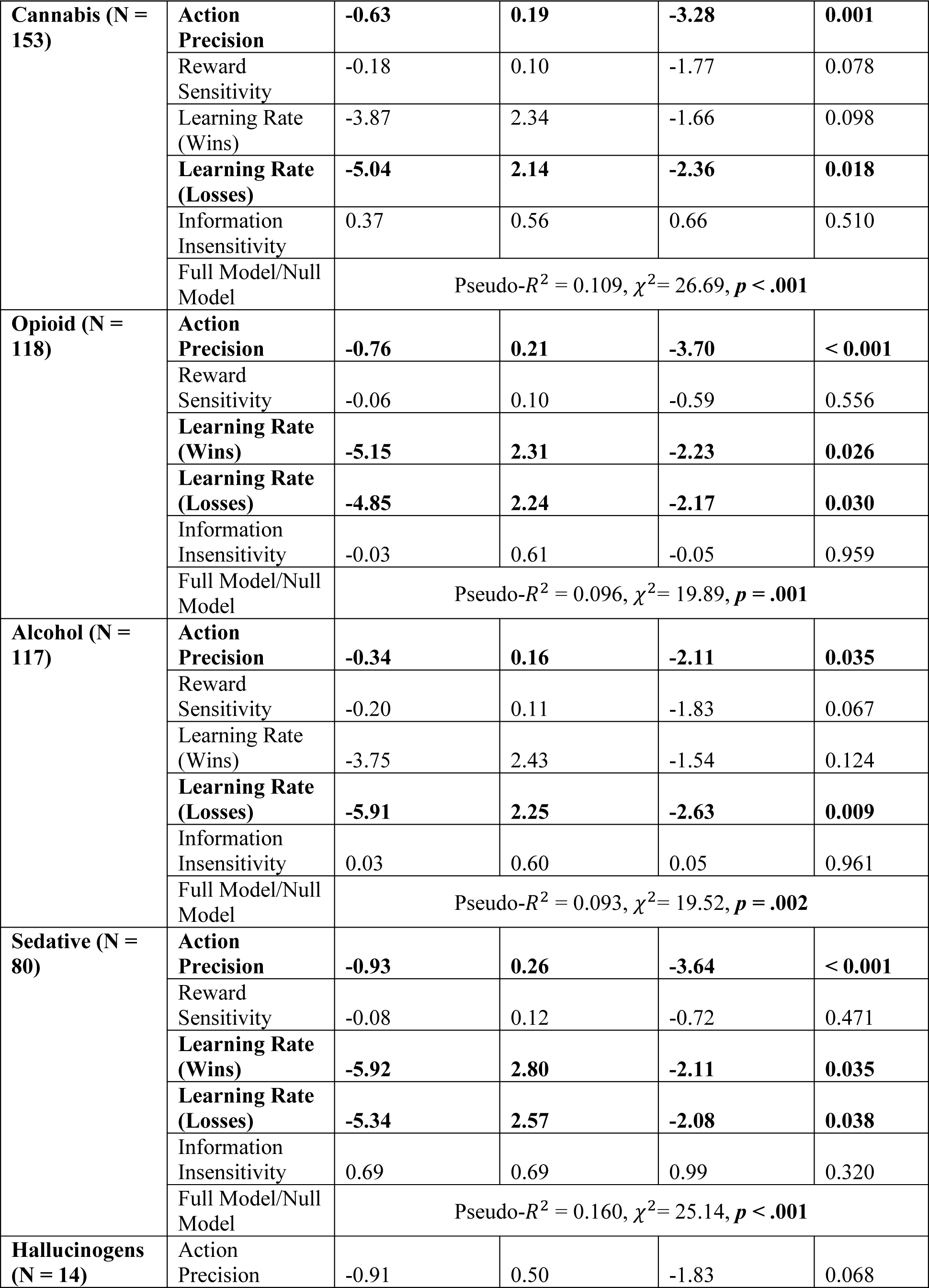
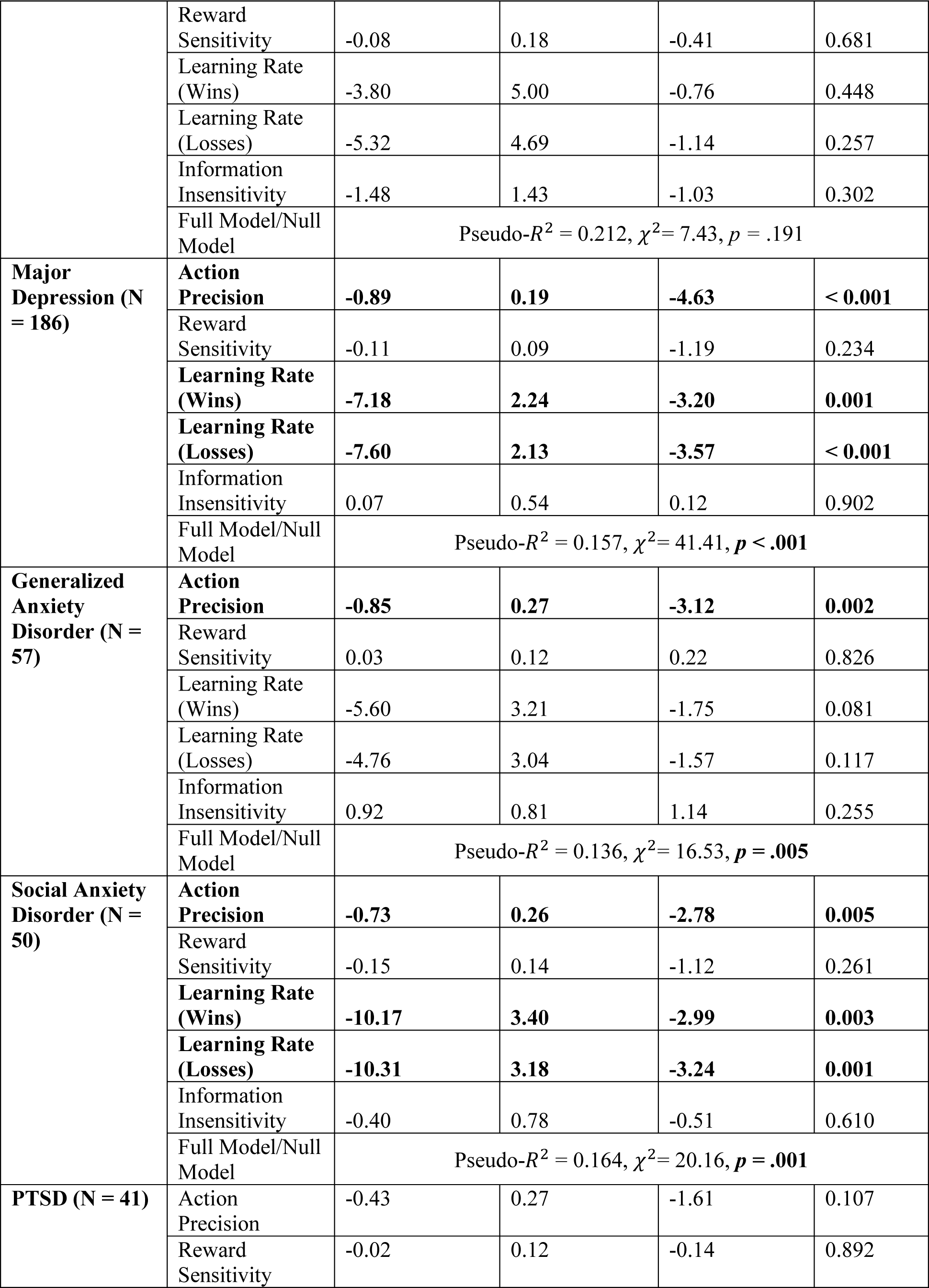
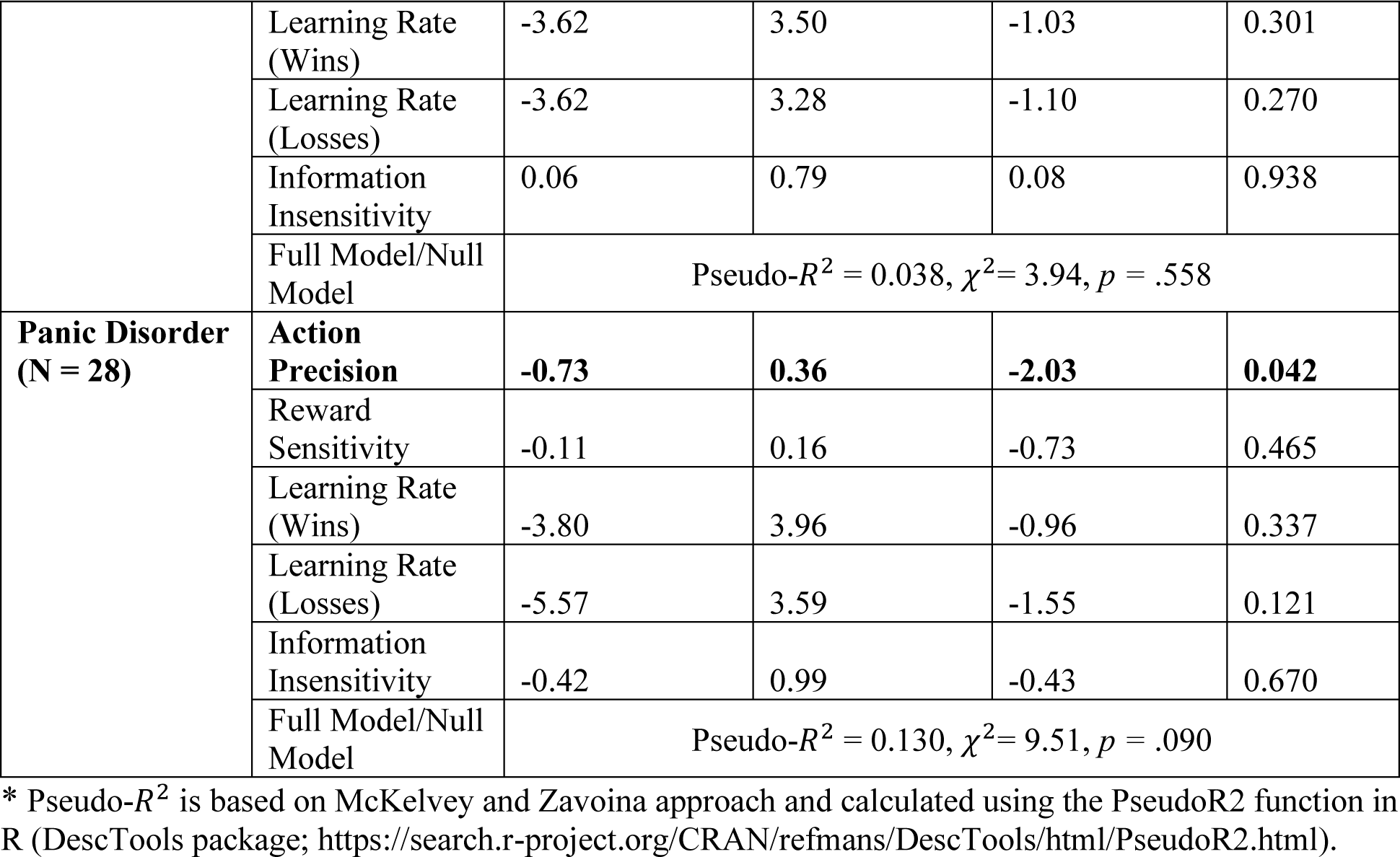
Logistic regressions predicting specific disorder vs. HCs (N = 153) in combined exploratory and confirmatory samples

| Disorder | Predictor | Estimate | SE | z | p |
| --- | --- | --- | --- | --- | --- |
| Only Stimulant (N = 55) | Action Precision | -0.85 | 0.28 | -3.05 | 0.002 |
|  | Reward Sensitivity | 0.03 | 0.11 | 0.30 | 0.766 |
|  | Learning Rate (Wins) | -5.49 | 3.13 | -1.76 | 0.079 |
|  | Learning Rate (Losses) | -6.26 | 2.80 | -2.24 | 0.025 |
|  | Information Insensitivity | -0.10 | 0.71 | -0.15 | 0.883 |
| | Full Model/Null Model | *Pseudo-$R^2 = 0.133$, $\chi^2 = 16.48$, $p = .006$ | | | |
| Stimulant (N = 265) | Action Precision | -0.39 | 0.15 | -2.67 | 0.008 |
|  | Reward Sensitivity | -0.15 | 0.08 | -1.80 | 0.072 |
|  | Learning Rate (Wins) | -4.02 | 2.05 | -1.96 | 0.049 |
|  | Learning Rate (Losses) | -5.59 | 1.90 | -2.95 | 0.003 |
|  | Information Insensitivity | 0.12 | 0.50 | 0.23 | 0.815 |
| | Full Model/Null Model | Pseudo-$R^2 = 0.088$, $\chi^2 = 26.59$, $p < .001$ | | | |
| **Cannabis (N = 153)** | **Action Precision** | **-0.63** | **0.19** | **-3.28** | **0.001** |
|  | Reward Sensitivity | -0.18 | 0.10 | -1.77 | 0.078 |
|  | Learning Rate (Wins) | -3.87 | 2.34 | -1.66 | 0.098 |
|  | **Learning Rate (Losses)** | **-5.04** | **2.14** | **-2.36** | **0.018** |
|  | Information Insensitivity | 0.37 | 0.56 | 0.66 | 0.510 |
| | Full Model/Null Model | Pseudo-$R^2 = 0.109$, $\chi^2 = 26.69$, $p < .001$ | | | |
| **Opioid (N = 118)** | **Action Precision** | **-0.76** | **0.21** | **-3.70** | **< 0.001** |
|  | Reward Sensitivity | -0.06 | 0.10 | -0.59 | 0.556 |
|  | **Learning Rate (Wins)** | **-5.15** | **2.31** | **-2.23** | **0.026** |
|  | **Learning Rate (Losses)** | **-4.85** | **2.24** | **-2.17** | **0.030** |
|  | Information Insensitivity | -0.03 | 0.61 | -0.05 | 0.959 |
| | Full Model/Null Model | Pseudo-$R^2 = 0.096$, $\chi^2 = 19.89$, $p = .001$ | | | |
| **Alcohol (N = 117)** | **Action Precision** | **-0.34** | **0.16** | **-2.11** | **0.035** |
|  | Reward Sensitivity | -0.20 | 0.11 | -1.83 | 0.067 |
|  | Learning Rate (Wins) | -3.75 | 2.43 | -1.54 | 0.124 |
|  | **Learning Rate (Losses)** | **-5.91** | **2.25** | **-2.63** | **0.009** |
|  | Information Insensitivity | 0.03 | 0.60 | 0.05 | 0.961 |
| | Full Model/Null Model | Pseudo-$R^2 = 0.093$, $\chi^2 = 19.52$, $p = .002$ | | | |
| **Sedative (N = 80)** | **Action Precision** | **-0.93** | **0.26** | **-3.64** | **< 0.001** |
|  | Reward Sensitivity | -0.08 | 0.12 | -0.72 | 0.471 |
|  | **Learning Rate (Wins)** | **-5.92** | **2.80** | **-2.11** | **0.035** |
|  | **Learning Rate (Losses)** | **-5.34** | **2.57** | **-2.08** | **0.038** |
|  | Information Insensitivity | 0.69 | 0.69 | 0.99 | 0.320 |
| | Full Model/Null Model | Pseudo-$R^2 = 0.160$, $\chi^2 = 25.14$, $p < .001$ | | | |
| **Hallucinogens (N = 14)** | Action Precision | -0.91 | 0.50 | -1.83 | 0.068 |
|  | Reward Sensitivity | -0.08 | 0.18 | -0.41 | 0.681 |
|  | Learning Rate (Wins) | -3.80 | 5.00 | -0.76 | 0.448 |
|  | Learning Rate (Losses) | -5.32 | 4.69 | -1.14 | 0.257 |
|  | Information Insensitivity | -1.48 | 1.43 | -1.03 | 0.302 |
| | Full Model/Null Model | Pseudo-$R^2 = 0.212$, $\chi^2 = 7.43$, $p = .191$ | | | |
| **Major Depression (N = 186)** | **Action Precision** | **-0.89** | **0.19** | **-4.63** | **< 0.001** |
|  | Reward Sensitivity | -0.11 | 0.09 | -1.19 | 0.234 |
|  | **Learning Rate (Wins)** | **-7.18** | **2.24** | **-3.20** | **0.001** |
|  | **Learning Rate (Losses)** | **-7.60** | **2.13** | **-3.57** | **< 0.001** |
|  | Information Insensitivity | 0.07 | 0.54 | 0.12 | 0.902 |
| | Full Model/Null Model | Pseudo-$R^2 = 0.157$, $\chi^2 = 41.41$, $p < .001$ | | | |
| **Generalized Anxiety Disorder (N = 57)** | **Action Precision** | **-0.85** | **0.27** | **-3.12** | **0.002** |
|  | Reward Sensitivity | 0.03 | 0.12 | 0.22 | 0.826 |
|  | Learning Rate (Wins) | -5.60 | 3.21 | -1.75 | 0.081 |
|  | Learning Rate (Losses) | -4.76 | 3.04 | -1.57 | 0.117 |
|  | Information Insensitivity | 0.92 | 0.81 | 1.14 | 0.255 |
| | Full Model/Null Model | Pseudo-$R^2 = 0.136$, $\chi^2 = 16.53$, $p = .005$ | | | |
| **Social Anxiety Disorder (N = 50)** | **Action Precision** | **-0.73** | **0.26** | **-2.78** | **0.005** |
|  | Reward Sensitivity | -0.15 | 0.14 | -1.12 | 0.261 |
|  | **Learning Rate (Wins)** | **-10.17** | **3.40** | **-2.99** | **0.003** |
|  | **Learning Rate (Losses)** | **-10.31** | **3.18** | **-3.24** | **0.001** |
|  | Information Insensitivity | -0.40 | 0.78 | -0.51 | 0.610 |
| | Full Model/Null Model | Pseudo-$R^2 = 0.164$, $\chi^2 = 20.16$, $p = .001$ | | | |
| **PTSD (N = 41)** | Action Precision | -0.43 | 0.27 | -1.61 | 0.107 |
|  | Reward Sensitivity | -0.02 | 0.12 | -0.14 | 0.892 |
|  | Learning Rate (Wins) | -3.62 | 3.50 | -1.03 | 0.301 |
|  | Learning Rate (Losses) | -3.62 | 3.28 | -1.10 | 0.270 |
|  | Information Insensitivity | 0.06 | 0.79 | 0.08 | 0.938 |
| | Full Model/Null Model | Pseudo-$R^2 = 0.038$, $\chi^2 = 3.94$, $p = .558$ | | | |
| **Panic Disorder (N = 28)** | **Action Precision** | **-0.73** | **0.36** | **-2.03** | **0.042** |
|  | Reward Sensitivity | -0.11 | 0.16 | -0.73 | 0.465 |
|  | Learning Rate (Wins) | -3.80 | 3.96 | -0.96 | 0.337 |
|  | Learning Rate (Losses) | -5.57 | 3.59 | -1.55 | 0.121 |
|  | Information Insensitivity | -0.42 | 0.99 | -0.43 | 0.670 |
| | Full Model/Null Model | Pseudo-$R^2 = 0.130$, $\chi^2 = 9.51$, $p = .090$ | | | |

\* Pseudo- $R^2$ is based on McKelvey and Zavoina approach and calculated using the PseudoR2 function in R (DescTools package; https://search.r-project.org/CRAN/refmans/DescTools/html/PseudoR2.html).

**Table S1.11.**
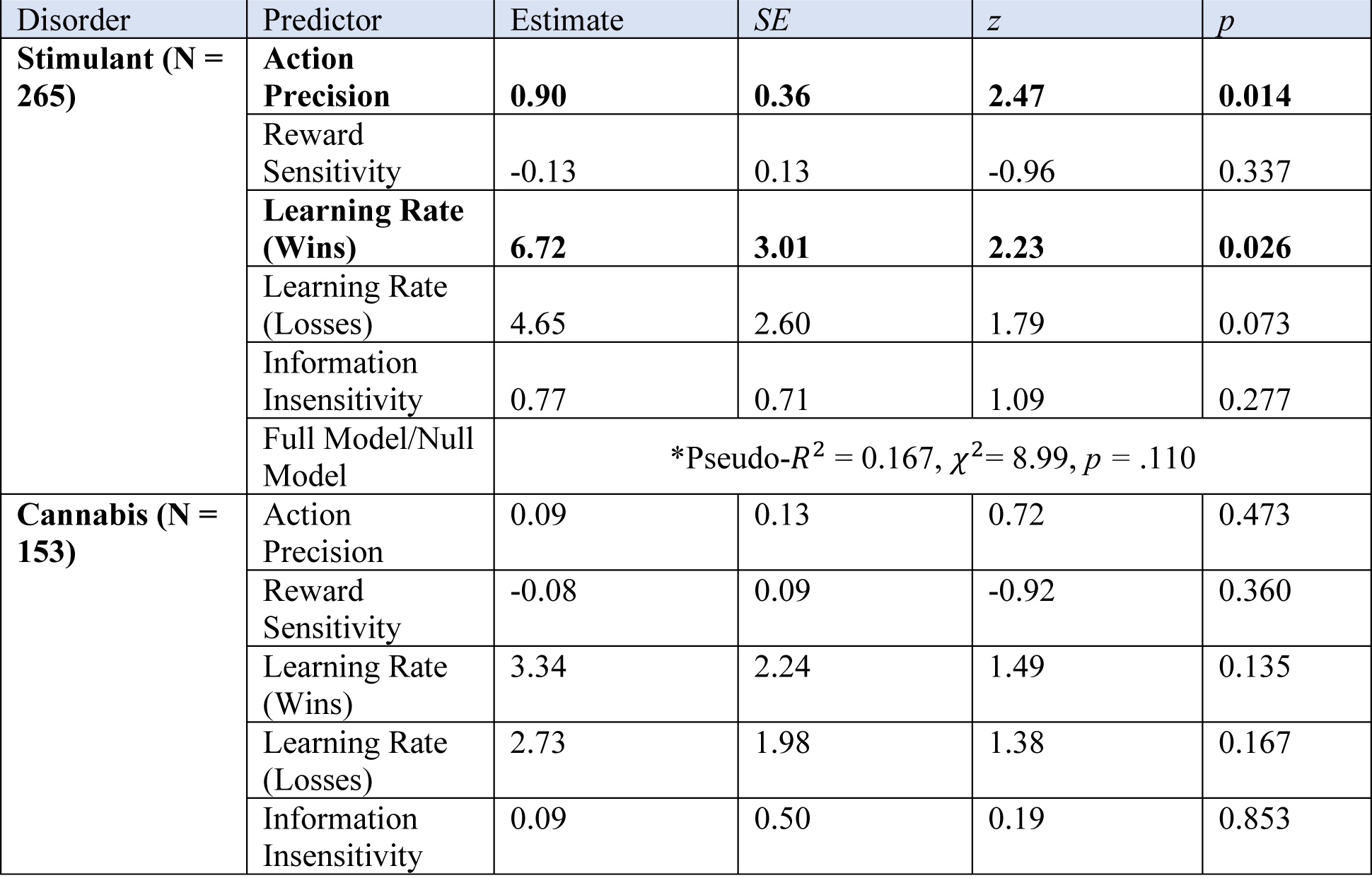
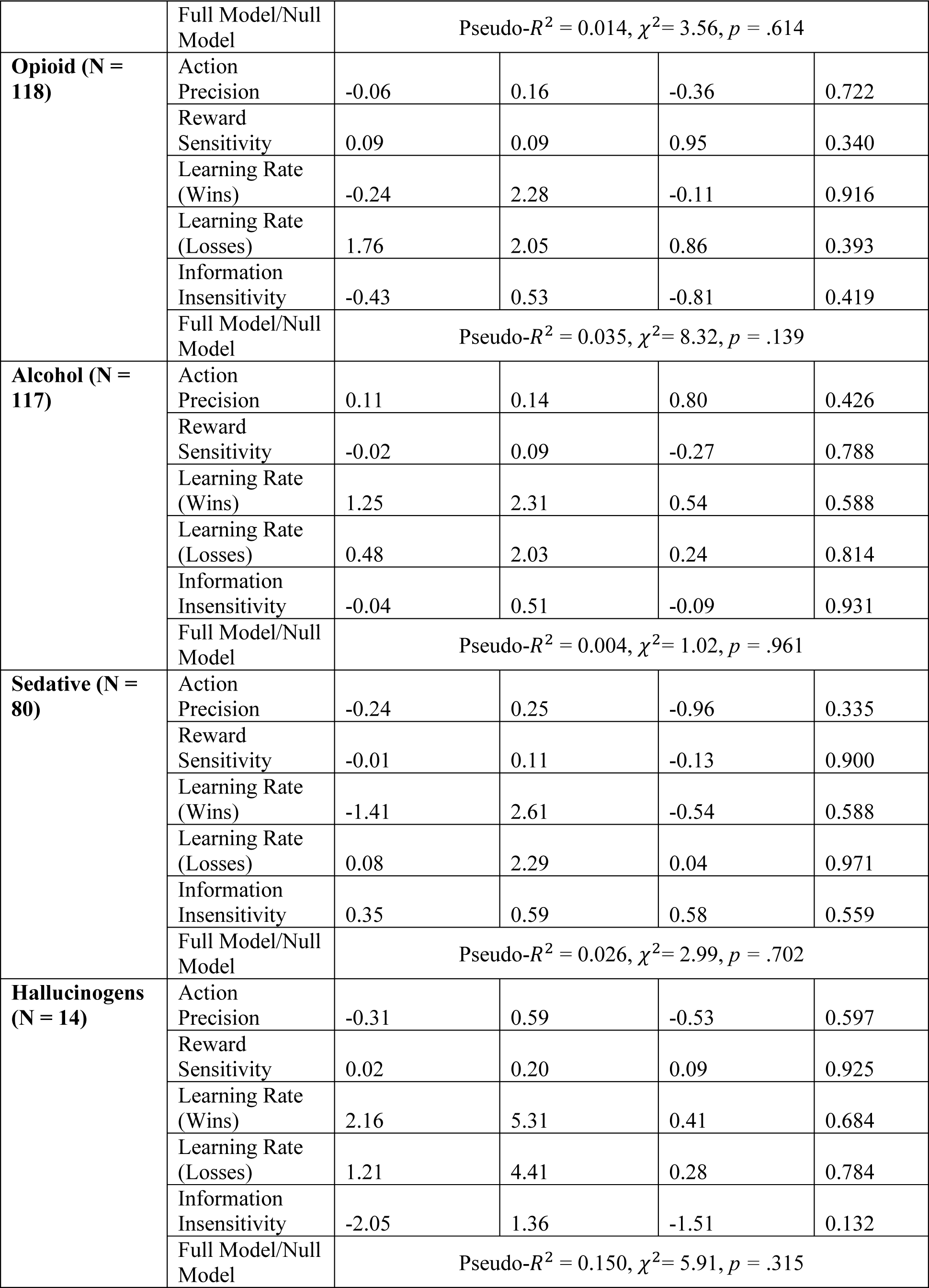
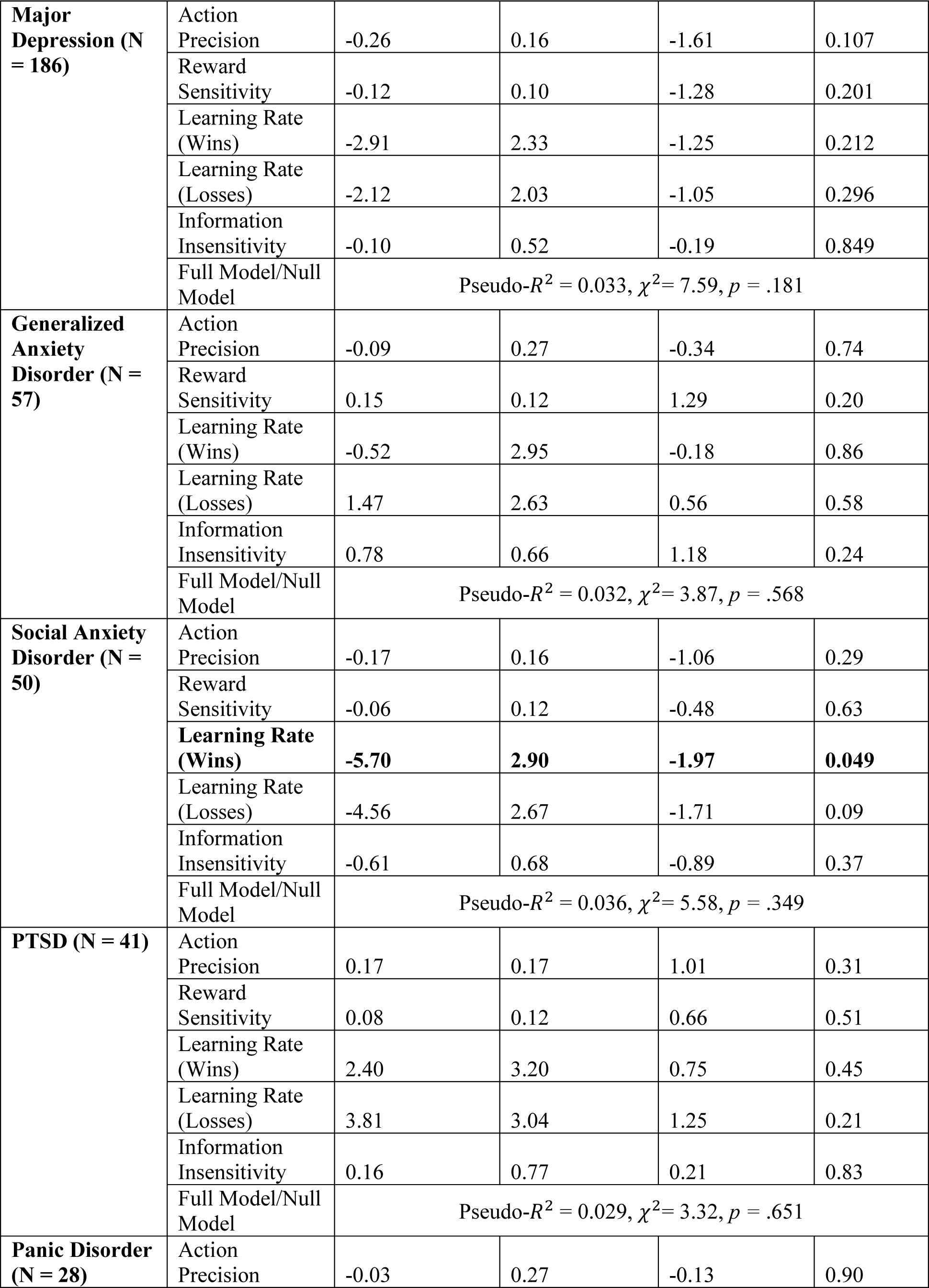
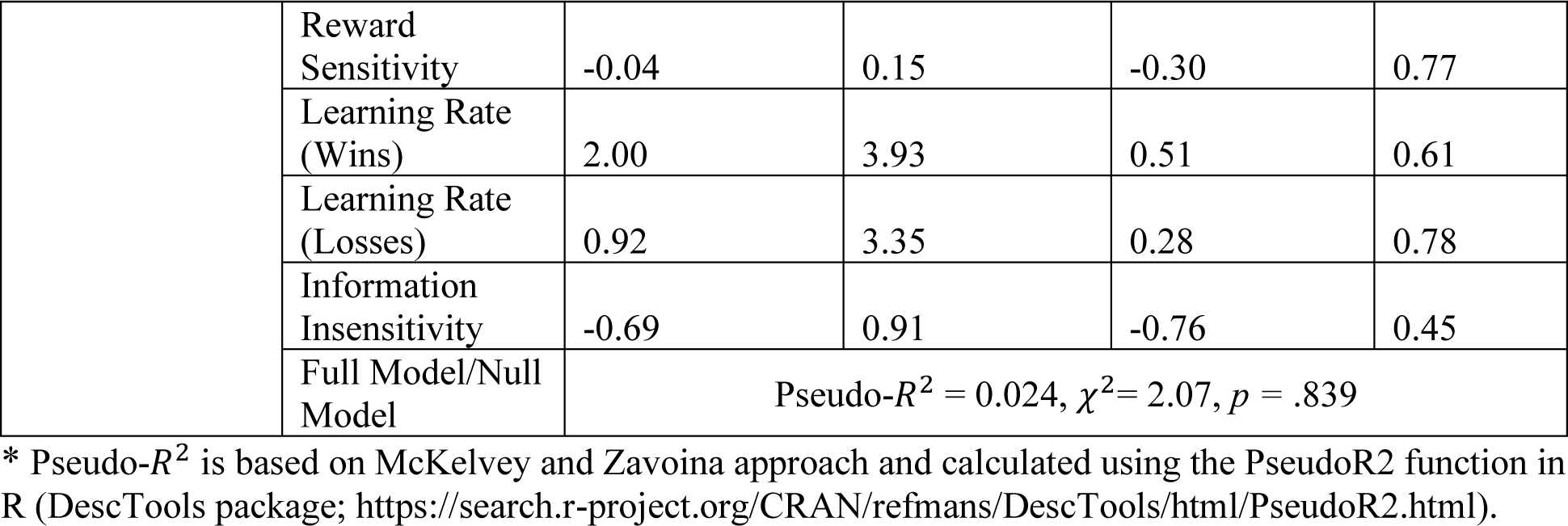
Logistic regressions predicting each specific disorder vs. all other disorders in combined samples

| Disorder | Predictor | Estimate | SE | z | p |
| --- | --- | --- | --- | --- | --- |
| **Stimulant (N = 265)** | **Action Precision** | **0.90** | **0.36** | **2.47** | **0.014** |
|  | Reward Sensitivity | -0.13 | 0.13 | -0.96 | 0.337 |
|  | **Learning Rate (Wins)** | **6.72** | **3.01** | **2.23** | **0.026** |
|  | Learning Rate (Losses) | 4.65 | 2.60 | 1.79 | 0.073 |
|  | Information Insensitivity | 0.77 | 0.71 | 1.09 | 0.277 |
| | Full Model/Null Model | *Pseudo-$R^2 = 0.167$, $\chi^2 = 8.99$, $p = .110$ | | | |
| **Cannabis (N = 153)** | Action Precision | 0.09 | 0.13 | 0.72 | 0.473 |
|  | Reward Sensitivity | -0.08 | 0.09 | -0.92 | 0.360 |
|  | Learning Rate (Wins) | 3.34 | 2.24 | 1.49 | 0.135 |
|  | Learning Rate (Losses) | 2.73 | 1.98 | 1.38 | 0.167 |
|  | Information Insensitivity | 0.09 | 0.50 | 0.19 | 0.853 |
| | Full Model/Null Model | Pseudo-$R^2 = 0.014$, $\chi^2 = 3.56$, $p = .614$ | | | |
| **Opioid (N = 118)** | Action Precision | -0.06 | 0.16 | -0.36 | 0.722 |
|  | Reward Sensitivity | 0.09 | 0.09 | 0.95 | 0.340 |
|  | Learning Rate (Wins) | -0.24 | 2.28 | -0.11 | 0.916 |
|  | Learning Rate (Losses) | 1.76 | 2.05 | 0.86 | 0.393 |
|  | Information Insensitivity | -0.43 | 0.53 | -0.81 | 0.419 |
| | Full Model/Null Model | Pseudo-$R^2 = 0.035$, $\chi^2 = 8.32$, $p = .139$ | | | |
| **Alcohol (N = 117)** | Action Precision | 0.11 | 0.14 | 0.80 | 0.426 |
|  | Reward Sensitivity | -0.02 | 0.09 | -0.27 | 0.788 |
|  | Learning Rate (Wins) | 1.25 | 2.31 | 0.54 | 0.588 |
|  | Learning Rate (Losses) | 0.48 | 2.03 | 0.24 | 0.814 |
|  | Information Insensitivity | -0.04 | 0.51 | -0.09 | 0.931 |
| | Full Model/Null Model | Pseudo-$R^2 = 0.004$, $\chi^2 = 1.02$, $p = .961$ | | | |
| **Sedative (N = 80)** | Action Precision | -0.24 | 0.25 | -0.96 | 0.335 |
|  | Reward Sensitivity | -0.01 | 0.11 | -0.13 | 0.900 |
|  | Learning Rate (Wins) | -1.41 | 2.61 | -0.54 | 0.588 |
|  | Learning Rate (Losses) | 0.08 | 2.29 | 0.04 | 0.971 |
|  | Information Insensitivity | 0.35 | 0.59 | 0.58 | 0.559 |
| | Full Model/Null Model | Pseudo-$R^2 = 0.026$, $\chi^2 = 2.99$, $p = .702$ | | | |
| **Hallucinogens (N = 14)** | Action Precision | -0.31 | 0.59 | -0.53 | 0.597 |
|  | Reward Sensitivity | 0.02 | 0.20 | 0.09 | 0.925 |
|  | Learning Rate (Wins) | 2.16 | 5.31 | 0.41 | 0.684 |
|  | Learning Rate (Losses) | 1.21 | 4.41 | 0.28 | 0.784 |
|  | Information Insensitivity | -2.05 | 1.36 | -1.51 | 0.132 |
| | Full Model/Null Model | Pseudo-$R^2 = 0.150$, $\chi^2 = 5.91$, $p = .315$ | | | |
| **Major Depression (N = 186)** | Action Precision | -0.26 | 0.16 | -1.61 | 0.107 |
|  | Reward Sensitivity | -0.12 | 0.10 | -1.28 | 0.201 |
|  | Learning Rate (Wins) | -2.91 | 2.33 | -1.25 | 0.212 |
|  | Learning Rate (Losses) | -2.12 | 2.03 | -1.05 | 0.296 |
|  | Information Insensitivity | -0.10 | 0.52 | -0.19 | 0.849 |
| | Full Model/Null Model | Pseudo-$R^2 = 0.033$, $\chi^2 = 7.59$, $p = .181$ | | | |
| **Generalized Anxiety Disorder (N = 57)** | Action Precision | -0.09 | 0.27 | -0.34 | 0.74 |
|  | Reward Sensitivity | 0.15 | 0.12 | 1.29 | 0.20 |
|  | Learning Rate (Wins) | -0.52 | 2.95 | -0.18 | 0.86 |
|  | Learning Rate (Losses) | 1.47 | 2.63 | 0.56 | 0.58 |
|  | Information Insensitivity | 0.78 | 0.66 | 1.18 | 0.24 |
| | Full Model/Null Model | Pseudo-$R^2 = 0.032$, $\chi^2 = 3.87$, $p = .568$ | | | |
| **Social Anxiety Disorder (N = 50)** | Action Precision | -0.17 | 0.16 | -1.06 | 0.29 |
|  | Reward Sensitivity | -0.06 | 0.12 | -0.48 | 0.63 |
|  | **Learning Rate (Wins)** | **-5.70** | **2.90** | **-1.97** | **0.049** |
|  | Learning Rate (Losses) | -4.56 | 2.67 | -1.71 | 0.09 |
|  | Information Insensitivity | -0.61 | 0.68 | -0.89 | 0.37 |
| | Full Model/Null Model | Pseudo-$R^2 = 0.036$, $\chi^2 = 5.58$, $p = .349$ | | | |
| **PTSD (N = 41)** | Action Precision | 0.17 | 0.17 | 1.01 | 0.31 |
|  | Reward Sensitivity | 0.08 | 0.12 | 0.66 | 0.51 |
|  | Learning Rate (Wins) | 2.40 | 3.20 | 0.75 | 0.45 |
|  | Learning Rate (Losses) | 3.81 | 3.04 | 1.25 | 0.21 |
|  | Information Insensitivity | 0.16 | 0.77 | 0.21 | 0.83 |
| | Full Model/Null Model | Pseudo-$R^2 = 0.029$, $\chi^2 = 3.32$, $p = .651$ | | | |
| **Panic Disorder (N = 28)** | Action Precision | -0.03 | 0.27 | -0.13 | 0.90 |
|  | Reward Sensitivity | -0.04 | 0.15 | -0.30 | 0.77 |
|  | Learning Rate (Wins) | 2.00 | 3.93 | 0.51 | 0.61 |
|  | Learning Rate (Losses) | 0.92 | 3.35 | 0.28 | 0.78 |
|  | Information Insensitivity | -0.69 | 0.91 | -0.76 | 0.45 |
| | Full Model/Null Model | Pseudo-$R^2 = 0.024$, $\chi^2 = 2.07$, $p = .839$ | | | |

\* Pseudo- $R^2$ is based on McKelvey and Zavoina approach and calculated using the PseudoR2 function in R (DescTools package; https://search.r-project.org/CRAN/refmans/DescTools/html/PseudoR2.html).

### Supplementary Figures

**Figure S1.1:**
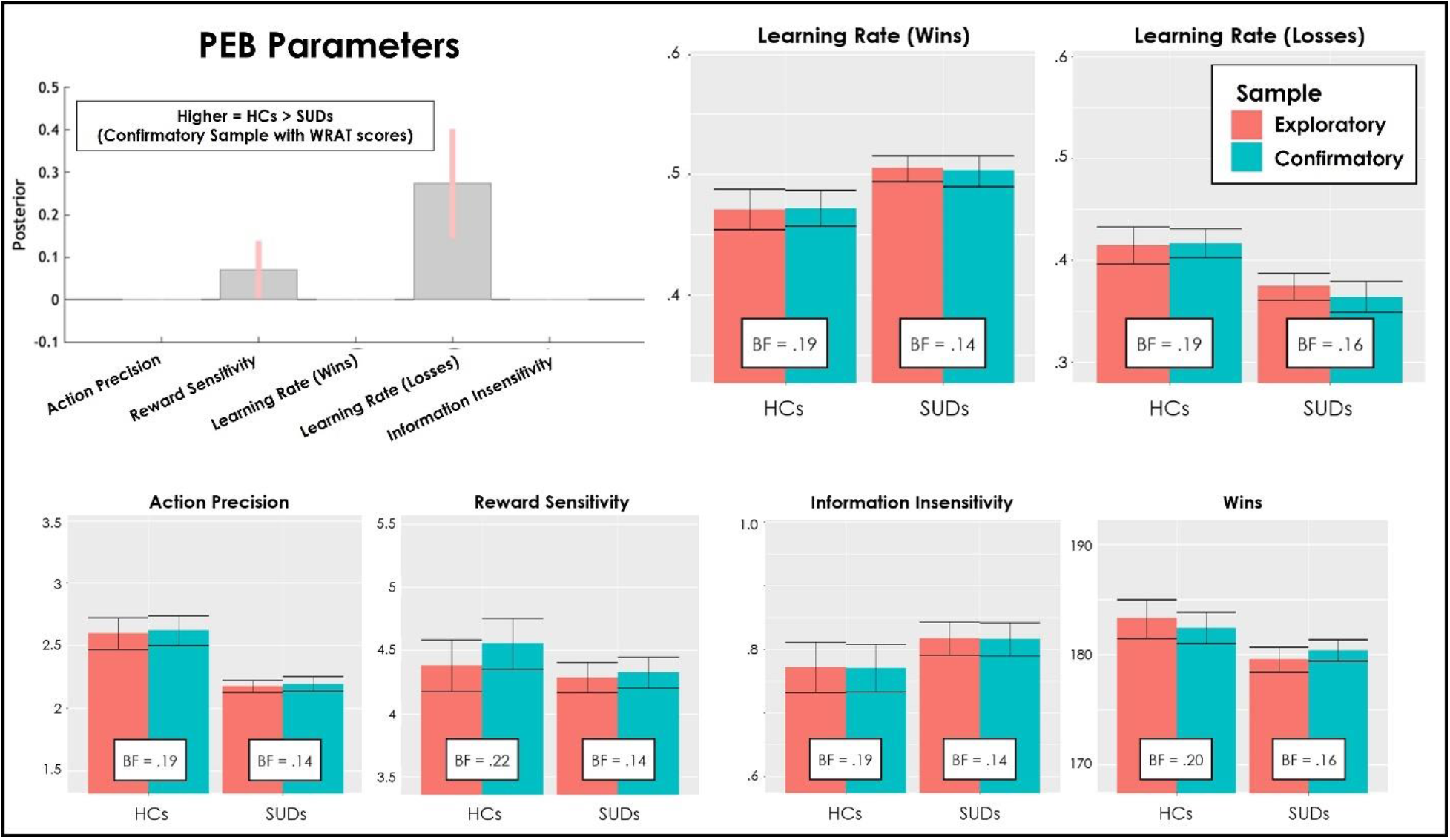
The top-left panel depicts results from Parametric Empirical Bayes (PEB) analyses showing the posterior means and credible intervals for effects of group (when accounting effects of age, sex, and WRAT reading scores). Positive effects indicate greater values in HCs relative to SUDs. Learning rates are estimated/displayed in logit space, while all other parameters are in log space. Results replicate the group difference in learning rate for losses found in our prior study (posterior probability [*pp*] = 0.99). There was also some evidence for differences in reward sensitivity not observed previously (greater in HCs; *pp* =0.72). All other panels compare the means and standard errors for each parameter in our prior study (exploratory sample; 54 HCs, 147 SUDs) to those in the current study (confirmatory sample, including only individuals with available WRAT reading scores; 87 HCs, 120 SUDs), when separating parameters by group. Also shown are Bayes factors (BFs) evaluating the evidence for differences between the two samples (BF < 1 indicates greater evidence for the absence of a difference). As can be seen, the BFs range from.14 to .22, indicating that the data are between 4.5 and 7.1 times more likely under a model with no difference between samples, which supports the idea that the new sample successfully replicates the findings in our prior study. For an analogous plot when including all participants in the confirmatory sample, see Figure 3 in the main text.

**Figure S1.2:**
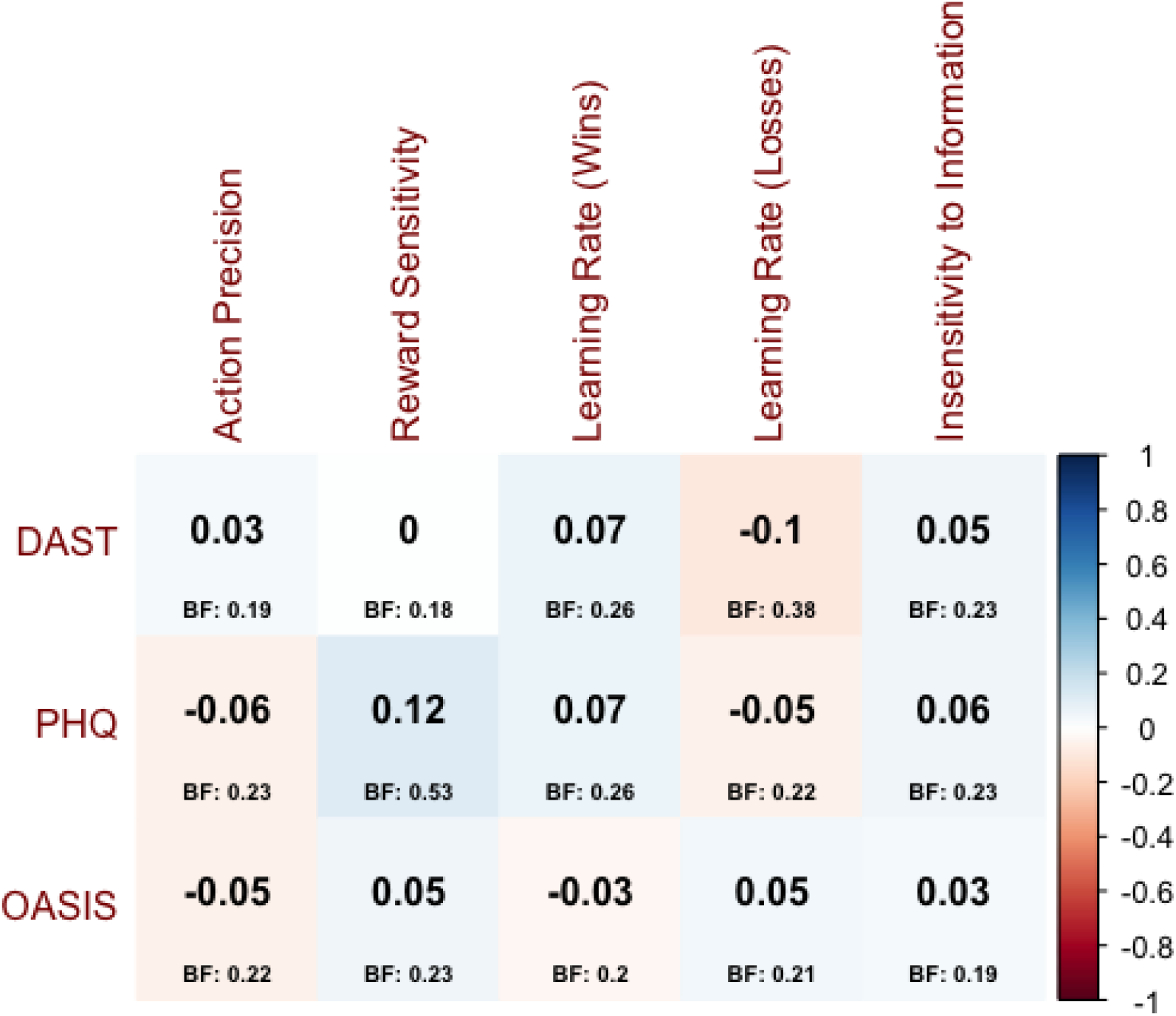
Correlations between symptom measures (rows) and model parameters (columns). No significant associations were observed. This failed to replicate the relationships observed in our previous study between PHQ/OASIS and information insensitivity, and between OASIS and reward sensitivity.

**Figure S1.3:**
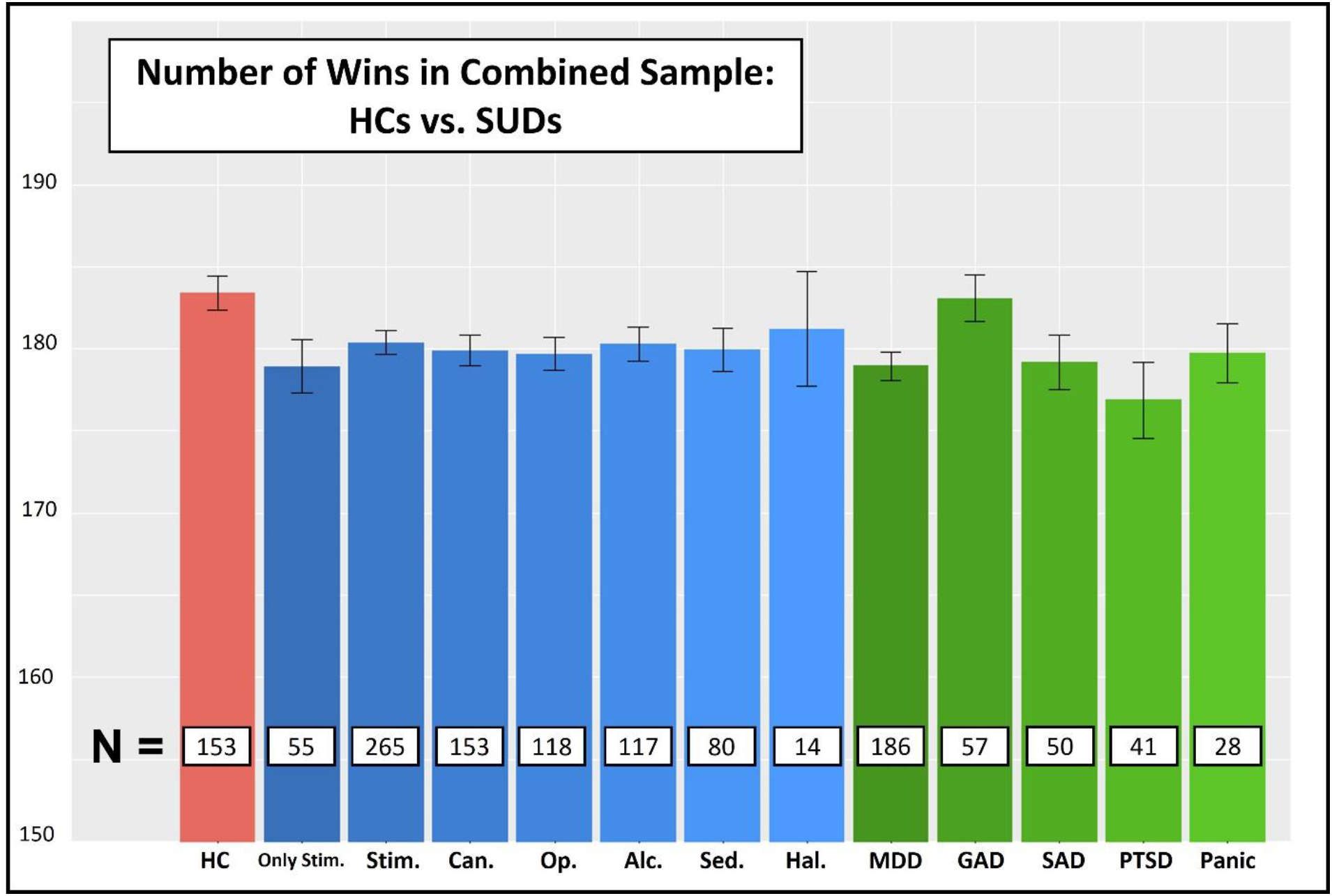
Task performance (number of wins) divided by group across the combined exploratory and confirmatory samples. Each column includes all participants who had a particular diagnosis. Thus, there is partial overlap across groups (with the exception of 55 individuals who had stimulant use disorder without co-morbidities; second bar from the left). These descriptive results suggest that the significantly greater wins seen in HCs than SUDs overall reflect a similar pattern across individuals with different SUDs.

## Supplementary Materials 2: Detailed Linear Model Statistics for All Effects

**Table S2.1** and **S2.2** below provide full statistics for all effects associated with results reported in **Table S1.5** in **Supplementary Materials 1**.

**Tables S2.3** and **S2.4** below provide full statistics for all effects associated with results reported in **Tables 4** and **5** in the main text and **Tables S1.6** and **S1.7** in **Supplementary Materials 1**.

**Table S2.1:**
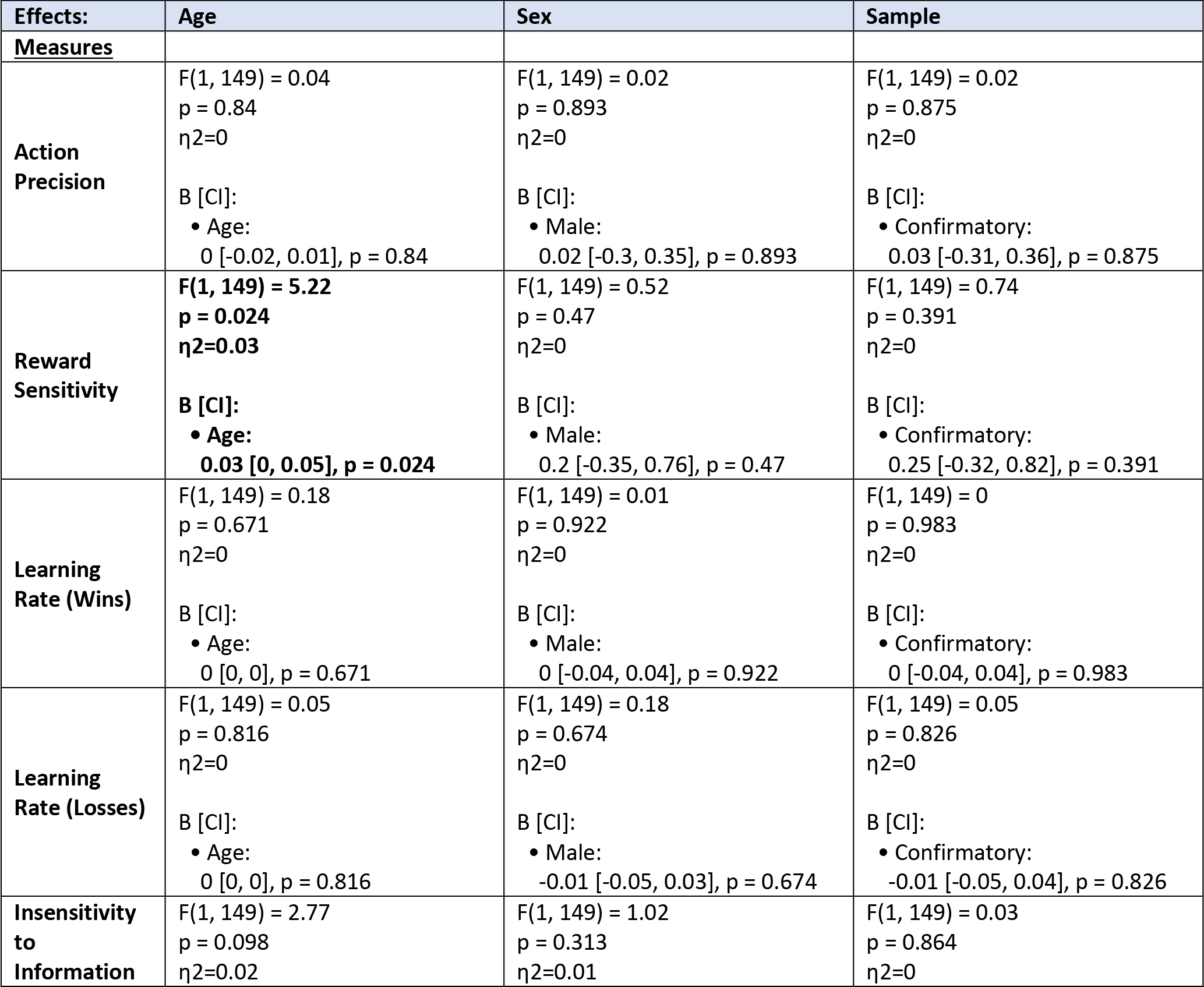
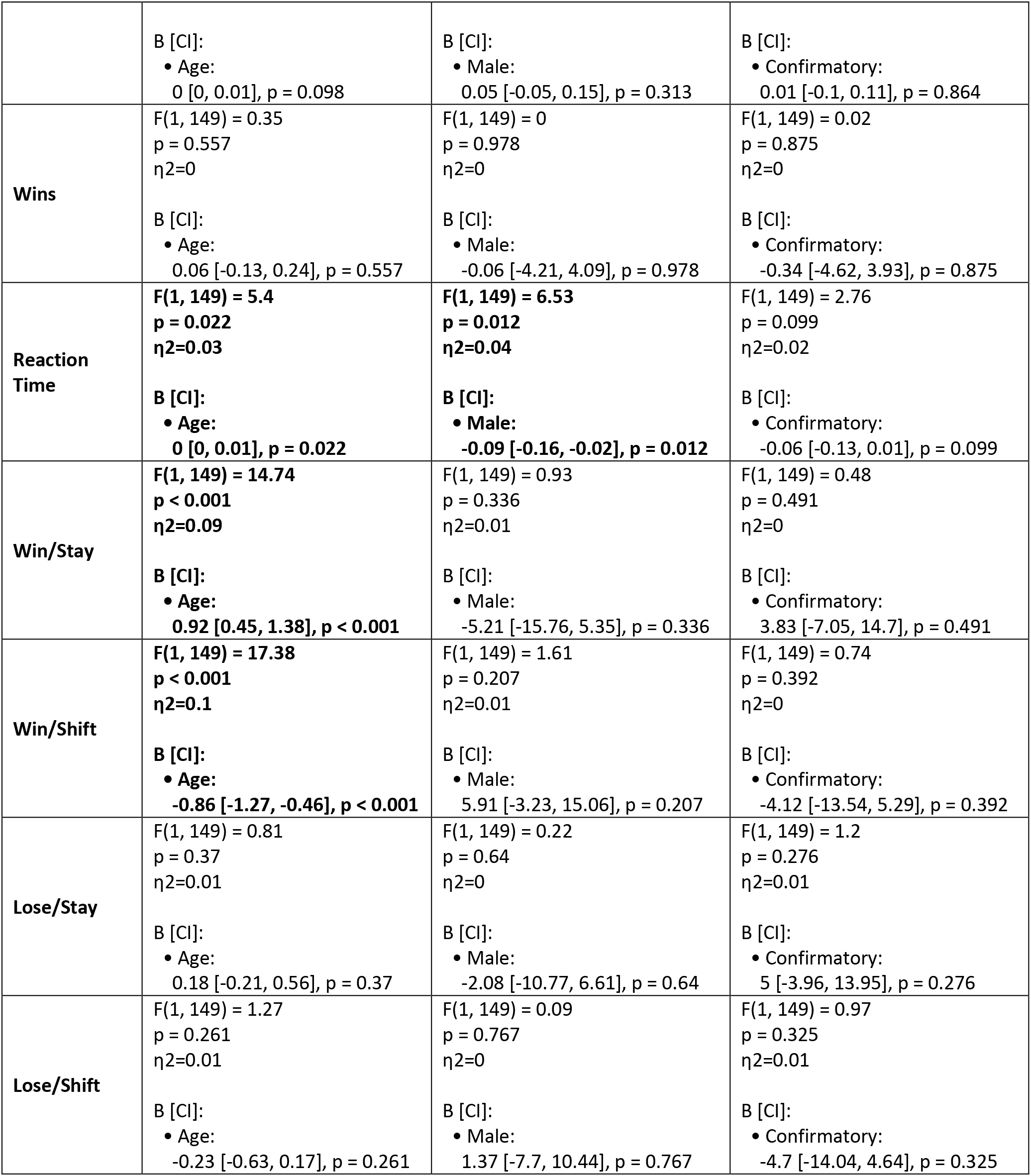
Full Linear Model Statistics Comparing HCs Between Exploratory and Confirmatory Samples

| Effects: | Age | Sex | Sample |
| --- | --- | --- | --- |
| Measures |  |  |  |
| **Action Precision** | $F(1, 149) = 0.04$<br>$p = 0.84$<br>$\eta^2 = 0$<br>B [CI]:<br>• Age:<br>$0 [-0.02, 0.01], p = 0.84$ | $F(1, 149) = 0.02$<br>$p = 0.893$<br>$\eta^2 = 0$<br>B [CI]:<br>• Male:<br>$0.02 [-0.3, 0.35], p = 0.893$ | $F(1, 149) = 0.02$<br>$p = 0.875$<br>$\eta^2 = 0$<br>B [CI]:<br>• Confirmatory:<br>$0.03 [-0.31, 0.36], p = 0.875$ |
| **Reward Sensitivity** | $F(1, 149) = 5.22$<br>$p = 0.024$<br>$\eta^2 = 0.03$<br>B [CI]:<br>• Age:<br>$0.03 [0, 0.05], p = 0.024$ | $F(1, 149) = 0.52$<br>$p = 0.47$<br>$\eta^2 = 0$<br>B [CI]:<br>• Male:<br>$0.2 [-0.35, 0.76], p = 0.47$ | $F(1, 149) = 0.74$<br>$p = 0.391$<br>$\eta^2 = 0$<br>B [CI]:<br>• Confirmatory:<br>$0.25 [-0.32, 0.82], p = 0.391$ |
| **Learning Rate (Wins)** | $F(1, 149) = 0.18$<br>$p = 0.671$<br>$\eta^2 = 0$<br>B [CI]:<br>• Age:<br>$0 [0, 0], p = 0.671$ | $F(1, 149) = 0.01$<br>$p = 0.922$<br>$\eta^2 = 0$<br>B [CI]:<br>• Male:<br>$0 [-0.04, 0.04], p = 0.922$ | $F(1, 149) = 0$<br>$p = 0.983$<br>$\eta^2 = 0$<br>B [CI]:<br>• Confirmatory:<br>$0 [-0.04, 0.04], p = 0.983$ |
| **Learning Rate (Losses)** | $F(1, 149) = 0.05$<br>$p = 0.816$<br>$\eta^2 = 0$<br>B [CI]:<br>• Age:<br>$0 [0, 0], p = 0.816$ | $F(1, 149) = 0.18$<br>$p = 0.674$<br>$\eta^2 = 0$<br>B [CI]:<br>• Male:<br>$-0.01 [-0.05, 0.03], p = 0.674$ | $F(1, 149) = 0.05$<br>$p = 0.826$<br>$\eta^2 = 0$<br>B [CI]:<br>• Confirmatory:<br>$-0.01 [-0.05, 0.04], p = 0.826$ |
| **Insensitivity to Information** | $F(1, 149) = 2.77$<br>$p = 0.098$<br>$\eta^2 = 0.02$ | $F(1, 149) = 1.02$<br>$p = 0.313$<br>$\eta^2 = 0.01$ | $F(1, 149) = 0.03$<br>$p = 0.864$<br>$\eta^2 = 0$ |
| | B [CI]:<br>• Age:<br>0 [0, 0.01], $p = 0.098$ | B [CI]:<br>• Male:<br>0.05 [-0.05, 0.15], $p = 0.313$ | B [CI]:<br>• Confirmatory:<br>0.01 [-0.1, 0.11], $p = 0.864$ |
| **Wins** | F(1, 149) = 0.35<br>$p = 0.557$<br>$\eta^2 = 0$<br>B [CI]:<br>• Age:<br>0.06 [-0.13, 0.24], $p = 0.557$ | F(1, 149) = 0<br>$p = 0.978$<br>$\eta^2 = 0$<br>B [CI]:<br>• Male:<br>-0.06 [-4.21, 4.09], $p = 0.978$ | F(1, 149) = 0.02<br>$p = 0.875$<br>$\eta^2 = 0$<br>B [CI]:<br>• Confirmatory:<br>-0.34 [-4.62, 3.93], $p = 0.875$ |
| **Reaction Time** | **F(1, 149) = 5.4**<br>**$p = 0.022$**<br>**$\eta^2 = 0.03$**<br>B [CI]:<br>• Age:<br>0 [0, 0.01], $p = 0.022$ | **F(1, 149) = 6.53**<br>**$p = 0.012$**<br>**$\eta^2 = 0.04$**<br>B [CI]:<br>• Male:<br>-0.09 [-0.16, -0.02], $p = 0.012$ | F(1, 149) = 2.76<br>$p = 0.099$<br>$\eta^2 = 0.02$<br>B [CI]:<br>• Confirmatory:<br>-0.06 [-0.13, 0.01], $p = 0.099$ |
| **Win/Stay** | **F(1, 149) = 14.74**<br>**$p < 0.001$**<br>**$\eta^2 = 0.09$**<br>B [CI]:<br>• Age:<br>0.92 [0.45, 1.38], $p < 0.001$ | F(1, 149) = 0.93<br>$p = 0.336$<br>$\eta^2 = 0.01$<br>B [CI]:<br>• Male:<br>-5.21 [-15.76, 5.35], $p = 0.336$ | F(1, 149) = 0.48<br>$p = 0.491$<br>$\eta^2 = 0$<br>B [CI]:<br>• Confirmatory:<br>3.83 [-7.05, 14.7], $p = 0.491$ |
| **Win/Shift** | **F(1, 149) = 17.38**<br>**$p < 0.001$**<br>**$\eta^2 = 0.1$**<br>B [CI]:<br>• Age:<br>-0.86 [-1.27, -0.46], $p < 0.001$ | F(1, 149) = 1.61<br>$p = 0.207$<br>$\eta^2 = 0.01$<br>B [CI]:<br>• Male:<br>5.91 [-3.23, 15.06], $p = 0.207$ | F(1, 149) = 0.74<br>$p = 0.392$<br>$\eta^2 = 0$<br>B [CI]:<br>• Confirmatory:<br>-4.12 [-13.54, 5.29], $p = 0.392$ |
| **Lose/Stay** | F(1, 149) = 0.81<br>$p = 0.37$<br>$\eta^2 = 0.01$<br>B [CI]:<br>• Age:<br>0.18 [-0.21, 0.56], $p = 0.37$ | F(1, 149) = 0.22<br>$p = 0.64$<br>$\eta^2 = 0$<br>B [CI]:<br>• Male:<br>-2.08 [-10.77, 6.61], $p = 0.64$ | F(1, 149) = 1.2<br>$p = 0.276$<br>$\eta^2 = 0.01$<br>B [CI]:<br>• Confirmatory:<br>5 [-3.96, 13.95], $p = 0.276$ |
| **Lose/Shift** | F(1, 149) = 1.27<br>$p = 0.261$<br>$\eta^2 = 0.01$<br>B [CI]:<br>• Age:<br>-0.23 [-0.63, 0.17], $p = 0.261$ | F(1, 149) = 0.09<br>$p = 0.767$<br>$\eta^2 = 0$<br>B [CI]:<br>• Male:<br>1.37 [-7.7, 10.44], $p = 0.767$ | F(1, 149) = 0.97<br>$p = 0.325$<br>$\eta^2 = 0.01$<br>B [CI]:<br>• Confirmatory:<br>-4.7 [-14.04, 4.64], $p = 0.325$ |

**Table S2.2:**
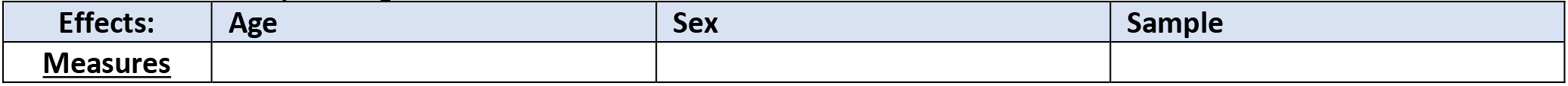
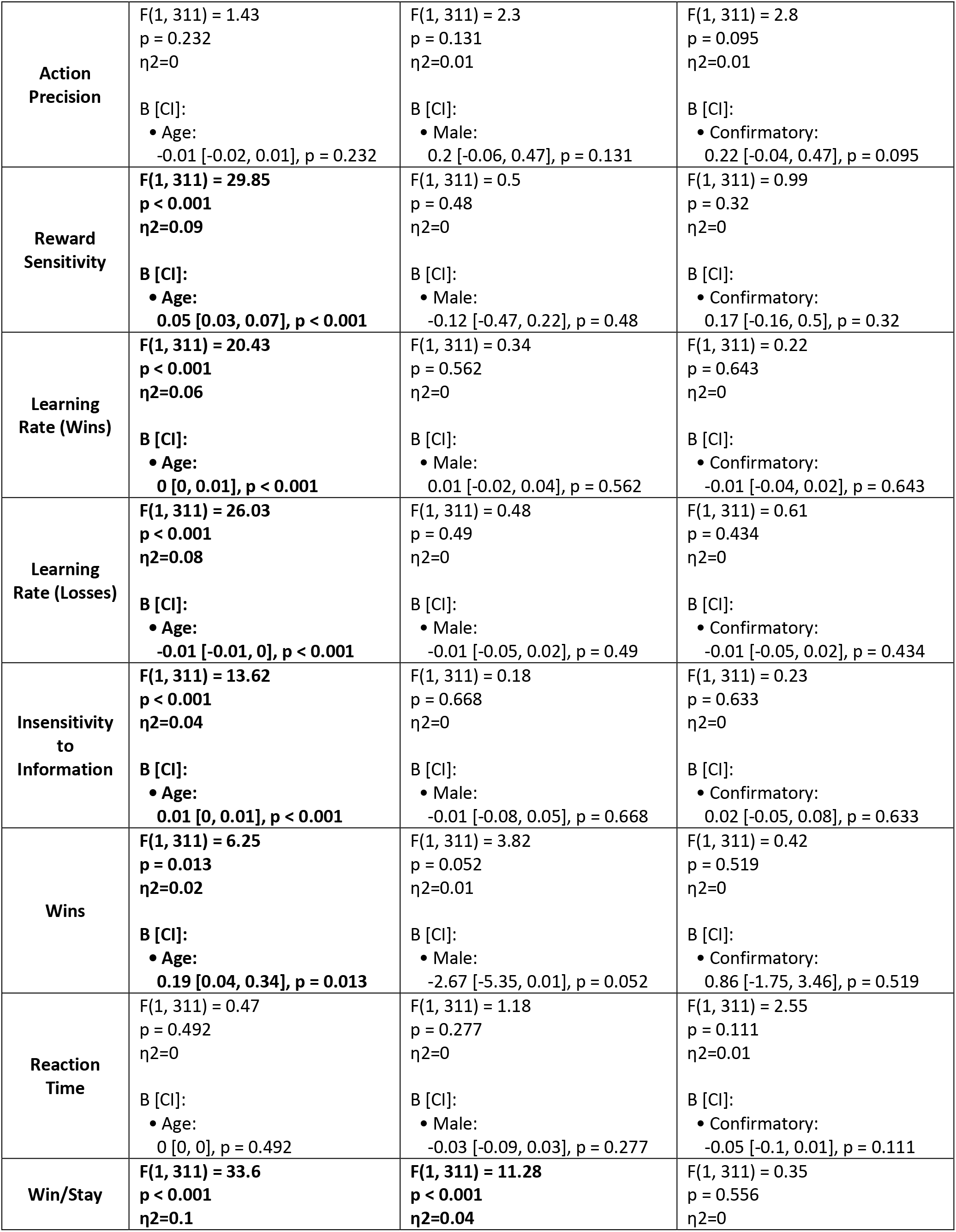
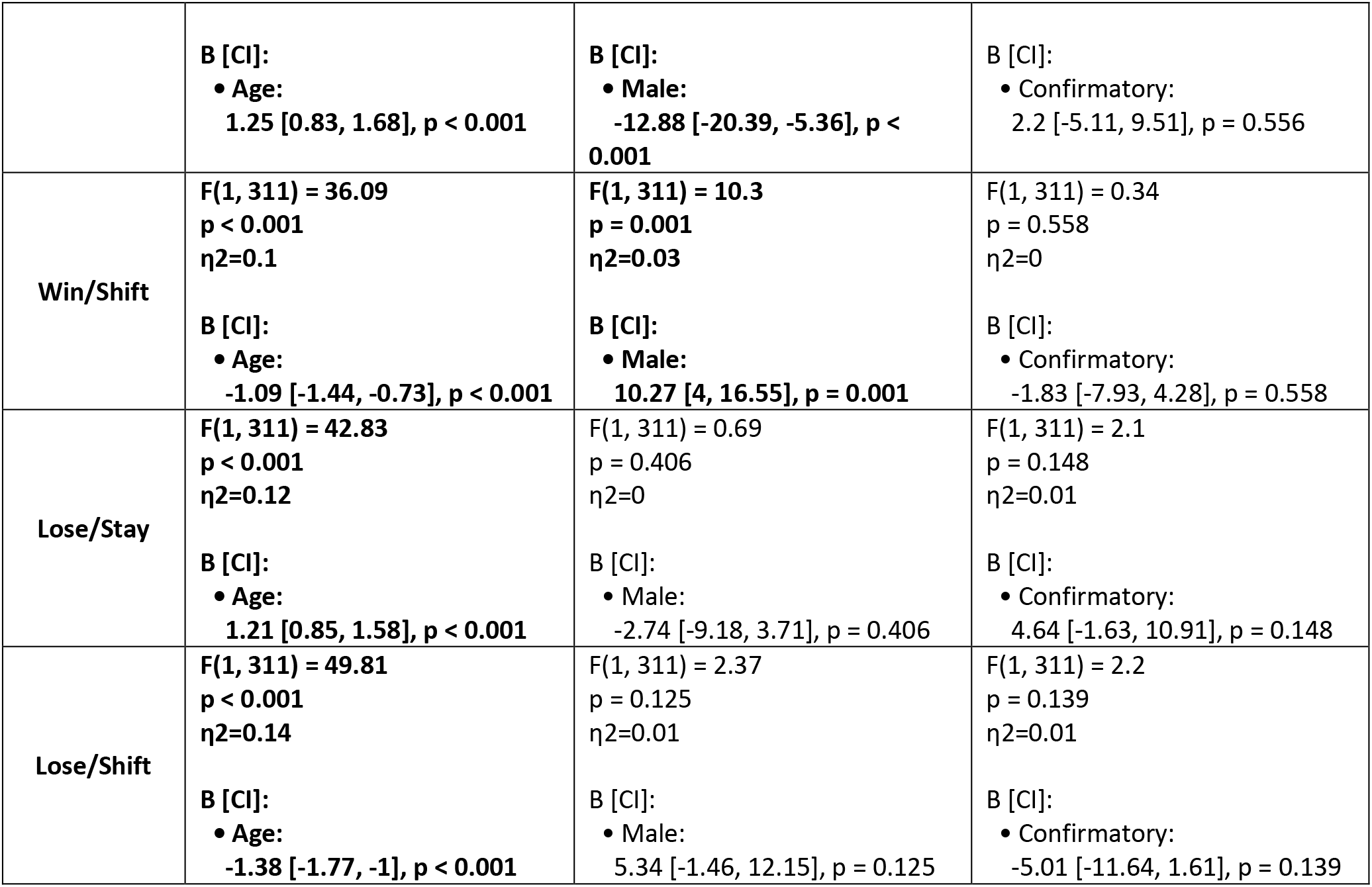
Full Linear Model Statistics Comparing SUDs Between Exploratory and Confirmatory Samples

| Effects: | Age | Sex | Sample |
| --- | --- | --- | --- |
| <u>Measures</u> |  |  |  |
| **Action Precision** | $F(1, 311) = 1.43$<br>$p = 0.232$<br>$\eta^2=0$<br>B [CI]:<br>• Age:<br>$-0.01 [-0.02, 0.01], p = 0.232$ | $F(1, 311) = 2.3$<br>$p = 0.131$<br>$\eta^2=0.01$<br>B [CI]:<br>• Male:<br>$0.2 [-0.06, 0.47], p = 0.131$ | $F(1, 311) = 2.8$<br>$p = 0.095$<br>$\eta^2=0.01$<br>B [CI]:<br>• Confirmatory:<br>$0.22 [-0.04, 0.47], p = 0.095$ |
| **Reward Sensitivity** | **$F(1, 311) = 29.85$**<br>**$p < 0.001$**<br>**$\eta^2=0.09$**<br>B [CI]:<br>• Age:<br>**$0.05 [0.03, 0.07], p < 0.001$** | $F(1, 311) = 0.5$<br>$p = 0.48$<br>$\eta^2=0$<br>B [CI]:<br>• Male:<br>$-0.12 [-0.47, 0.22], p = 0.48$ | $F(1, 311) = 0.99$<br>$p = 0.32$<br>$\eta^2=0$<br>B [CI]:<br>• Confirmatory:<br>$0.17 [-0.16, 0.5], p = 0.32$ |
| **Learning Rate (Wins)** | **$F(1, 311) = 20.43$**<br>**$p < 0.001$**<br>**$\eta^2=0.06$**<br>B [CI]:<br>• Age:<br>**$0 [0, 0.01], p < 0.001$** | $F(1, 311) = 0.34$<br>$p = 0.562$<br>$\eta^2=0$<br>B [CI]:<br>• Male:<br>$0.01 [-0.02, 0.04], p = 0.562$ | $F(1, 311) = 0.22$<br>$p = 0.643$<br>$\eta^2=0$<br>B [CI]:<br>• Confirmatory:<br>$-0.01 [-0.04, 0.02], p = 0.643$ |
| **Learning Rate (Losses)** | **$F(1, 311) = 26.03$**<br>**$p < 0.001$**<br>**$\eta^2=0.08$**<br>B [CI]:<br>• Age:<br>**$-0.01 [-0.01, 0], p < 0.001$** | $F(1, 311) = 0.48$<br>$p = 0.49$<br>$\eta^2=0$<br>B [CI]:<br>• Male:<br>$-0.01 [-0.05, 0.02], p = 0.49$ | $F(1, 311) = 0.61$<br>$p = 0.434$<br>$\eta^2=0$<br>B [CI]:<br>• Confirmatory:<br>$-0.01 [-0.05, 0.02], p = 0.434$ |
| **Insensitivity to Information** | **$F(1, 311) = 13.62$**<br>**$p < 0.001$**<br>**$\eta^2=0.04$**<br>B [CI]:<br>• Age:<br>**$0.01 [0, 0.01], p < 0.001$** | $F(1, 311) = 0.18$<br>$p = 0.668$<br>$\eta^2=0$<br>B [CI]:<br>• Male:<br>$-0.01 [-0.08, 0.05], p = 0.668$ | $F(1, 311) = 0.23$<br>$p = 0.633$<br>$\eta^2=0$<br>B [CI]:<br>• Confirmatory:<br>$0.02 [-0.05, 0.08], p = 0.633$ |
| **Wins** | **$F(1, 311) = 6.25$**<br>**$p = 0.013$**<br>**$\eta^2=0.02$**<br>B [CI]:<br>• Age:<br>**$0.19 [0.04, 0.34], p = 0.013$** | $F(1, 311) = 3.82$<br>$p = 0.052$<br>$\eta^2=0.01$<br>B [CI]:<br>• Male:<br>$-2.67 [-5.35, 0.01], p = 0.052$ | $F(1, 311) = 0.42$<br>$p = 0.519$<br>$\eta^2=0$<br>B [CI]:<br>• Confirmatory:<br>$0.86 [-1.75, 3.46], p = 0.519$ |
| **Reaction Time** | $F(1, 311) = 0.47$<br>$p = 0.492$<br>$\eta^2=0$<br>B [CI]:<br>• Age:<br>$0 [0, 0], p = 0.492$ | $F(1, 311) = 1.18$<br>$p = 0.277$<br>$\eta^2=0$<br>B [CI]:<br>• Male:<br>$-0.03 [-0.09, 0.03], p = 0.277$ | $F(1, 311) = 2.55$<br>$p = 0.111$<br>$\eta^2=0.01$<br>B [CI]:<br>• Confirmatory:<br>$-0.05 [-0.1, 0.01], p = 0.111$ |
| **Win/Stay** | **$F(1, 311) = 33.6$**<br>**$p < 0.001$**<br>**$\eta^2=0.1$** | **$F(1, 311) = 11.28$**<br>**$p < 0.001$**<br>**$\eta^2=0.04$** | $F(1, 311) = 0.35$<br>$p = 0.556$<br>$\eta^2=0$ |
| | **B [CI]:**<br>• Age:<br>1.25 [0.83, 1.68], $p < 0.001$ | **B [CI]:**<br>• Male:<br>-12.88 [-20.39, -5.36], $p < 0.001$ | **B [CI]:**<br>• Confirmatory:<br>2.2 [-5.11, 9.51], $p = 0.556$ |
| **Win/Shift** | **F(1, 311) = 36.09**<br>$p < 0.001$<br>$\eta^2=0.1$<br>**B [CI]:**<br>• Age:<br>-1.09 [-1.44, -0.73], $p < 0.001$ | **F(1, 311) = 10.3**<br>$p = 0.001$<br>$\eta^2=0.03$<br>**B [CI]:**<br>• Male:<br>10.27 [4, 16.55], $p = 0.001$ | **F(1, 311) = 0.34**<br>$p = 0.558$<br>$\eta^2=0$<br>**B [CI]:**<br>• Confirmatory:<br>-1.83 [-7.93, 4.28], $p = 0.558$ |
| **Lose/Stay** | **F(1, 311) = 42.83**<br>$p < 0.001$<br>$\eta^2=0.12$<br>**B [CI]:**<br>• Age:<br>1.21 [0.85, 1.58], $p < 0.001$ | **F(1, 311) = 0.69**<br>$p = 0.406$<br>$\eta^2=0$<br>**B [CI]:**<br>• Male:<br>-2.74 [-9.18, 3.71], $p = 0.406$ | **F(1, 311) = 2.1**<br>$p = 0.148$<br>$\eta^2=0.01$<br>**B [CI]:**<br>• Confirmatory:<br>4.64 [-1.63, 10.91], $p = 0.148$ |
| **Lose/Shift** | **F(1, 311) = 49.81**<br>$p < 0.001$<br>$\eta^2=0.14$<br>**B [CI]:**<br>• Age:<br>-1.38 [-1.77, -1], $p < 0.001$ | **F(1, 311) = 2.37**<br>$p = 0.125$<br>$\eta^2=0.01$<br>**B [CI]:**<br>• Male:<br>5.34 [-1.46, 12.15], $p = 0.125$ | **F(1, 311) = 2.2**<br>$p = 0.139$<br>$\eta^2=0.01$<br>**B [CI]:**<br>• Confirmatory:<br>-5.01 [-11.64, 1.61], $p = 0.139$ |

**Table S2.3:**
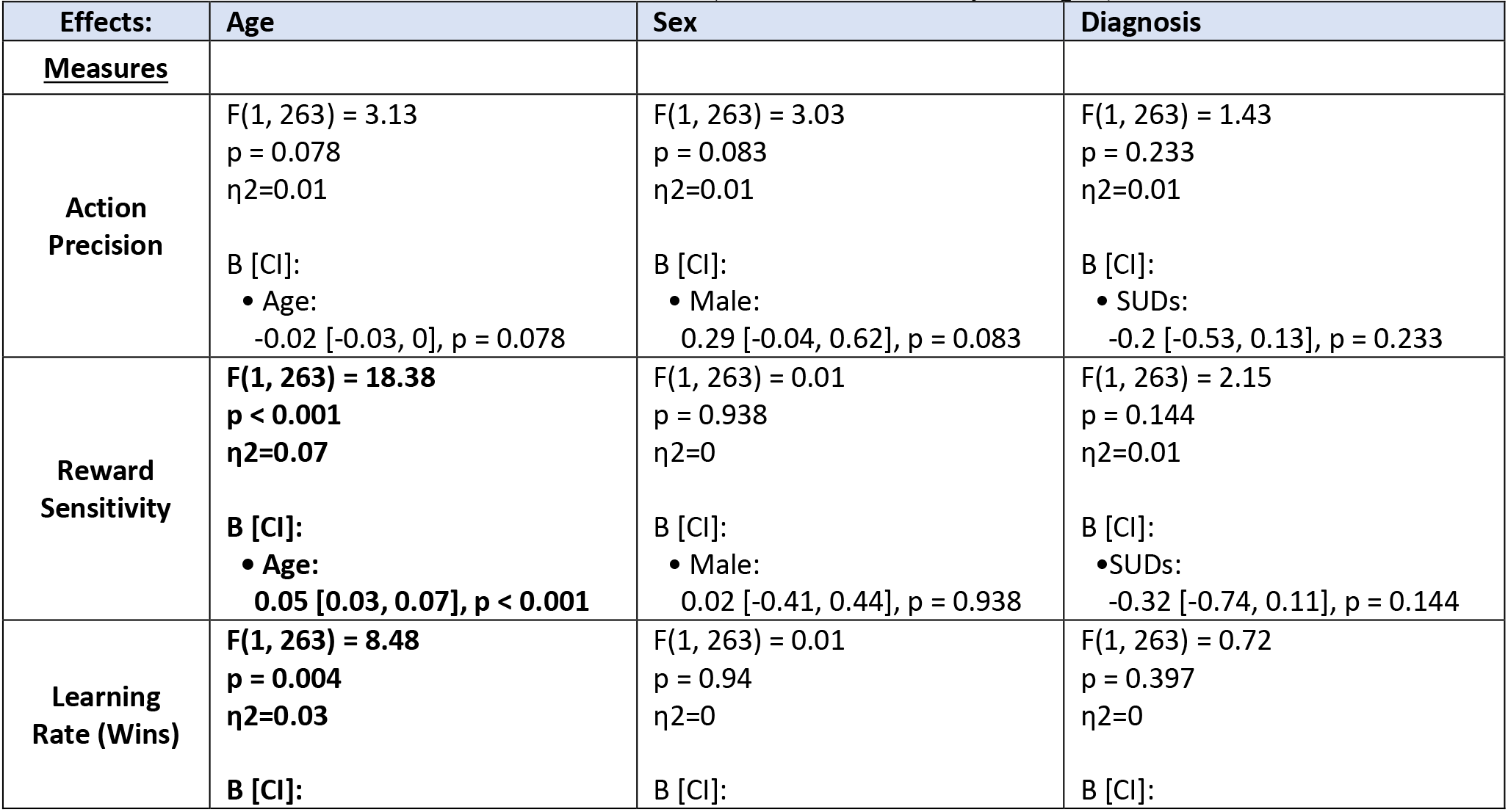
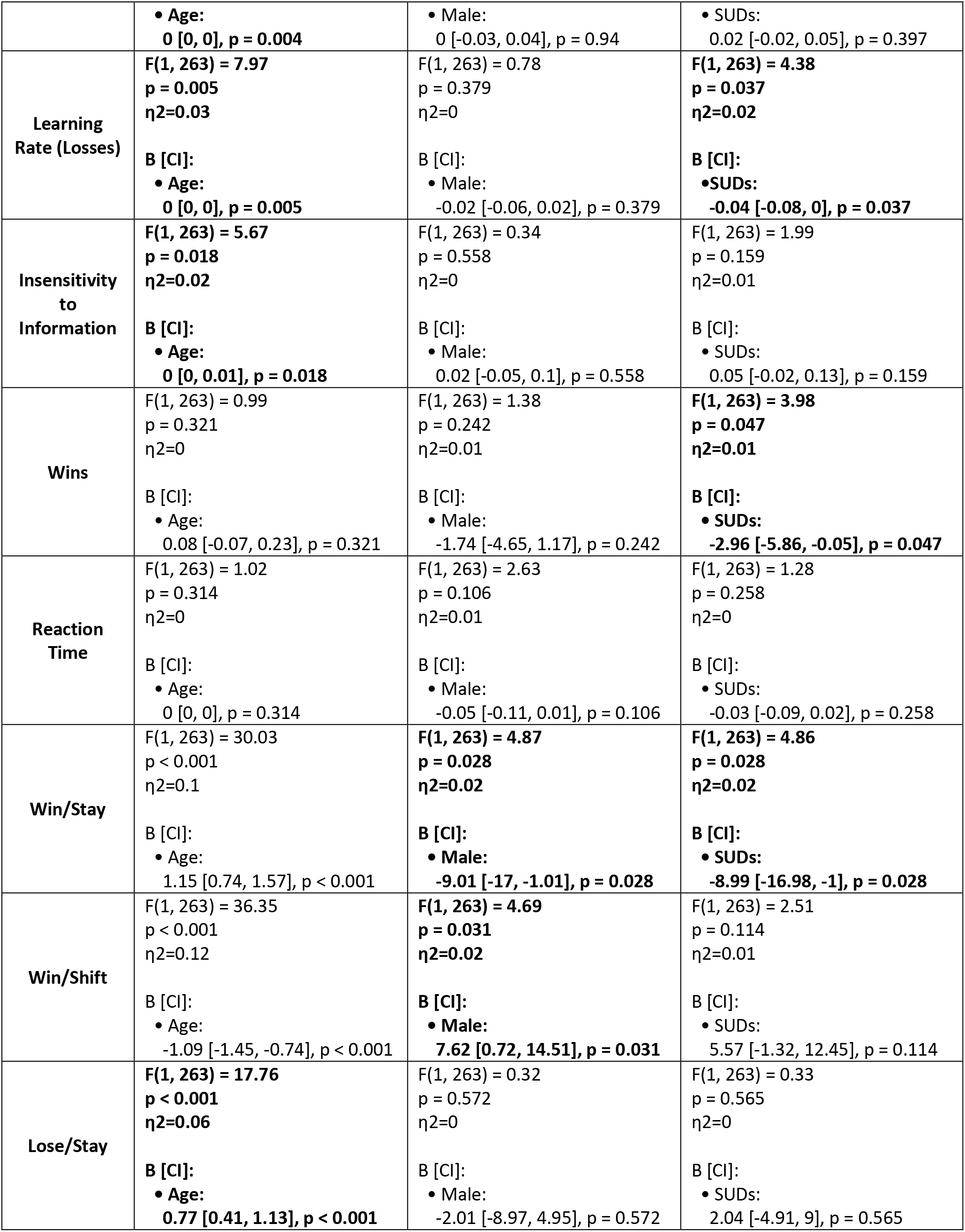
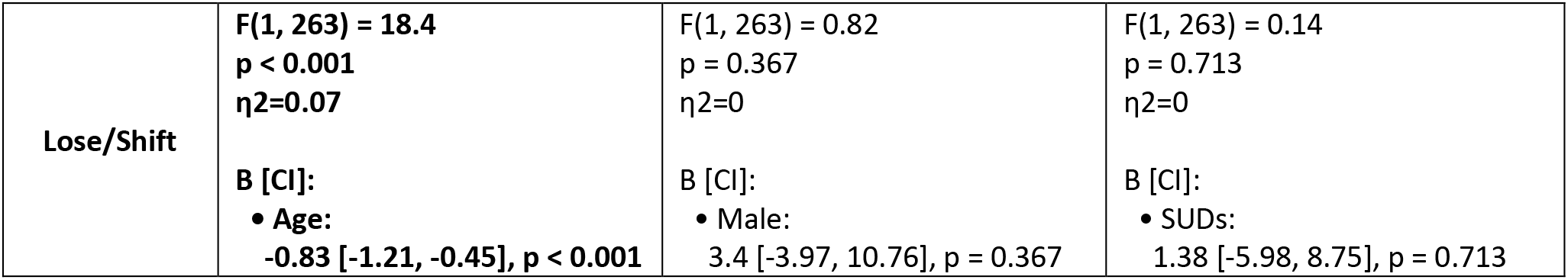
Full Linear Model Statistics (full confirmatory sample)

| **Effects:** | **Age** | **Sex** | **Diagnosis** |
| --- | --- | --- | --- |
| **Measures** |  |  |  |
| **Action Precision** | **F(1, 263) = 3.13**<br>$p = 0.078$<br>$\eta^2=0.01$<br>**B [CI]:**<br>• Age:<br>-0.02 [-0.03, 0], $p = 0.078$ | **F(1, 263) = 3.03**<br>$p = 0.083$<br>$\eta^2=0.01$<br>**B [CI]:**<br>• Male:<br>0.29 [-0.04, 0.62], $p = 0.083$ | **F(1, 263) = 1.43**<br>$p = 0.233$<br>$\eta^2=0.01$<br>**B [CI]:**<br>• SUDs:<br>-0.2 [-0.53, 0.13], $p = 0.233$ |
| **Reward Sensitivity** | **F(1, 263) = 18.38**<br>$p < 0.001$<br>$\eta^2=0.07$<br>**B [CI]:**<br>• Age:<br>0.05 [0.03, 0.07], $p < 0.001$ | **F(1, 263) = 0.01**<br>$p = 0.938$<br>$\eta^2=0$<br>**B [CI]:**<br>• Male:<br>0.02 [-0.41, 0.44], $p = 0.938$ | **F(1, 263) = 2.15**<br>$p = 0.144$<br>$\eta^2=0.01$<br>**B [CI]:**<br>• SUDs:<br>-0.32 [-0.74, 0.11], $p = 0.144$ |
| **Learning Rate (Wins)** | **F(1, 263) = 8.48**<br>$p = 0.004$<br>$\eta^2=0.03$<br>**B [CI]:** | **F(1, 263) = 0.01**<br>$p = 0.94$<br>$\eta^2=0$<br>**B [CI]:** | **F(1, 263) = 0.72**<br>$p = 0.397$<br>$\eta^2=0$<br>**B [CI]:** |
|  | Age:<br>**0 [0, 0], p = 0.004** | Male:<br>0 [-0.03, 0.04], p = 0.94 | SUDs:<br>0.02 [-0.02, 0.05], p = 0.397 |
| Learning Rate (Losses) | **F(1, 263) = 7.97**<br>**p = 0.005**<br>**$\eta^2=0.03$**<br>**B [CI]:**<br>Age:<br>**0 [0, 0], p = 0.005** | F(1, 263) = 0.78<br>p = 0.379<br>$\eta^2=0$<br>B [CI]:<br>Male:<br>-0.02 [-0.06, 0.02], p = 0.379 | **F(1, 263) = 4.38**<br>**p = 0.037**<br>**$\eta^2=0.02$**<br>**B [CI]:**<br>SUDs:<br>**-0.04 [-0.08, 0], p = 0.037** |
| Insensitivity to Information | **F(1, 263) = 5.67**<br>**p = 0.018**<br>**$\eta^2=0.02$**<br>**B [CI]:**<br>Age:<br>**0 [0, 0.01], p = 0.018** | F(1, 263) = 0.34<br>p = 0.558<br>$\eta^2=0$<br>B [CI]:<br>Male:<br>0.02 [-0.05, 0.1], p = 0.558 | F(1, 263) = 1.99<br>p = 0.159<br>$\eta^2=0.01$<br>B [CI]:<br>SUDs:<br>0.05 [-0.02, 0.13], p = 0.159 |
| Wins | F(1, 263) = 0.99<br>p = 0.321<br>$\eta^2=0$<br>B [CI]:<br>Age:<br>0.08 [-0.07, 0.23], p = 0.321 | F(1, 263) = 1.38<br>p = 0.242<br>$\eta^2=0.01$<br>B [CI]:<br>Male:<br>-1.74 [-4.65, 1.17], p = 0.242 | **F(1, 263) = 3.98**<br>**p = 0.047**<br>**$\eta^2=0.01$**<br>**B [CI]:**<br>SUDs:<br>**-2.96 [-5.86, -0.05], p = 0.047** |
| Reaction Time | F(1, 263) = 1.02<br>p = 0.314<br>$\eta^2=0$<br>B [CI]:<br>Age:<br>0 [0, 0], p = 0.314 | F(1, 263) = 2.63<br>p = 0.106<br>$\eta^2=0.01$<br>B [CI]:<br>Male:<br>-0.05 [-0.11, 0.01], p = 0.106 | F(1, 263) = 1.28<br>p = 0.258<br>$\eta^2=0$<br>B [CI]:<br>SUDs:<br>-0.03 [-0.09, 0.02], p = 0.258 |
| Win/Stay | F(1, 263) = 30.03<br>p < 0.001<br>$\eta^2=0.1$<br>B [CI]:<br>Age:<br>1.15 [0.74, 1.57], p < 0.001 | **F(1, 263) = 4.87**<br>**p = 0.028**<br>**$\eta^2=0.02$**<br>**B [CI]:**<br>Male:<br>**-9.01 [-17, -1.01], p = 0.028** | **F(1, 263) = 4.86**<br>**p = 0.028**<br>**$\eta^2=0.02$**<br>**B [CI]:**<br>SUDs:<br>**-8.99 [-16.98, -1], p = 0.028** |
| Win/Shift | F(1, 263) = 36.35<br>p < 0.001<br>$\eta^2=0.12$<br>B [CI]:<br>Age:<br>-1.09 [-1.45, -0.74], p < 0.001 | **F(1, 263) = 4.69**<br>**p = 0.031**<br>**$\eta^2=0.02$**<br>**B [CI]:**<br>Male:<br>**7.62 [0.72, 14.51], p = 0.031** | F(1, 263) = 2.51<br>p = 0.114<br>$\eta^2=0.01$<br>B [CI]:<br>SUDs:<br>5.57 [-1.32, 12.45], p = 0.114 |
| Lose/Stay | **F(1, 263) = 17.76**<br>**p < 0.001**<br>**$\eta^2=0.06$**<br>**B [CI]:**<br>Age:<br>**0.77 [0.41, 1.13], p < 0.001** | F(1, 263) = 0.32<br>p = 0.572<br>$\eta^2=0$<br>B [CI]:<br>Male:<br>-2.01 [-8.97, 4.95], p = 0.572 | F(1, 263) = 0.33<br>p = 0.565<br>$\eta^2=0$<br>B [CI]:<br>SUDs:<br>2.04 [-4.91, 9], p = 0.565 |
| **Lose/Shift** | **F(1, 263) = 18.4**<br>**p < 0.001**<br>**η<sup>2</sup>=0.07**<br>**B [CI]:**<br>• Age:<br>-0.83 [-1.21, -0.45], p < 0.001 | F(1, 263) = 0.82<br>p = 0.367<br>η<sup>2</sup>=0<br>B [CI]:<br>• Male:<br>3.4 [-3.97, 10.76], p = 0.367 | F(1, 263) = 0.14<br>p = 0.713<br>η<sup>2</sup>=0<br>B [CI]:<br>• SUDs:<br>1.38 [-5.98, 8.75], p = 0.713 |

**Table S2.4:**
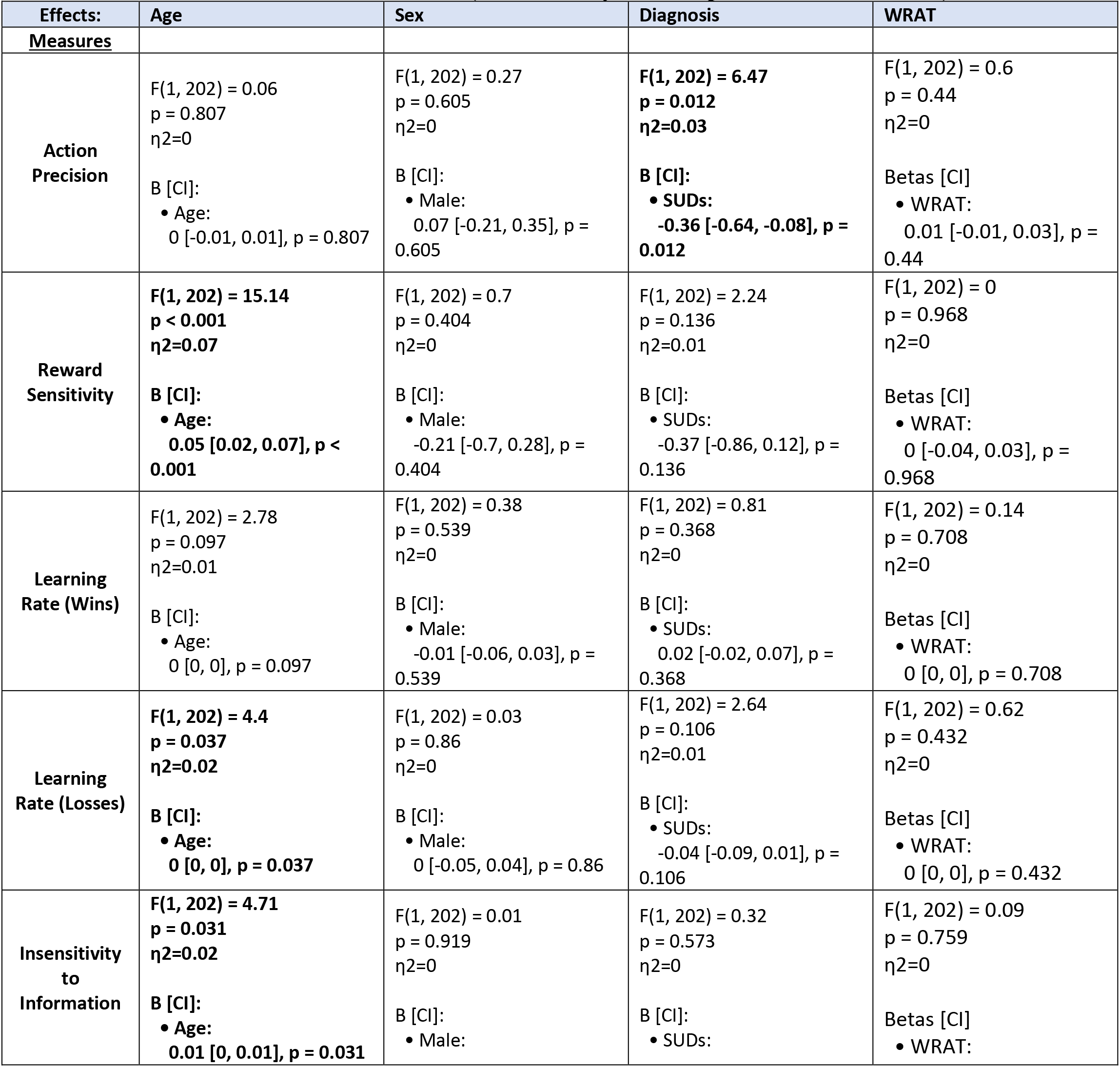
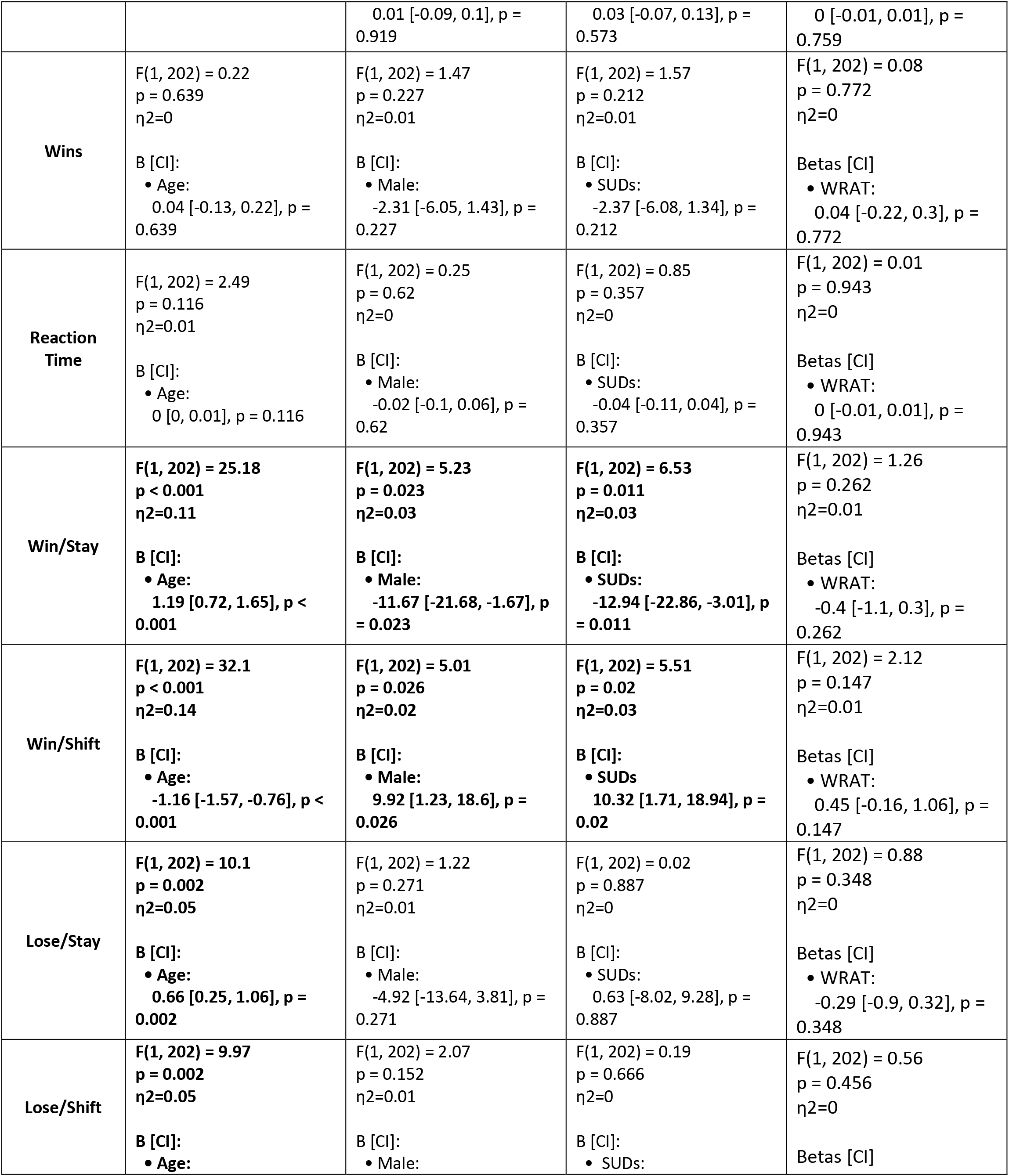
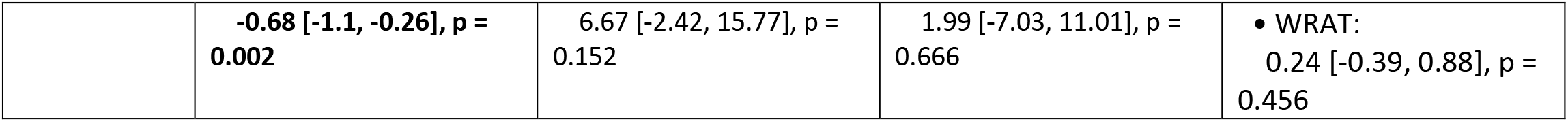
Full Linear Model Statistics (confirmatory sub-sample with WRAT scores)

| Effects: | Age | Sex | Diagnosis | WRAT |
| --- | --- | --- | --- | --- |
| Measures |  |  |  |  |
| **Action Precision** | F(1, 202) = 0.06<br>p = 0.807<br>η<sup>2</sup>=0<br>B [CI]:<br>• Age:<br>0 [-0.01, 0.01], p = 0.807 | F(1, 202) = 0.27<br>p = 0.605<br>η<sup>2</sup>=0<br>B [CI]:<br>• Male:<br>0.07 [-0.21, 0.35], p = 0.605 | **F(1, 202) = 6.47**<br>**p = 0.012**<br>**η<sup>2</sup>=0.03**<br>**B [CI]:**<br>• SUDs:<br>-0.36 [-0.64, -0.08], p = 0.012 | F(1, 202) = 0.6<br>p = 0.44<br>η<sup>2</sup>=0<br>Betas [CI]<br>• WRAT:<br>0.01 [-0.01, 0.03], p = 0.44 |
| **Reward Sensitivity** | **F(1, 202) = 15.14**<br>**p < 0.001**<br>**η<sup>2</sup>=0.07**<br>**B [CI]:**<br>• Age:<br>0.05 [0.02, 0.07], p < 0.001 | F(1, 202) = 0.7<br>p = 0.404<br>η<sup>2</sup>=0<br>B [CI]:<br>• Male:<br>-0.21 [-0.7, 0.28], p = 0.404 | F(1, 202) = 2.24<br>p = 0.136<br>η<sup>2</sup>=0.01<br>B [CI]:<br>• SUDs:<br>-0.37 [-0.86, 0.12], p = 0.136 | F(1, 202) = 0<br>p = 0.968<br>η<sup>2</sup>=0<br>Betas [CI]<br>• WRAT:<br>0 [-0.04, 0.03], p = 0.968 |
| **Learning Rate (Wins)** | F(1, 202) = 2.78<br>p = 0.097<br>η<sup>2</sup>=0.01<br>B [CI]:<br>• Age:<br>0 [0, 0], p = 0.097 | F(1, 202) = 0.38<br>p = 0.539<br>η<sup>2</sup>=0<br>B [CI]:<br>• Male:<br>-0.01 [-0.06, 0.03], p = 0.539 | F(1, 202) = 0.81<br>p = 0.368<br>η<sup>2</sup>=0<br>B [CI]:<br>• SUDs:<br>0.02 [-0.02, 0.07], p = 0.368 | F(1, 202) = 0.14<br>p = 0.708<br>η<sup>2</sup>=0<br>Betas [CI]<br>• WRAT:<br>0 [0, 0], p = 0.708 |
| **Learning Rate (Losses)** | **F(1, 202) = 4.4**<br>**p = 0.037**<br>**η<sup>2</sup>=0.02**<br>**B [CI]:**<br>• Age:<br>0 [0, 0], p = 0.037 | F(1, 202) = 0.03<br>p = 0.86<br>η<sup>2</sup>=0<br>B [CI]:<br>• Male:<br>0 [-0.05, 0.04], p = 0.86 | F(1, 202) = 2.64<br>p = 0.106<br>η<sup>2</sup>=0.01<br>B [CI]:<br>• SUDs:<br>-0.04 [-0.09, 0.01], p = 0.106 | F(1, 202) = 0.62<br>p = 0.432<br>η<sup>2</sup>=0<br>Betas [CI]<br>• WRAT:<br>0 [0, 0], p = 0.432 |
| **Insensitivity to Information** | **F(1, 202) = 4.71**<br>**p = 0.031**<br>**η<sup>2</sup>=0.02**<br>**B [CI]:**<br>• Age:<br>0.01 [0, 0.01], p = 0.031 | F(1, 202) = 0.01<br>p = 0.919<br>η<sup>2</sup>=0<br>B [CI]:<br>• Male: | F(1, 202) = 0.32<br>p = 0.573<br>η<sup>2</sup>=0<br>B [CI]:<br>• SUDs: | F(1, 202) = 0.09<br>p = 0.759<br>η<sup>2</sup>=0<br>Betas [CI]<br>• WRAT: |
|  |  | 0.01 [-0.09, 0.1], p = 0.919 | 0.03 [-0.07, 0.13], p = 0.573 | 0 [-0.01, 0.01], p = 0.759 |
| **Wins** | F(1, 202) = 0.22<br>p = 0.639<br>$\eta^2=0$<br>B [CI]:<br>• Age:<br>0.04 [-0.13, 0.22], p = 0.639 | F(1, 202) = 1.47<br>p = 0.227<br>$\eta^2=0.01$<br>B [CI]:<br>• Male:<br>-2.31 [-6.05, 1.43], p = 0.227 | F(1, 202) = 1.57<br>p = 0.212<br>$\eta^2=0.01$<br>B [CI]:<br>• SUDs:<br>-2.37 [-6.08, 1.34], p = 0.212 | F(1, 202) = 0.08<br>p = 0.772<br>$\eta^2=0$<br>Betas [CI]<br>• WRAT:<br>0.04 [-0.22, 0.3], p = 0.772 |
| **Reaction Time** | F(1, 202) = 2.49<br>p = 0.116<br>$\eta^2=0.01$<br>B [CI]:<br>• Age:<br>0 [0, 0.01], p = 0.116 | F(1, 202) = 0.25<br>p = 0.62<br>$\eta^2=0$<br>B [CI]:<br>• Male:<br>-0.02 [-0.1, 0.06], p = 0.62 | F(1, 202) = 0.85<br>p = 0.357<br>$\eta^2=0$<br>B [CI]:<br>• SUDs:<br>-0.04 [-0.11, 0.04], p = 0.357 | F(1, 202) = 0.01<br>p = 0.943<br>$\eta^2=0$<br>Betas [CI]<br>• WRAT:<br>0 [-0.01, 0.01], p = 0.943 |
| **Win/Stay** | F(1, 202) = 25.18<br>p < 0.001<br>$\eta^2=0.11$<br>B [CI]:<br>• Age:<br>1.19 [0.72, 1.65], p < 0.001 | F(1, 202) = 5.23<br>p = 0.023<br>$\eta^2=0.03$<br>B [CI]:<br>• Male:<br>-11.67 [-21.68, -1.67], p = 0.023 | F(1, 202) = 6.53<br>p = 0.011<br>$\eta^2=0.03$<br>B [CI]:<br>• SUDs:<br>-12.94 [-22.86, -3.01], p = 0.011 | F(1, 202) = 1.26<br>p = 0.262<br>$\eta^2=0.01$<br>Betas [CI]<br>• WRAT:<br>-0.4 [-1.1, 0.3], p = 0.262 |
| **Win/Shift** | F(1, 202) = 32.1<br>p < 0.001<br>$\eta^2=0.14$<br>B [CI]:<br>• Age:<br>-1.16 [-1.57, -0.76], p < 0.001 | F(1, 202) = 5.01<br>p = 0.026<br>$\eta^2=0.02$<br>B [CI]:<br>• Male:<br>9.92 [1.23, 18.6], p = 0.026 | F(1, 202) = 5.51<br>p = 0.02<br>$\eta^2=0.03$<br>B [CI]:<br>• SUDs:<br>10.32 [1.71, 18.94], p = 0.02 | F(1, 202) = 2.12<br>p = 0.147<br>$\eta^2=0.01$<br>Betas [CI]<br>• WRAT:<br>0.45 [-0.16, 1.06], p = 0.147 |
| **Lose/Stay** | F(1, 202) = 10.1<br>p = 0.002<br>$\eta^2=0.05$<br>B [CI]:<br>• Age:<br>0.66 [0.25, 1.06], p = 0.002 | F(1, 202) = 1.22<br>p = 0.271<br>$\eta^2=0.01$<br>B [CI]:<br>• Male:<br>-4.92 [-13.64, 3.81], p = 0.271 | F(1, 202) = 0.02<br>p = 0.887<br>$\eta^2=0$<br>B [CI]:<br>• SUDs:<br>0.63 [-8.02, 9.28], p = 0.887 | F(1, 202) = 0.88<br>p = 0.348<br>$\eta^2=0$<br>Betas [CI]<br>• WRAT:<br>-0.29 [-0.9, 0.32], p = 0.348 |
| **Lose/Shift** | F(1, 202) = 9.97<br>p = 0.002<br>$\eta^2=0.05$<br>B [CI]:<br>• Age: | F(1, 202) = 2.07<br>p = 0.152<br>$\eta^2=0.01$<br>B [CI]:<br>• Male: | F(1, 202) = 0.19<br>p = 0.666<br>$\eta^2=0$<br>B [CI]:<br>• SUDs: | F(1, 202) = 0.56<br>p = 0.456<br>$\eta^2=0$<br>Betas [CI] |
|  | **-0.68 [-1.1, -0.26], p = 0.002** | 6.67 [-2.42, 15.77], p = 0.152 | 1.99 [-7.03, 11.01], p = 0.666 | WRAT: 0.24 [-0.39, 0.88], p = 0.456 |

